# The potential impact, cost and cost-effectiveness of tuberculosis interventions - a modelling exercise

**DOI:** 10.1101/2025.09.02.25334943

**Authors:** Katherine C. Horton, Alvaro Schwalb, Martin J. Harker, Lara Goscé, Elena Venero-Garcia, Lily O’Brien, Arkaprabha Gun, Tom Sumner, C. Finn McQuaid, Rebecca A. Clark, Tomos O. Prys-Jones, Roel Bakker, Yiran E Liu, Mmamapudi Kubjane, Christian Lienhardt, Richard G. White, Rein M.G.J. Houben

## Abstract

**Background:** While a range of interventions exist for tuberculosis prevention, screening, diagnosis, and treatment, their potential population impact and cost-effectiveness are seldom directly compared, or evaluated between settings with different background TB epidemiology and structural drivers.

**Methods:** We calibrated a deterministic TB model to epidemiological indicators in Brazil, India, and South Africa. We implemented seven interventions across countries focusing on prevention, screening and diagnosis, and treatment of TB, as well as TB screening in prisons in Brazil and nutritional supplementation in India. We standardised scale-up (2025-2030), coverage (80% of target population), and strength of evidence for epidemiological impact using published efficacy data. We estimated epidemiological impact and incremental cost-effectiveness ratios (ICERs), expressed as costs per disability-adjusted life year (DALY) averted by 2050.

**Results:** Only three interventions prevented >10% of incident TB episodes by 2050: vaccination (median 15-28% across countries), symptom-agnostic community-wide screening (32-38%) and screening in prisons (23%). The impact of other interventions was more limited, ranging from 0% (shortened drug-susceptible treatment) to 5% (nutritional supplementation). ICERs varied widely by intervention and setting. Shortened drug-resistant treatment was cost-saving across settings, with the next lowest ICERs for prison screening in Brazil (72 USD/DALY) and nutritional supplementation in India (167 USD/DALY). Within each country, both low-cost community-wide screening and TB vaccine campaigns had lower USD/DALY than TB preventive treatment.

**Conclusion:** Interventions with meaningful epidemiological impact can also be cost-effective, but need to target populations beyond clinic-diagnosed individuals or their households. Achieving such potential requires a priority shift in funding, policy and product development.

## Background

An estimated 10 million people develop tuberculosis (TB) each year, resulting in 1.3 million deaths. ^1^ Although TB trends have shown a slow decline, their recovery from post-Covid-19 TB programme disruptions is now again threatened by global funding cuts ^2,3^ and potential increases in known structural drivers of TB such as HIV, undernutrition and incarceration.^4,5^

The armamentarium against the global TB epidemic has expanded substantially in the past decade, including expanded access to PCR-based tools for rapid screening and diagnosis ^6^, shortened treatment for drug-susceptible (DS) ^7^ and, in particular, drug-resistant (DR) TB ^8^, and promising TB vaccine candidates to prevent progression to disease. ^9^ In addition, interventions addressing key structural drivers of TB – such as anti-retroviral therapy (ART) for individuals co-infected with HIV, nutritional supplementation, and systematic screening for TB in prisons have shown efficacy to reduce disease burden. ^10,11^

However, at a time of constrained budgets for TB care and management globally, an expanding toolkit presents more choices to make in terms of investments, i.e. both in terms of research and product development as well as selecting interventions to implement at national programme level. For example, policy makers must decide whether the health benefit of nutritional supplementation is worth the added implementation cost to a TB programme, whilst funding and research agencies must weigh the large investment required for a phase 3 vaccine trial against the costs of developing and testing new approaches to community-wide screening. ^9,10,12^

Evidence of direct comparisons between such different interventions is, however, scarce. Development of new tools is largely guided by Target Product Profiles (TPPs), which often focus on the cost and benefits of a single class of interventions or a single product ^13,14^. Furthermore, TPP-associated modelling exercises are often limited, considering only the health benefit of single products or interventions, and often within targeted sub-populations rather than the wider population. ^14,15^

Also, few projects have compared the potential impact of interventions across domains of prevention, screening and diagnosis, and treatment, ^16,17^ so policy-directed discussions most often focus on individual intervention domains. ^18–21^ Lastly, most multi-intervention studies were conducted over a decade ago, and reflected policy choices for a single country, ^16,22^ region ^19^ or aggregated global population. ^17^ It is unclear whether such findings would still hold true in reference to current tools, or remain valid when considering wide geographical heterogeneity in TB burden, trends, and distribution of structural drivers of TB. In addition, these studies seldom considered costs, a critical component for policy and product development decisions. Combined with recent progress in our understanding of TB natural history ^23^, there is a high need to revisit the question of potential epidemiological benefit and costs of current TB interventions.

In this study, we compared the epidemiological and budgetary impact, as well as cost-effectiveness, of a standardised set of interventions targeting the intervention domains of TB Prevention, Screening and Diagnosis, and Treatment, taking into account locally relevant structural determinants.

## Methods

### Epidemiological archetypes

We considered three country archetypes to explore robustness of findings across diverse geography and TB epidemiology, including current burden, historical trends and structural drivers. We selected Brazil, India, and South Africa, where estimated TB incidence in 2023 ranged between 49 and 427 per 100,000. ^1^ In each country, we identified the structural determinant with the highest estimated population attributable fraction (PAF), which were incarceration (36.9%) in Brazil ^5^, undernutrition (13-45.2%) in India ^1,24^ and HIV (49%) in South Africa. ^1^

### Principles of intervention comparison

As our objective was to provide a standardised comparison of TB interventions across domains of Prevention, Screening and Diagnosis, and Treatment, we used a set of principles for the choice of intervention within each intervention category and implementation in the model. Firstly, interventions were included based on the availability of published empirical evidence for the target population, epidemiological mechanism, and efficacy. This, for example, excluded products with as yet unknown efficacy, such as the recently proposed Pan-TB treatment. ^15^ Secondly, we standardised implementation where possible, aligning start year (2025), scale-up duration (2025-2030) and shape (linear), as well as the coverage target (80% of the target population) and time horizon to accumulate potential indirect effects (2030-2050). Finally, coverage was assumed to apply to the complete process, from initiation to completion (e.g. initiation to completion of TB preventive treatment (TPT)). While this is a simplification of existing operational challenges ^25^ or availability (e.g. TPT or TB vaccination), it enables a full comparison of potential epidemiological impact rather than a mix between potential impact and operational constraints specific to individual interventions.

### Choice and data on interventions

In each country, we compared interventions covering key intervention domains. For the Prevention domain, this included a prevention-of-disease (POD) vaccine ^9^ and TPT. ^26^ Within the Screening and Diagnosis domain, we included a symptom-agnostic community-wide screening intervention ^12^, expanding new diagnostic tools in TB clinics ^6^ and expanded drug susceptibility testing (DST). ^6^ For Treatment, we included the introduction of a shorter, non-inferior (i.e. same treatment efficacy) regimen for DS-TB ^7^ and a shortened and more efficacious regimen for DR-TB. ^8^ Interventions targeting the country-specific structural drivers included annual symptom-agnostic screening of incarcerated individuals in Brazil ^11^, and a nationwide implementation of the recently completed RATIONS nutritional intervention in India.^10^ We did not model a separate intervention for South Africa as the key intervention addressing HIV (wide availability of ART) is already implemented. Table 1 describes the interventions for each category. Full details, including test positivity of diagnostic algorithms, can be found in supp mat S2, section 2.

**TABLE 1:**
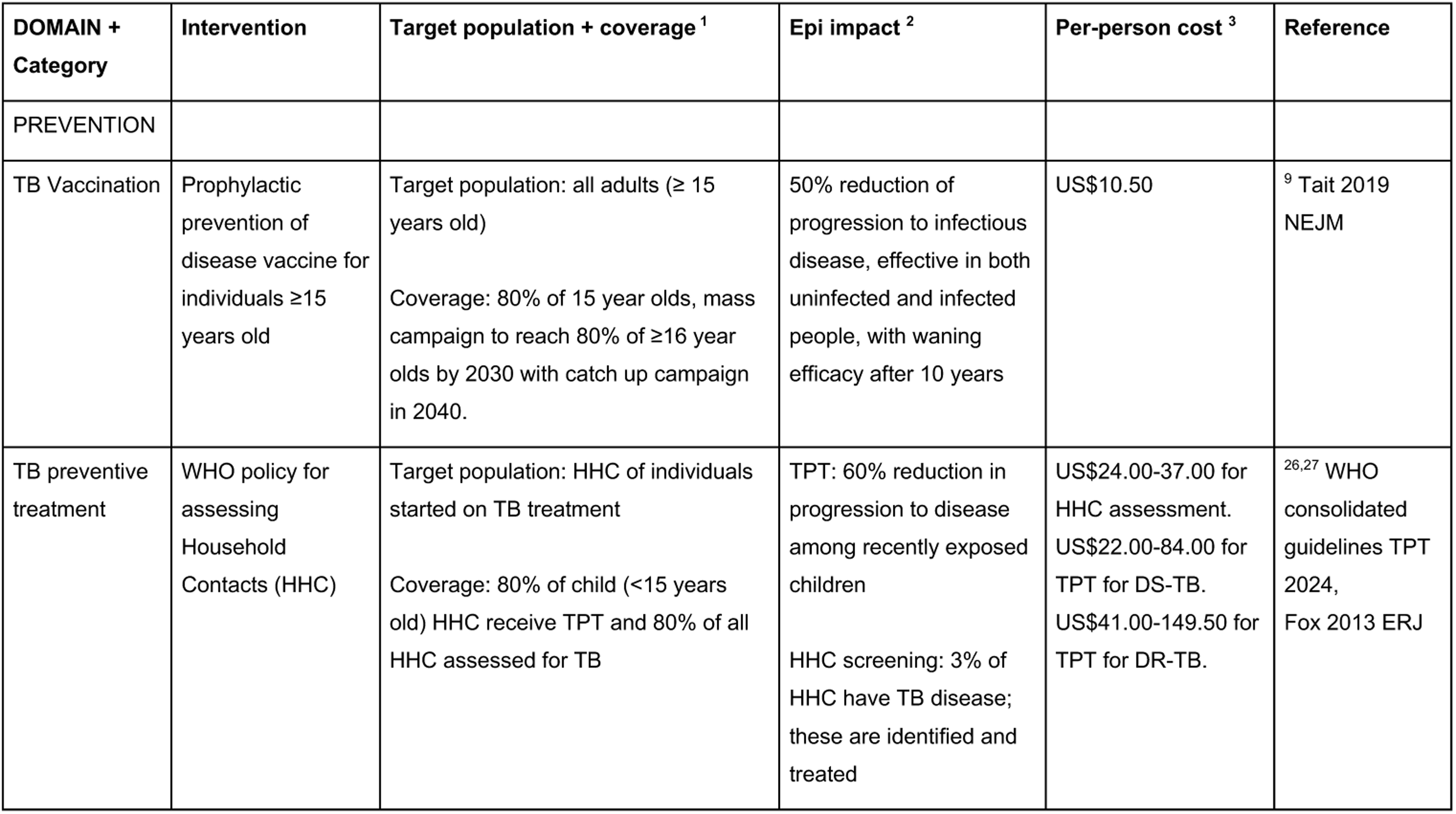
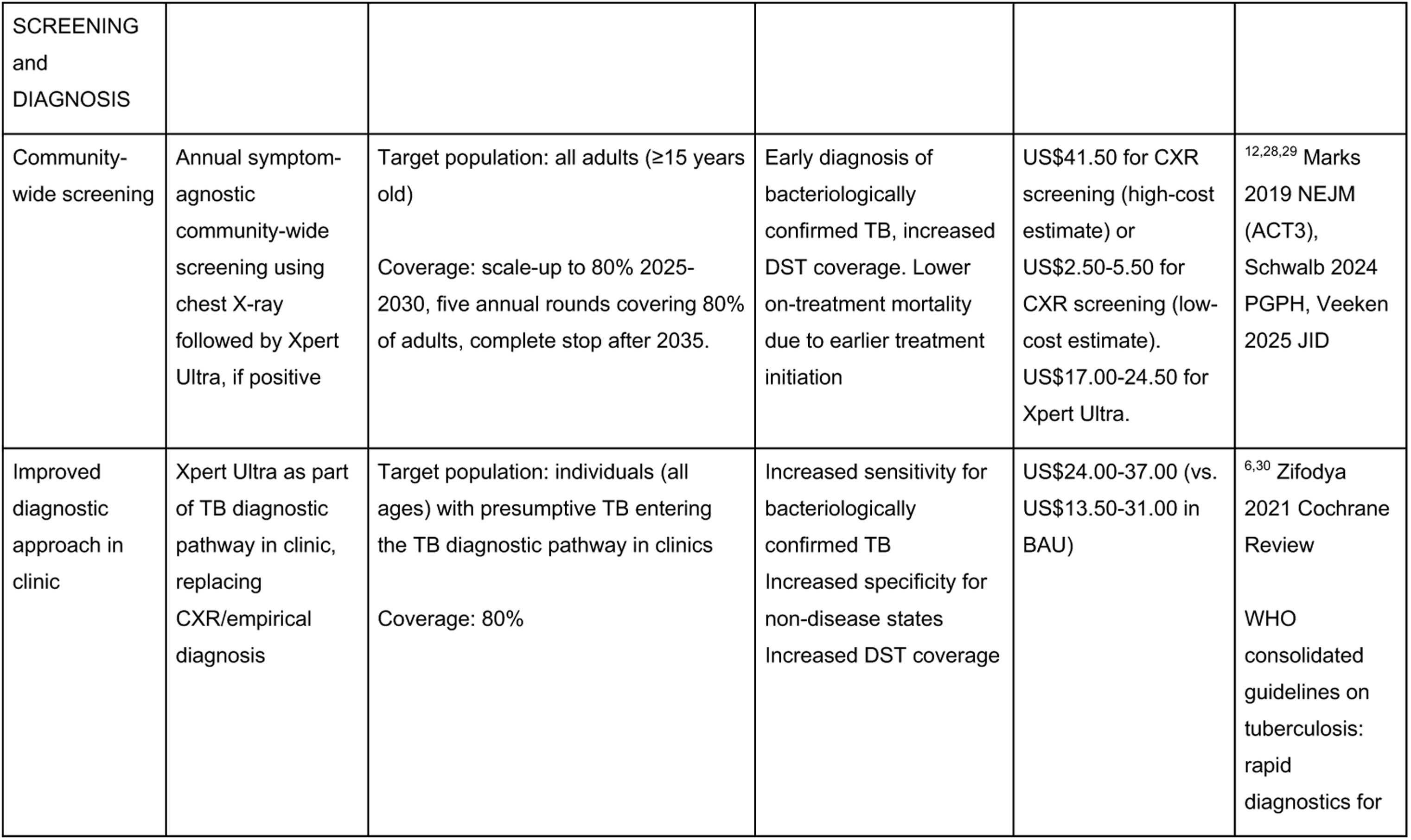
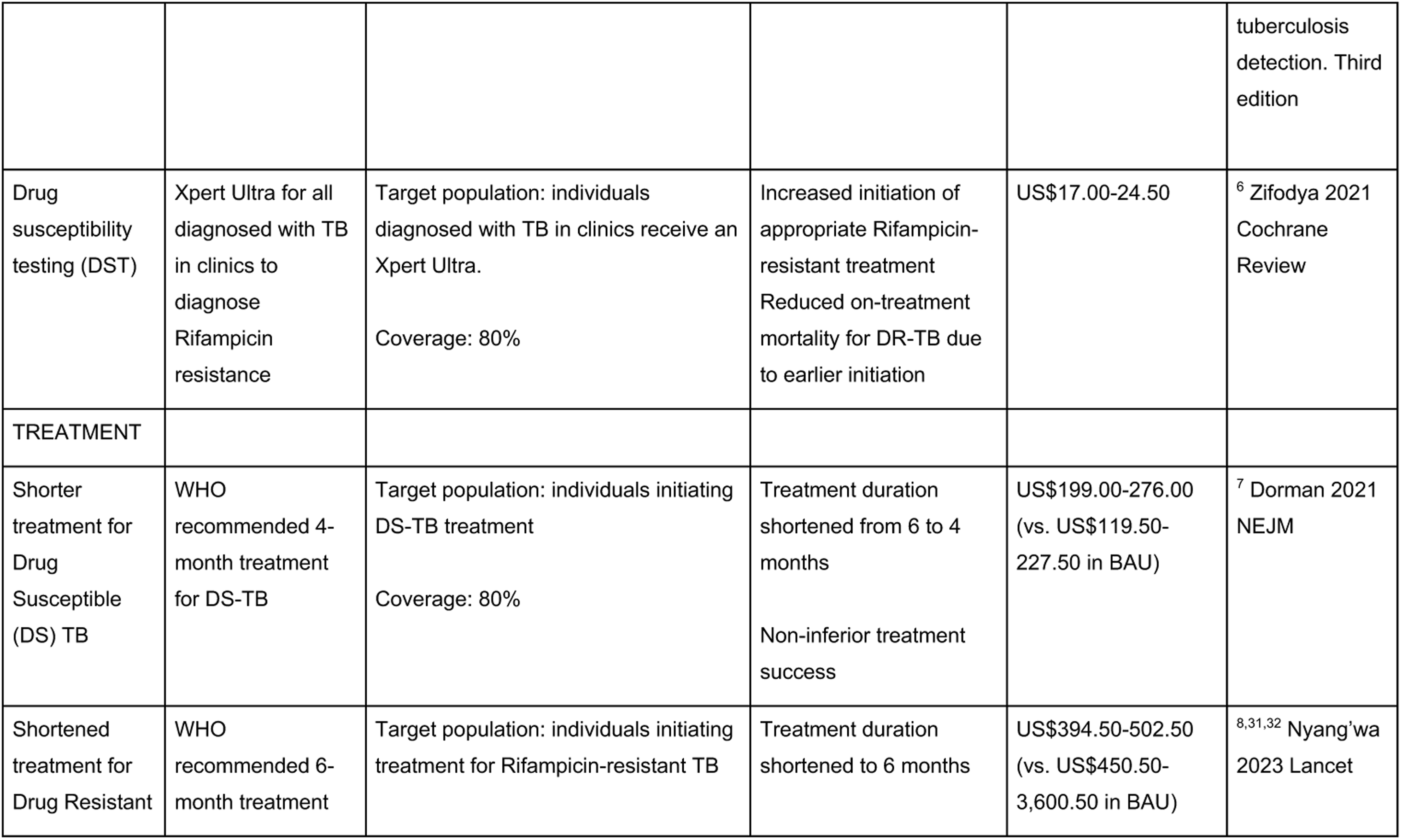
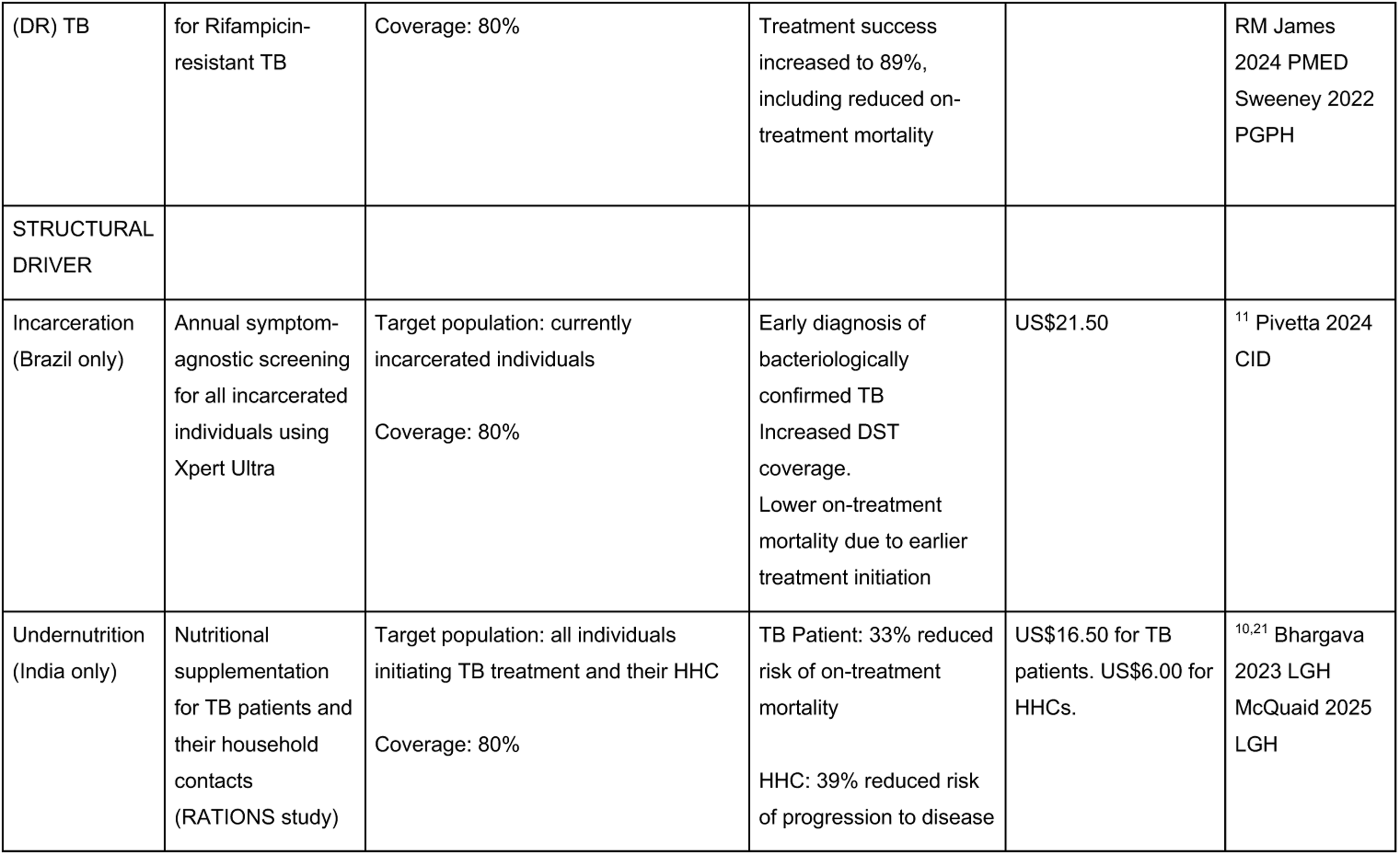
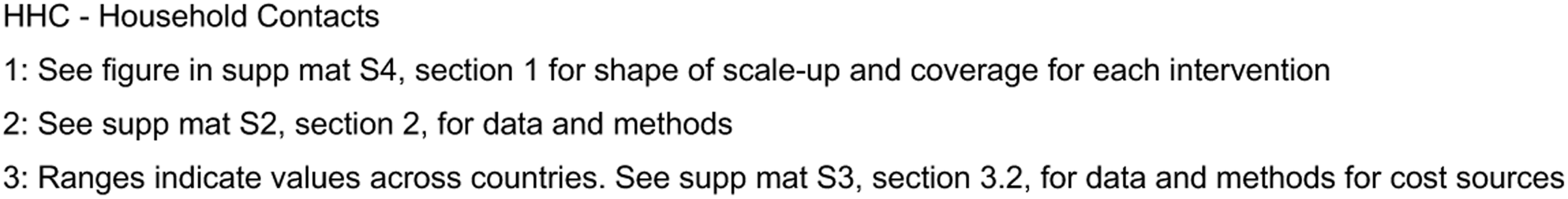
Interventions included.

| **DOMAIN + Category** | **Intervention** | **Target population + coverage <sup>1</sup>** | **Epi impact <sup>2</sup>** | **Per-person cost <sup>3</sup>** | **Reference** |
| --- | --- | --- | --- | --- | --- |
| PREVENTION |  |  |  |  |  |
| TB Vaccination | Prophylactic prevention of disease vaccine for individuals ≥15 years old | Target population: all adults (≥ 15 years old)<br>Coverage: 80% of 15 year olds, mass campaign to reach 80% of ≥16 year olds by 2030 with catch up campaign in 2040. | 50% reduction of progression to infectious disease, effective in both uninfected and infected people, with waning efficacy after 10 years | US\$10.50 | <sup>9</sup> Tait 2019 NEJM |
| TB preventive treatment | WHO policy for assessing Household Contacts (HHC) | Target population: HHC of individuals started on TB treatment<br>Coverage: 80% of child (<15 years old) HHC receive TPT and 80% of all HHC assessed for TB | TPT: 60% reduction in progression to disease among recently exposed children<br>HHC screening: 3% of HHC have TB disease; these are identified and treated | US\$24.00-37.00 for HHC assessment.<br>US\$22.00-84.00 for TPT for DS-TB.<br>US\$41.00-149.50 for TPT for DR-TB. | <sup>26,27</sup> WHO consolidated guidelines TPT 2024, Fox 2013 ERJ |
| SCREENING<br>and<br>DIAGNOSIS |  |  |  |  |  |
| Community-wide screening | Annual symptom-agnostic<br>community-wide<br>screening using<br>chest X-ray<br>followed by Xpert<br>Ultra, if positive | Target population: all adults ($\geq 15$ years<br>old)<br>Coverage: scale-up to 80% 2025-2030, five annual rounds covering 80%<br>of adults, complete stop after 2035. | Early diagnosis of<br>bacteriologically<br>confirmed TB, increased<br>DST coverage. Lower<br>on-treatment mortality<br>due to earlier treatment<br>initiation | US\$41.50 for CXR<br>screening (high-cost<br>estimate) or<br>US\$2.50-5.50 for<br>CXR screening (low-cost estimate).<br>US\$17.00-24.50 for<br>Xpert Ultra. | <sup>12,28,29</sup> Marks<br>2019 NEJM<br>(ACT3),<br>Schwalb 2024<br>PGPH, Veeken<br>2025 JID |
| Improved<br>diagnostic<br>approach in<br>clinic | Xpert Ultra as part<br>of TB diagnostic<br>pathway in clinic,<br>replacing<br>CXR/empirical<br>diagnosis | Target population: individuals (all<br>ages) with presumptive TB entering<br>the TB diagnostic pathway in clinics<br>Coverage: 80% | Increased sensitivity for<br>bacteriologically<br>confirmed TB<br>Increased specificity for<br>non-disease states<br>Increased DST coverage | US\$24.00-37.00 (vs.<br>US\$13.50-31.00 in<br>BAU) | <sup>6,30</sup> Zifodya<br>2021 Cochrane<br>Review<br>WHO<br>consolidated<br>guidelines on<br>tuberculosis:<br>rapid<br>diagnostics for tuberculosis detection. Third edition |
| Drug susceptibility testing (DST) | Xpert Ultra for all diagnosed with TB in clinics to diagnose Rifampicin resistance | Target population: individuals diagnosed with TB in clinics receive an Xpert Ultra.<br>Coverage: 80% | Increased initiation of appropriate Rifampicin-resistant treatment<br>Reduced on-treatment mortality for DR-TB due to earlier initiation | US\$17.00-24.50 | <sup>6</sup> Zifodya 2021 Cochrane Review |
| TREATMENT |  |  |  |  |  |
| Shorter treatment for Drug Susceptible (DS) TB | WHO recommended 4-month treatment for DS-TB | Target population: individuals initiating DS-TB treatment<br>Coverage: 80% | Treatment duration shortened from 6 to 4 months<br>Non-inferior treatment success | US\$199.00-276.00 (vs. US\$119.50-227.50 in BAU) | <sup>7</sup> Dorman 2021 NEJM |
| Shortened treatment for Drug Resistant | WHO recommended 6-month treatment | Target population: individuals initiating treatment for Rifampicin-resistant TB | Treatment duration shortened to 6 months | US\$394.50-502.50 (vs. US\$450.50-3,600.50 in BAU) | <sup>8,31,32</sup> Nyang'wa 2023 Lancet |
| (DR) TB | for Rifampicin-resistant TB | Coverage: 80% | Treatment success increased to 89%, including reduced on-treatment mortality |  | RM James<br>2024 PMED<br>Sweeney 2022<br>PGPH |
| STRUCTURAL DRIVER |  |  |  |  |  |
| Incarceration (Brazil only) | Annual symptom-agnostic screening for all incarcerated individuals using Xpert Ultra | Target population: currently incarcerated individuals<br>Coverage: 80% | Early diagnosis of bacteriologically confirmed TB<br>Increased DST coverage.<br>Lower on-treatment mortality due to earlier treatment initiation | US\$21.50 | <sup>11</sup> Pivetta 2024<br>CID |
| Undernutrition (India only) | Nutritional supplementation for TB patients and their household contacts (RATIONS study) | Target population: all individuals initiating TB treatment and their HHC<br>Coverage: 80% | TB Patient: 33% reduced risk of on-treatment mortality<br>HHC: 39% reduced risk of progression to disease | US\$16.50 for TB patients. US\$6.00 for HHCs. | <sup>10,21</sup> Bhargava<br>2023 LGH<br>McQuaid 2025<br>LGH |

Scale-up and coverage scenarios are illustrated in supp mat S4, section 1. Intervention coverage was scaled up between 2025-2030 and then kept constant. For TB vaccines, we implemented routine vaccination of adolescents 15-19 years old and two mass catch-up campaigns (at scale-up and in 2040). ^20^ Mass community-wide screening would not be maintained for 25 years, as historical and contemporary data suggest impact will reduce over time. ^12,29^ Instead, we assumed a five-year scale-up to 80%, followed by five annual campaigns, after which screening would stop.

### Epidemiological Model

To estimate the potential epidemiological impact of all selected interventions, we developed a dynamic transmission model, extending an existing modelling infrastructure used to explore TB vaccine impact ^20^ with previously published structure and parameters on TB natural history. ^33^ The model was age-stratified, with differential TB natural history parameters on progression, diagnosis, infectivity and mortality between children (<15 years old) and adults (see supp material section S1, section 2.3).

The model core natural history structure and subdivisions are shown in Figure 1 and described in detail with parameterisation in supp mat S1. We recognise three disease states, aligning with recent re-categorisation by the World Health Organisation (WHO). ^23,34^ In brief, the model captures two bacteriologically positive infectious states, differentiated by symptomatic and asymptomatic TB (sTB and aTB), which contribute to transmission (i.e. can be infectious). Bacteriologically negative non-infectious TB (nTB) does not contribute to transmission, and includes individuals who have TB pathology that may be recognised by imaging, e.g. chest X-ray (CXR), but lack bacteriological evidence of TB, although a proportion will progress to bacteriologically confirmed TB. ^35^ In addition, the model recognises self-clearance of *Mycobacterium tuberculosis* (*Mtb*) infection, as well as regression and recovery from disease states. Extrapulmonary TB was not modelled explicitly, but included within the non-infectious TB state. The model was further subdivided to include treatment and infection with drug-susceptible or rifampicin-resistant strains.

**FIGURE 1:**
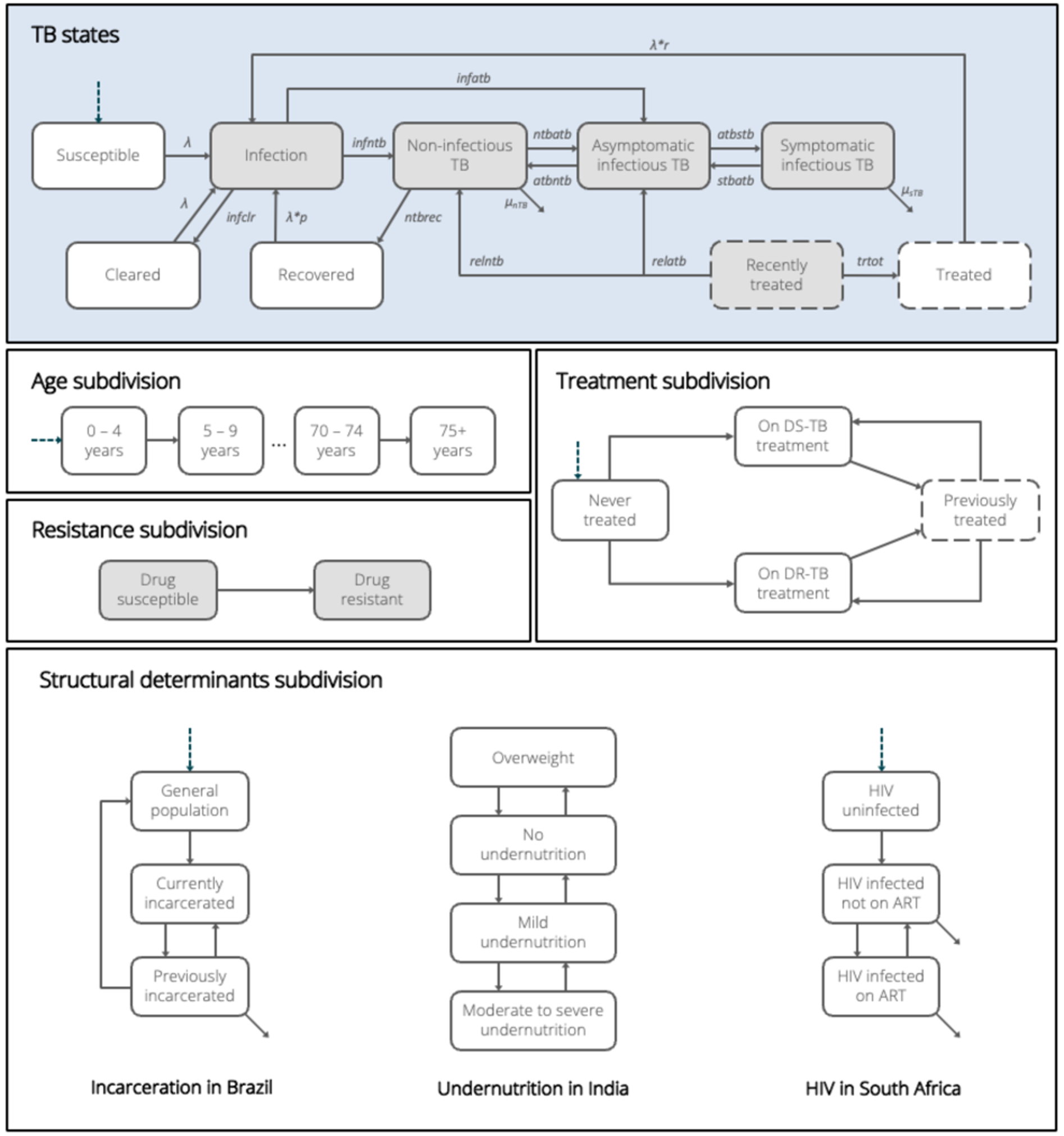
Epidemiological model structure and subdivisions. Model structure for the natural history of TB, incorporating both general subdivisions and country-specific structural determinants: incarceration (Brazil), undernutrition (India), and HIV (South Africa). Background mortality is assumed for all states but not shown; arrows exiting from the corners of certain states represent specific mortality rates. The model is age-stratified in 5-year bands up to 75+ years. Compartments shaded in grey indicate states in which individuals harbour viable Mycobacterium tuberculosis, further subdivided by drug resistance. Compartments with dashed lines indicate states which appear only in the previously treated stratum within the treatment subdivision.

We implemented diagnostic pathways for individuals undergoing screening and diagnosis in clinic- and community-based settings. We explicitly considered individuals without TB entering the diagnostic pathways, with test performance (expressed as test positivity) specified across the different model states. See supp mat S1, table S3 in section 1.3 for values and data sources.

Structural drivers were included to capture the key population differences by epidemiological risk and intervention impact, including incarceration (not incarcerated, currently incarcerated, recently incarcerated, i.e., individuals who have been released from prison within the past 7 years) in Brazil, nutritional status (Body Mass Index (BMI) of <17kg/m^2^, 17-18.4kg/m^2^, 18.5-24.9kg/m^2^, >25kg/m^2^ (adult thresholds)) in India, and HIV (HIV-negative, HIV-positive, HIV-positive and receiving ART) in South Africa. Data on subgroup sizes and trends are described in detail in sup mat S1 section 4-6. Parameterisation of natural history was based on review of published data and modelling. ^5,36–38^

Heterogeneous mixing was implemented by 5-year age groups until 75+. ^39^ In addition, we incorporated assortative mixing by incarceration status, and strongly increased mixing between incarcerated individuals to reflect known issues of overcrowding (**supp mat S1, section 6.5**). ^40^

### Economic model

To estimate budgetary impact, incremental costs, and cost-effectiveness, we constructed a costing model which linked the intervention activities and epidemiological outcomes to costs. This included costs for existing TB diagnosis and treatment services, for which demand was altered by the interventions, as well as costs of providing each intervention, over 2025-2050. See supp mat S3 for details of the economic model.

Costs were sourced for each country based on previous studies of these interventions or publicly available data sources (see supp mat S3, section 3.2 for details). ^10,20,41–46^ We used a health system perspective, thus providing results relevant to public health funders. All costs are expressed in 2023 USD. We used a standard 5% annual discounting rate for costs and benefits across all countries.^47^ For community-wide screening, we used both high- and low-cost scenarios for the diagnostic tools due to a wide discrepancy in the estimated diagnostic costs between countries (Table E3 in S4). ^42,44,45^ To capture uncertainty in costs, we generated a gamma distribution around each point estimate using a conservative range of +/-50%, from which we sampled 500 cost sets for each country. Each cost set was randomly matched to a parameter set from the epidemiological model to cost activities.

Estimated cost-effectiveness threshold ranges from Ochalek et al. 2018 were adopted and updated to 2023 GDP values (see supp mat S3, section 2.6). ^48^

### Model calibration

For each epidemiological archetype, we calibrated the model to trends in TB incidence, notifications, and mortality in 2010 and 2023 as reported by the WHO. In addition, we calibrated to prevalence and proportion of asymptomatic TB where available (India and South Africa). We also calibrated to indicators for structural drivers, including incarceration rate and notification rate among incarcerated individuals (Brazil), population attributable fraction for BMI under 18.5kg/m^2^ (India), and TB incidence, notification, and mortality among HIV-positive individuals (South Africa). A full list of calibration targets, data sources and methods is provided in sup matt S1, section 7.

Calibration was done by seeding an Approximate Bayesian Computation using Markov chain Monte-Carlo (ABC-MCMC) sampling with variable step size, with a manual fit to all calibration targets. After ABC-MCMC identified at least 25,000 further parameter sets consistent with all calibration targets, we selected 500 at random.

Models were calibrated up to 2025, after which we generated the business-as-usual (BAU) comparator scenario, by keeping all parameters constant from 2025, except for projections of HIV and ART prevalence in South Africa (based on UNAIDS projections) and changes in BMI distribution in India (based on Alexander et al. 2023 ^49^). The BAU scenario was used to compare against interventions, each of which was run across all 500 parameter sets per epidemiological archetype, providing paired estimates of epidemiological impact.

### Model outcomes

For each intervention, we generated trends in infectious TB prevalence (aTB+sTB) to match current disease measurement in prevalence surveys, as well as incident sTB episodes, TB deaths, and disability-adjusted life years (DALYs) averted compared to BAU up to 2050.

DALYs were calculated based on published disability weights for sTB ^50^ and life years lost due to TB deaths, and included post-TB DALYs incurred after treatment or recovery from sTB (in absence of DALY estimates available for aTB). ^51^ See supp mat S3, section 4 for more details. We calculated budgetary impact as total costs for each intervention over the 2025-2050 period, as well as incremental costs and incremental cost-effectiveness ratios (ICERs), expressed as cost per DALY averted relative to BAU. We generated median estimates for results, with corresponding 95% uncertainty intervals (UI), calculated as the 2.5th to 97.5th percentiles of the parameter sets, to quantify uncertainty.

### Sensitivity analyses

We explored the epidemiological impact of between-country variation in parameterisation of natural history by assuming the same natural history parameters across countries (disregarding calibration targets).

### Data availability

Data and analysis scripts are available at https://github.com/lshtm-tbmg/PACE-TB.

## Results

### Calibration and baseline

All 500 selected parameter sets fitted to calibration targets, with a detailed overview of posterior parameter ranges and calibration targets, are provided in Supp mat S4, section 1. Figure 2 shows projected incidence trends under BAU up to 2050 (black dashed line) for each of the archetype countries, with different baseline burden and projected trends up to 2050.

**FIGURE 2:**
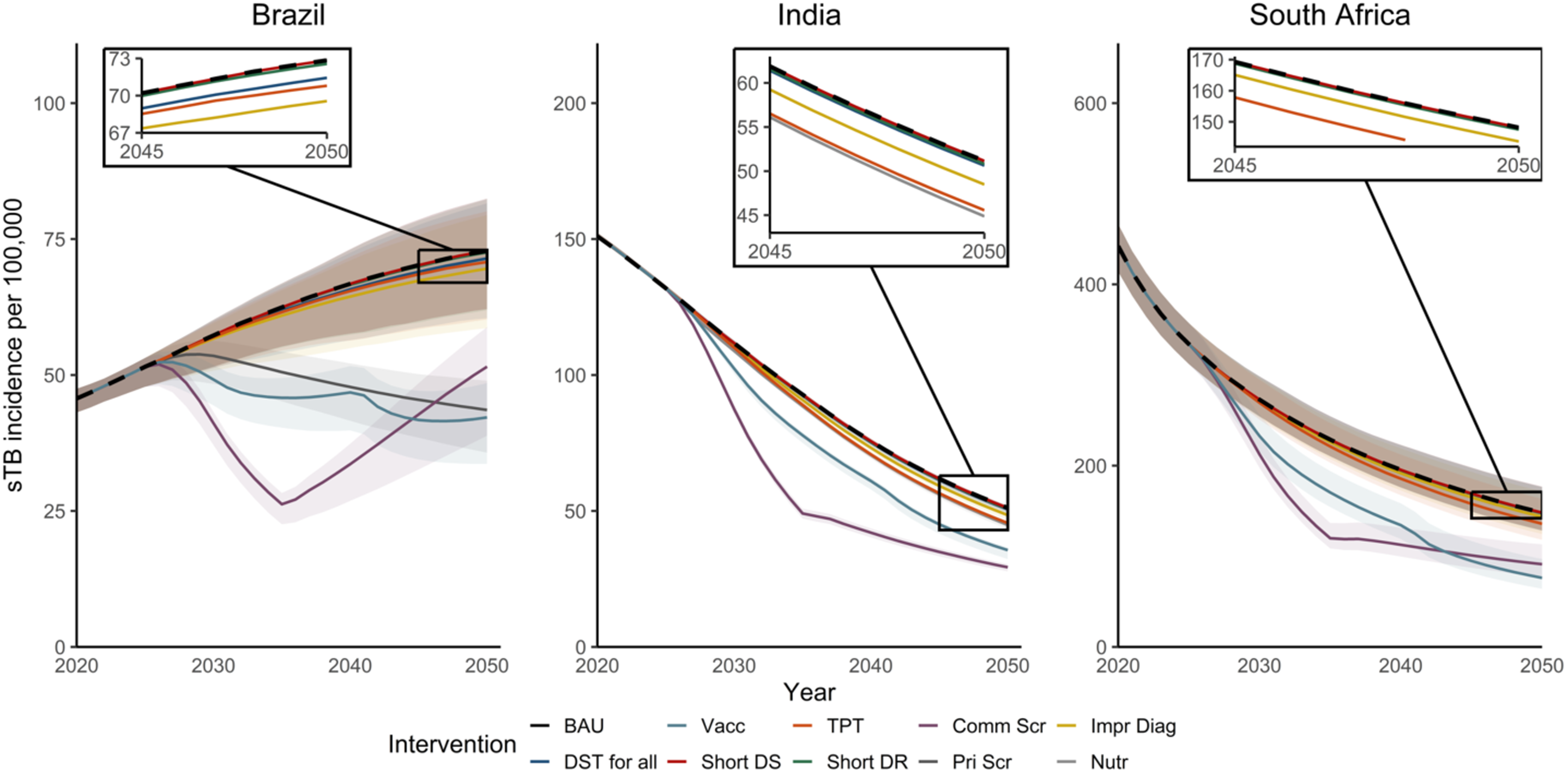
Incidence trends for Business as Usual and interventions. BAU = Business-As-Usual, Vacc = Vaccination, TPT = Tuberculosis Preventive Treatment, Comm Scr = Community-wide screening, Impr Diag = Improved diagnosis in clinics, DST for all = Drug Susceptibility Testing for all clinic-diagnosed individuals, Short DS = Shortened treatment for Drug Susceptible TB, Short DR = Shortened treatment for Drug Resistant TB, Pri Scr = Mass screening in incarcerated individuals, Nutr = Nutritional support for households of individuals receiving TB treatment. Lines show BAU incidence trends (black dashed line) and intervention trends in incidence for each country. Shading reflects 95% Uncertainty Intervals (UI). Note UIs overlap for most interventions. Error bars show 2023 incidence calibration target. Pop-outs show last five years for incidence curves (without uncertainty) that lie close to the baseline (black dashes line).

### Epidemiological impact

The coloured lines in Figure 2 show the estimated impact of the interventions, with relatively consistent patterns between countries for interventions as compared to BAU. Most trajectories remain close to BAU, indicating limited estimated population-level impact. Exceptions are vaccination (green line) and community-wide screening (purple line) as well as annual systematic screening in prisons in Brazil (grey line).

Trajectories for community-wide screening reflect the stop of activities in 2035. Thereafter, incidence slopes start to revert to BAU in each setting, reducing the difference with BAU in 2050. For vaccination, the catch-up campaign in 2040 is reflected in the change in trajectory. Trends in TB prevalence and mortality show similar trends (see supp mat S4, section 2).

When considering the difference between incidence trends in BAU and interventions we estimate the proportion of sTB episodes averted between 2025-2050 (Figure 3, supp matt S4, section 2). We found that three interventions showed a high impact (>10% averted): vaccination (median 15-28% of sTB episodes averted across countries), community-wide screening (31-38%) and screening in prisons in Brazil (23%). Other interventions, which targeted individuals accessing care through clinics (improved diagnostic approach in clinics, increased access to DST, shortened treatment for DS- and DR-TB) or their households (TPT, nutritional support) were each estimated to avert fewer than 5% of total episodes in the population. For these interventions, median sTB episodes averted ranged between 0% (shortened treatment for DS-TB) to 5% (nutritional support in India).

**FIGURE 3:**
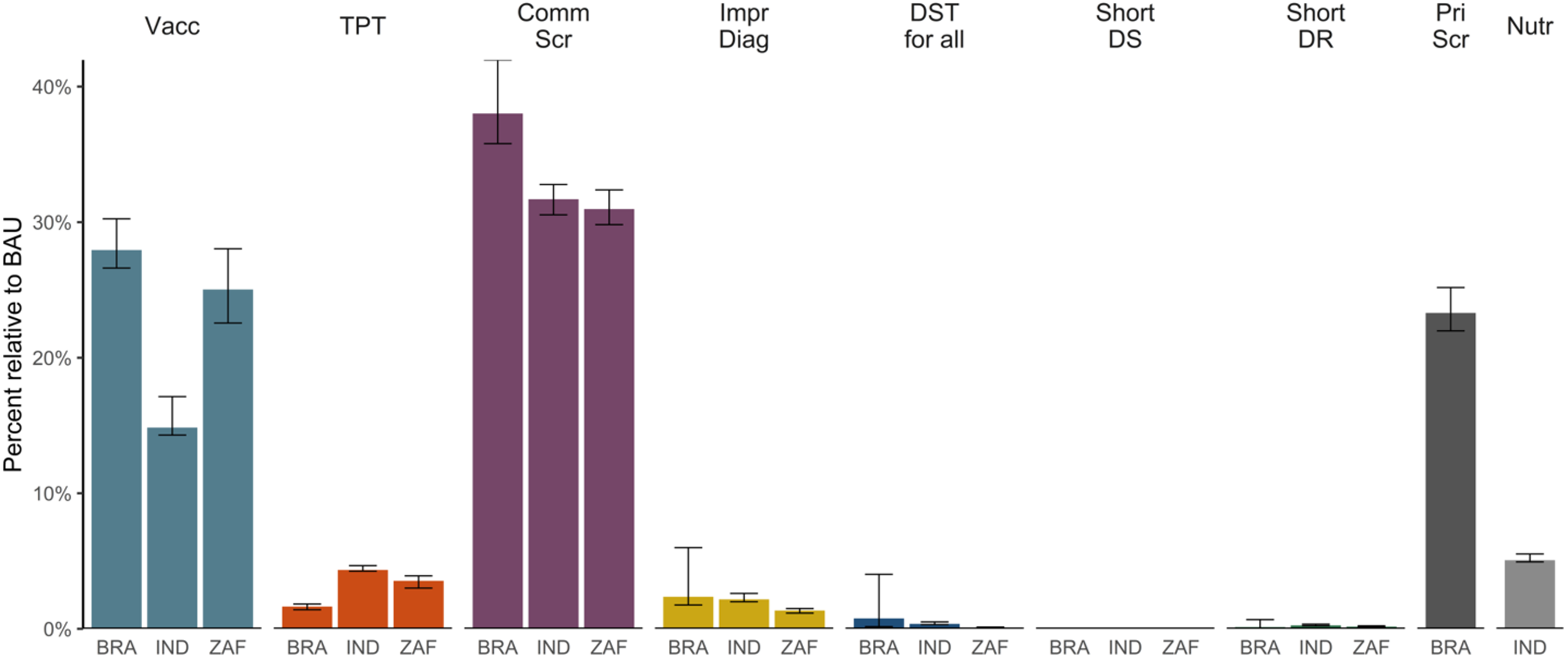
Percentage of incident symptomatic TB episodes averted (2025-2050) Vacc = Vaccination, TPT = Tuberculosis Preventive Treatment, Comm Scr = Community-wide screening, Impr Diag = Improved diagnosis in clinics, DST for all = Drug Susceptibility Testing for all clinic-diagnosed individuals, Short DS = Shortened treatment for Drug Susceptible TB, Short DR = Shortened treatment for Drug Resistant TB, Pri Scr = Mass screening in incarcerated individuals, Nutr = Nutritional support for households of individuals receiving TB treatment. BRA = Brazil, IND = India, ZAF = South Africa. Error bars reflect 95% Uncertainty Intervals.

Proportional reduction in prevalence in 2050 shows a similar pattern (see supp mat S4, section 2.5), although the overall impact was more similar between the vaccination, community-wide and prison screening interventions (median 30-52% reductions), in part driven by the rebound in prevalence following cessation of community-wide screening in 2035.

### Budget impact and incremental costs

Total budget impact (in USD and local currencies) and incremental costs for the interventions are provided in the supplementary table (supp mat S4, section 3). Differences between settings were driven in large part by the interaction between population size and TB burden, which set the absolute size of the target populations.

Incremental costs, compared to the costs for BAU, varied widely between cost-saving (-0.3bn, -0.3bn, -0.04bn USD for shortened DR-TB treatment in Brazil, India and South Africa respectively) to over 240 billion USD (high-cost community-wide screening in India). Figure 4 shows incremental costs for each intervention against DALYs averted.

**FIGURE 4:**
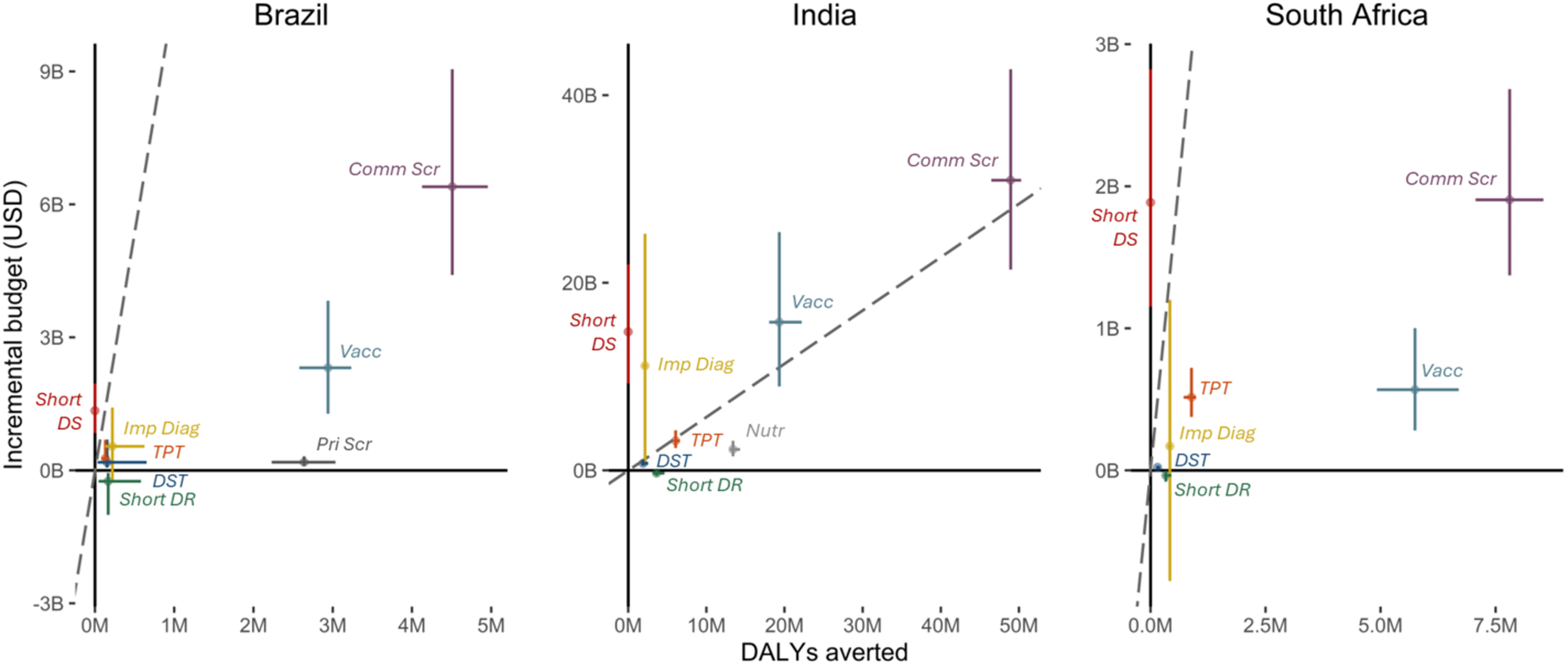
Incremental Cost-Effectiveness Ratios for interventions plots. Error bars show 95% uncertainty intervals for DALYs averted (horizontal lines) and incremental budget (vertical lines). Dashed line shows cost-effectiveness thresholds. Vacc = Vaccination, TPT = Tuberculosis Preventive Treatment, Comm Scr = Community-wide screening (low diagnostic cost), Impr Diag = Improved diagnosis in clinics, DST for all = Drug Susceptibility Testing for all clinic-diagnosed individuals, Short DS = Shortened treatment for Drug Susceptible TB, Short DR = Shortened treatment for Drug Resistant TB, Pri Scr = Mass screening in incarcerated individuals, Nutr = Nutritional support for households of individuals receiving TB treatment. Note, see supp mat S4, section 3 for community-wide screening with high cost.

Implementing prison-based screening cost only 0.19bn more than the 1.6bn for BAU, due to savings in TB treatment costs. In contrast, incremental costs in South Africa for replacing current standard DS-TB treatment with a shorter regimen (1.9bn) were above that of TB vaccination (0.5bn) due to differences in population impact and the associated cost-savings. Incremental costs for vaccination and community-wide screening were amongst the highest in each country, although the low-cost scenario for community-wide screening reduced the incremental budget by 63-81% compared to the high-cost scenario across all settings (Supp mat S4, section 3).

### Cost-effectiveness

We present the incremental costs and DALYs averted for each intervention compared to BAU in Figure 4, and provide the ICERs in Supp mat S4 section 3. We found that only shortened treatment for DR-TB would be cost-saving (negative incremental cost). With no epidemiological impact for shorter DS-TB treatments (following non-inferiority in trial findings ^7^), this intervention was above the estimated cost-effectiveness threshold in all countries (Figure 4 - dashed grey line).

Both interventions targeting key structural drivers had very low estimated ICERs, with 72 USD/DALY for prison screening in Brazil and 167 USD/DALY for nutritional support in India.

In Brazil, the ICERs fell below the estimated cost-effectiveness threshold range (6,968-10,735 USD/DALY) and would be cost-effective compared to BAU for all interventions apart from high-cost community-wide screening (ICER of 8,174 USD/DALY, 31-87% probability of being cost-effective) and shorter DS-TB treatment. In India, with a threshold range of 413-569 USD/DALY, shortened DR-TB treatment was cost-saving, nutrition had an ICER of 167 USD/DALY (100% probability cost-effective) and DST for all had an ICER of 390 USD/DALY (60-94% probability cost-effective), with all other interventions having ICERs above the threshold range and a 0-25% probability of cost-effectiveness compared to BAU. In South Africa, all interventions other than shorter DS-TB treatment produced ICERs well below the threshold (2,444-3,285 USD/DALY), with 93-100% probability of being cost-effective compared to BAU. Between countries, the ICERs for low-cost community-wide screening changed in line with baseline prevalence of infectious TB (1,437, 639 and 217 USD/DALY for Brazil, India and South Africa respectively).

### Sensitivity analyses

After alignment of core natural history parameters, the population impact for vaccination was similar between India and South Africa.

## Discussion

### Key findings

Our comparison of TB interventions found that only community-wide screening, vaccination, or annual screening in prisons in Brazil, prevented more than 10% of incident TB episodes in any setting. Other interventions which targeted a smaller population (e.g. TPT among HHCs or shorter DS-TB regimen for those diagnosed in clinics) prevented fewer than 5% of incident episodes. Cost-effectiveness ratios ranged widely between interventions, but some interventions with high absolute costs activities (vaccination, community-wide screening) provided value for money in some settings (Brazil and South Africa, but not India). Results were reasonably consistent across settings.

### Main messages

The results highlight how potential population-level impact is largely driven by the combination of size and burden of the target population, often summarised as the PAF. ^52^ If an intervention is restricted to clinic-diagnosed individuals or their household contacts, the target population size puts a hard ceiling on the potential population-level benefits, even if the within-group burden is high. If interventions prevent transmission either through early diagnosis (community-wide or prison screening) or preventing progression to disease (prevention of disease vaccine), they can provide substantial epidemiological impact. Our results also show the importance and value for money of addressing structural drivers of TB, with the examples showing promise similar to the known benefits of ART roll-out in South Africa. ^53,54^ Considering costs as part of evaluating interventions showed the benefits of reducing regimen costs (shortened DR-TB treatment) or diagnostic algorithms costs (low-cost community-wide screening). Conversely, if an intervention with limited epidemiological benefit (e.g. non-inferior shorter DS-TB treatment) also has a higher cost, it will struggle to be cost-effective.

Our country archetypes provide contrasting future projections under BAU, driven mostly by persistence or continued reduction of impact from the key structural drivers: increasing incarceration trends in Brazil, continued high coverage of ART in South Africa and reductions in undernutrition in India. While future trends in structural drivers and their impact are highly uncertain in the current global funding landscape ^2,20^, the relative consistency of findings across different background trajectories suggest our findings are robust.

### Contrast with previous findings

Our results update previous comparisons across intervention domains. ^16,17^ Some findings align, including the importance of addressing disease and transmission on a population level. ^17^ A key contrast is the projected population-level impact of new shortened treatment regimens, in particular for DS-TB, which are mainly driven by assumed improved treatment outcomes, while empirical evidence has only shown non-inferiority. ^13,16^ The low cost and high microbiological success of current DS-TB regimens in trial settings (∼95% treatment success in the standard of care group ^55^) give limited space for assuming or acquiring empirical evidence for improvements or cost savings from new regimens. For TPT, our population-level epidemiological impact of HHC-targeted TPT estimates were similar to others ^56,57^, whereas we did not model mass-TPT to complete populations. ^17^

### Limitations

As with any modelling exercise, there are limitations to our work. Our choice to compare interventions with empirical evidence for impact could constrain the potential of certain approaches and ignores the potential gap between intervention effectiveness in trial and operational settings. ^58^ However, going beyond current evidence would risk subjective imbalances in assumed potential efficacy between intervention categories, and would limit a more objective comparison of potential population-level health benefits and costs.

We did not consider operational limitations to achieving 80% coverage, which many interventions have not yet achieved outside of research settings, ^12,59^ nor did we consider differential costs that may be required to overcome operational challenges for each intervention when scaling up from . Rather than ignoring operational challenges, our results highlight the relative potential of interventions and the value of resolving logistical challenges to unlock the potential epidemiological benefits of an intervention or reduce costs.

Countries included in our analysis represent lower-middle income (India) and upper-middle income (South Africa and Brazil) countries. Cost-effectiveness interpretations may differ in low-income settings.

While we used local costs where possible, some health system costs may have been omitted. As such, absolute values for budget impact should be interpreted with caution. However, the level of detail should be comparable between intervention categories, enabling comparisons. We also did not consider patient costs, which limited quantification of potential cost benefit for individual patients rather than society. However, cost and DALY savings will always be dominated by prevention of new incident TB episodes, which was our main outcome, rather than savings on management of existing TB, suggesting our broad results are robust to this.

Our exercise only considered each intervention compared to BAU and did not explore independent improvements in BAU, such as increased clinic attendance or other changes in structural determinants of TB (e.g., long-acting antiretrovirals for HIV in South Africa). We also did not model intervention combinations where potential synergies are quantified, e.g. the benefit of shorter DS-TB treatments in the context of community screening, or a staggered introduction of community-wide screening followed by vaccination, which may have shown additional benefit for one or more of the interventions.

### Big takeaways

Despite these limitations, our work provides a much-needed perspective on the potential impact of intervention domains across the TB response portfolio. We show that interventions need to reach populations with substantial contributions to TB burden (as quantified in PAF) to make progress on prevention of new TB episodes and mortality. This point has been made before ^60–62^, but our results provide a stark picture of the continued morbidity and mortality we implicitly accept if we fail to move our interventions beyond current TB patients and their household contacts. Our incremental budget results highlight that this requires a different level of ambition in terms of funding, but also that such investments provide likely value for money as well as meaningful population-level impact.

Community-wide screening was highly impactful, but also costly, and the ICER increased with decreasing burden. While below the cost-effectiveness threshold in high burden populations, community-wide screening in settings like Brazil, where estimated incidence sits at or below 50/100,000, is unlikely to be the most efficient approach. Here, a combination of lower cost, high impact interventions, community-wide vaccination and screening targeted at a high-risk population (e.g. incarcerated individuals) may be more appropriate. ^11,63^ For higher burden settings, identifying cost-savings for screening approaches is key, e.g. simplifying algorithms to a single low-cost single bacteriological test with performance in line with current PCR-tools or identifying more efficient symptom-agnostic screening approaches. ^28,64^ In addition, linking such efforts with vaccination campaigns (either in parallel or sequential) could ensure impacts are sustained after cessation of community-wide screening at a set burden threshold. Such multi-faceted approaches, combining vaccination, mass-screening and treatment of multiple disease phenotypes, have been shown to have an impact historically in high burden settings, most notably among First Nation populations in Alaska during the mid-20th century. ^61,65^

Our work shows the value of a cross-silo approach when considering priorities for research and funding. In addition, development of TPPs should look beyond the benefit for individuals or within the limited target population, and also consider the desired population-level impact and what improvement over current performance is needed to achieve that impact. That should include how existing interventions can or should be adapted for performance in recently defined TB phenotypes and outcomes such as aTB among community-diagnosed individuals, or the risk of post-TB lung disease, which is a major driver of TB-related morbidity. ^51^

### Conclusion

Interventions with meaningful impact can also be cost-effective, but achieving their potential may require a priority shift in public health funding, policy and product development.

## Data Availability

All information, data and code to replicate these analyses are contained in the supplementary materials and the GitHub repository

https://github.com/lshtm-tbmg/NIH2

## Acknowledgements

We are grateful to Daniele Maria Pelissari, Fernanda Dockhorn Costa Johansen, Eduardo de Souza Alves for discussions around identifying epidemiological and cost data for Brazil. We also thank members of the PACE-TB advisory group (Nimalan Arinaminpathy, Sandip Mandal, Emily Kendall and Nick Menzies) for advice and review during model development and analysis.

