## Supplementary materials 1 (S1) for "The potential impact, cost and cost-effectiveness of tuberculosis interventions - a modelling exercise"

### SUPPLEMENTARY MATERIAL - S1 - MODEL STRUCTURE:
### The potential impact, cost and cost-effectiveness of tuberculosis interventions - a modelling exercise

Katherine C. Horton [1 2], Alvaro Schwalb [1 2 3], Martin J. Harker [1 2 4], Lara Goscé [1 2], Elena Venero-Garcia [1 2], Lily O'Brien [1 2], Arkaprabha Gun [1 2 5], Tom Sumner [1 2], C. Finn McQuaid [1 2], Rebecca A. Clark [1 2], Tomos O. Prys-Jones [1 2], Roel Bakker [1 2 6], Yiran E Liu [7 8], Mmamapudi Kubjane [9], Christian Lienhardt [10 11], Richard G. White [1 2], Rein M.G.J. Houben [1 2]

1 - TB Modelling Group, TB Centre, LSHTM, London, UK; 2 - Department of Infectious Disease Epidemiology, LSHTM, London, UK; 3 - Instituto de Medicina Tropical Alexander von Humboldt, Universidad Peruana Cayetano Heredia, Lima, Peru; 4 - Global Health Economics Centre, LSHTM, London, UK; 5 - Department of Epidemiology, Biostatistics, and Occupational Health, School of Population and Global Health, McGill University, Montreal, QC, Canada; 6 – KNCV Tuberculosis Foundation, The Hague, Netherlands; 7 - SEICHE Center for Health and Justice, Yale University School of Medicine, New Haven, CT, USA; 8 - Justice Collaboratory, Yale Law School, New Haven, CT, USA; 9 - Health Economics and Epidemiology Research Office, Wits Health Consortium, Johannesburg, South Africa; 10 - French Institute for Research in Sustainable Development (IRD), Montpellier, France; 11 - CRDF Global, Arlington, VA, USA

**Correspondence:** Rein M.G.J Houben, Department of Infectious Disease Epidemiology, London School of Hygiene and Tropical Medicine, Keppel St, London, WC1E 7HT United Kingdom

**GitHub:** https://github.com/lshtm-tbmg/PACE-TB

### MODEL STRUCTURE

#### 1. TB DIMENSION

This section aims to describe the TB dimension in TBMod. The TB dimension is the cornerstone of the structure encompassing the natural history of TB, while acknowledging treatment history and drug resistance. TBMod has been under development for ~7 years, and is an adaptation of TBVax informed by work on the natural history of TB (Clark, Mukandavire, et al., 2023; Horton et al., 2023; Portnoy, Arcand, et al., 2023; Portnoy, Clark, Quaife, et al., 2023; Portnoy, Clark, Weerasuriya, et al., 2023; Richards et al., 2023).

##### 1.1 OVERVIEW

- The TB dimension contains a total of 49 states describing natural history, diagnosis and treatment, and drug resistance (**Figure S1**). At its core, 25 states describe the natural history, with some states being replicated, as necessary, to reflect individuals never treated (_N) and those previously treated (_P), as well as individuals with drug-susceptible infection or disease (_s_) or drug-resistant infection or disease (_r_). A further 24 states describe individuals on treatment, accounting for the state from which an individual was diagnosed and whether an individual is placed on standard first-line treatment (ST_) or second-line treatment for drug resistant TB (RT_).
- As an overview of **Figure S1**, individuals never treated (_N) can be placed on treatment and after completion will move on to become individuals previously treated (_P), which can be placed on treatment once again.
- Further details on structure are shown in the next section. Components of the model structure - natural history, diagnosis and treatment, and drug resistance - are discussed in subsequent sections with simplified model structure diagrams.

#### 1.2 STRUCTURE

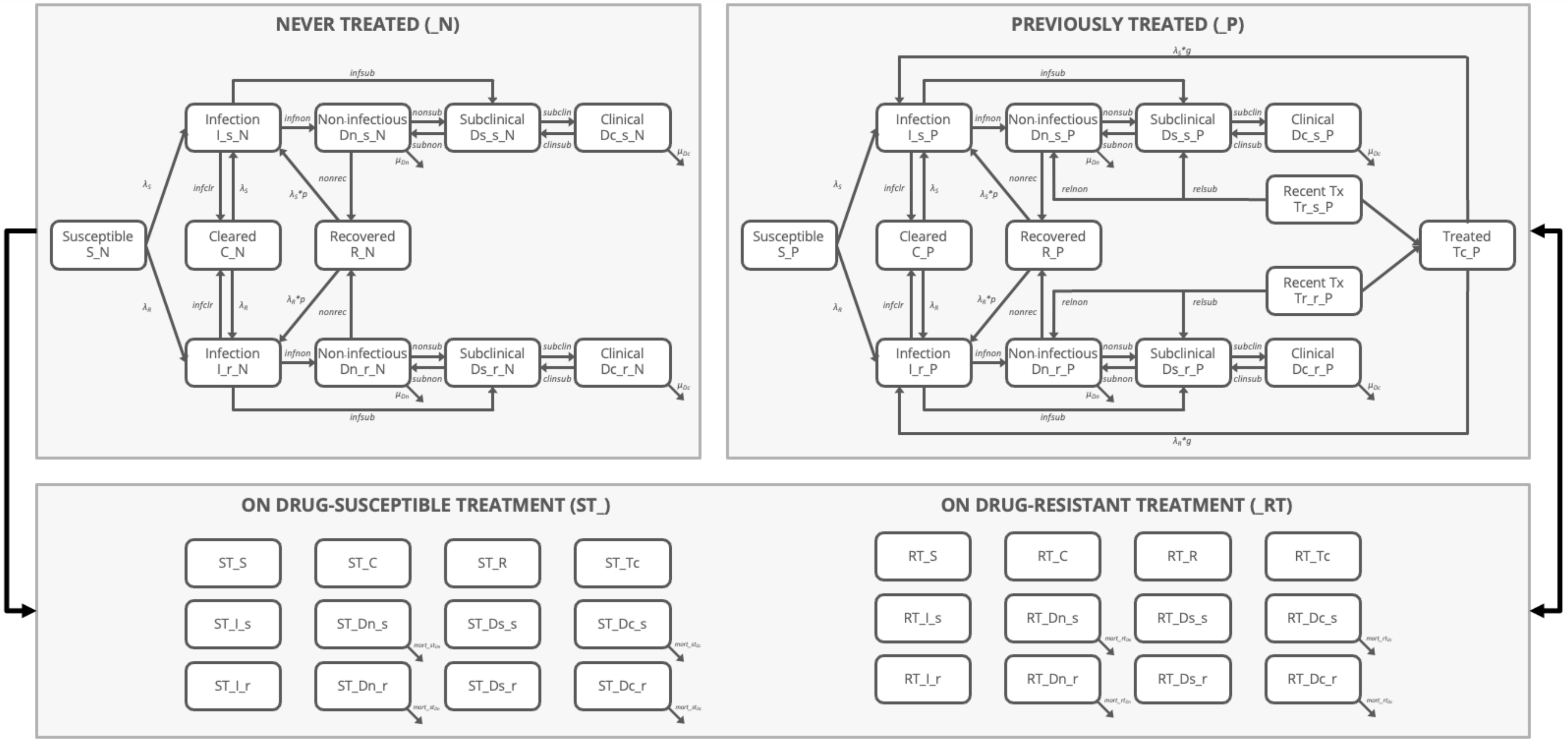

Figure S1. TB dimension structure. TB disease state and parameter naming follow conventions from research over the past decade; results are reported using current WHO terminology.

##### 1.2.1 NATURAL HISTORY

This section aims to describe in detail the TB natural history structure, without considerations of treatment history or strain susceptibility.

**Figure S2** shows 9 states that exemplify the essence of the TB natural history. Disease state classification was informed by the ICE-TB framework (Coussens et al., 2024) , and naming follows current WHO definitions (Falzon et al., 2024).

- Susceptible individuals (S) are infected at rate $\lambda$ and progress to the infected compartment. Infected individuals (I) may clear infection (C) at rate *infclr* or progress to non-infectious disease (Dn) or asymptomatic disease (Ds) at rates *infnon* and *infsub*, respectively.
- Individuals with non-infectious disease (Dn) recover (R) at rate *nonrec* or progress to asymptomatic disease (Ds) at rate *nonsub* or die from TB-associated mortality at rate $\mu_{Dn}$. Individuals with asymptomatic disease (Ds) regress to non-infectious disease (Dn) at rate *subnon* or progress to symptomatic disease (Dc) at rate *subclin*. Individuals with symptomatic disease (Dc) regress to asymptomatic disease (Ds) at rate *clinsub* or die from TB-associated mortality at rate $\mu_{Dc}$.
- Diagnosis and treatment are described in detail in the following section. Recently treated individuals (Tr) may relapse to non-infectious disease (Dn) or asymptomatic disease (Ds) at rates *relnon* and *relsub*, respectively. Furthemore, recently treated individuals (Tr) progress to Treated (Tc). Individuals who have recovered from non-infectious disease (R) and those who have been treated for infectious disease (Tc) have a different risk of reinfection expressed through a relative risk factor *p* (protection after recovery) and *g* (greater risk after treatment)

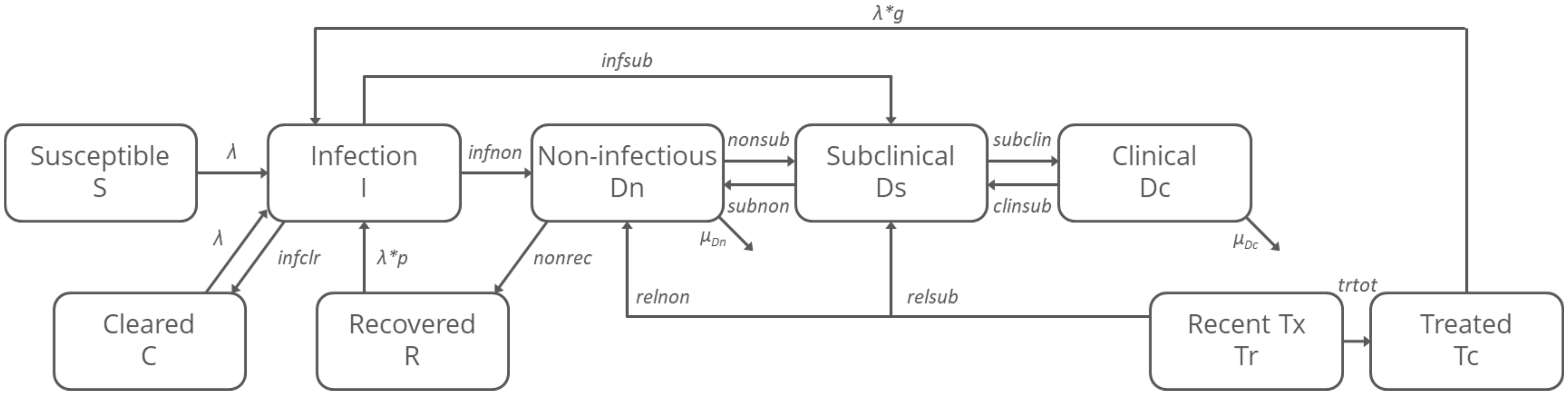

Figure S2. Natural history structure within TB dimension. TB disease state and parameter naming follow conventions from research over the past decade; results are reported using current WHO terminology.

Force of infection ($\lambda_{str}$)

The formula calculates the force of infection ($\lambda_{str}$) for a given strain of *Mycobacterium tuberculosis* (*Mtb*) by considering the probability of transmission per contact (*pT*), a scaling factor for contacts (*cscal*), and the sum of age-specific contact rates (*C[m,y]*). Additionally, it incorporates the number of individuals with symptomatic (*TDc*) and asymptomatic TB (*TDs*) (accounting for the relative infectiousness *t* of asymptomatic disease), the relative infectiousness of the strain (*relinf*), the relative infectiousness of children compared to adults (*k*), and the total population in each age group (*Ny*).

Susceptible individuals (S) are infected at rate $\lambda_S$ or $\lambda_R$. $\lambda$ is defined as follows:

$$\lambda_{str} = pT * cscal * \sum_{y=1}^{n_{ygroups}} C[m,y] * \left( \frac{\left( TDc_{str_y} + tTDs_{str_y} \right) * relinf_{str} * k}{N_y} \right)$$

where
- *str* indicates strain (drug susceptible or drug resistant)
- *pT* indicates probability of transmission per contact
- *cscal* indicates a multiplier to scale contacts to fit to burden data
- *y* indicates the age group of contact
- $n_{ygroups}$ indicates the number of contact age groups
- *C[m,y]* indicates the number of age-specific contacts. It is the element of the contact matrix ***C*** indicating the number of yearly contacts between individuals of age groups m and y.
- $TDc_{stry}$ indicates the total number of individuals with symptomatic TB of strain *str* in age group *y*
- *t* indicates the relative infectiousness of asymptomatic TB relative to symptomatic TB
- $TDs_{stry}$ indicates the total number of individuals with asymptomatic TB of strain *str* in age group *y*
- $relinf_{str}$ indicates the relative infectiousness of the strain
- *k* indicates the relative infectiousness of children (<15 years) relative to adults (≥15 years)
- *Ny* indicates the total population size in age group *y*

##### 1.2.2 DIAGNOSIS AND TREATMENT

This section aims to provide a detailed description of the transitions into and out of the treatment compartments. **Figure S3** shows the stratification of the TB natural history model described previously into two similar structures representing treatment history: never treated (_N) and previously treated (_P).

- Diagnosis and treatment are structured across two nearly parallel sub-strata, one for individuals never treated (_N) and one for individuals previously treated (_P). Note that Recently treated (Tr) and Treated (Tc) are only featured among those previously treated, as per definition.
- Individuals from any TB state (except those Recently treated (Tr)) may be placed on standard first-line treatment (ST_) or second-line treatment for drug-resistant TB (RT_).
- On-treatment states do not distinguish treatment history, i.e. individuals with asymptomatic TB (Ds) will be placed on either ST or RT treatment regardless of their treatment history.

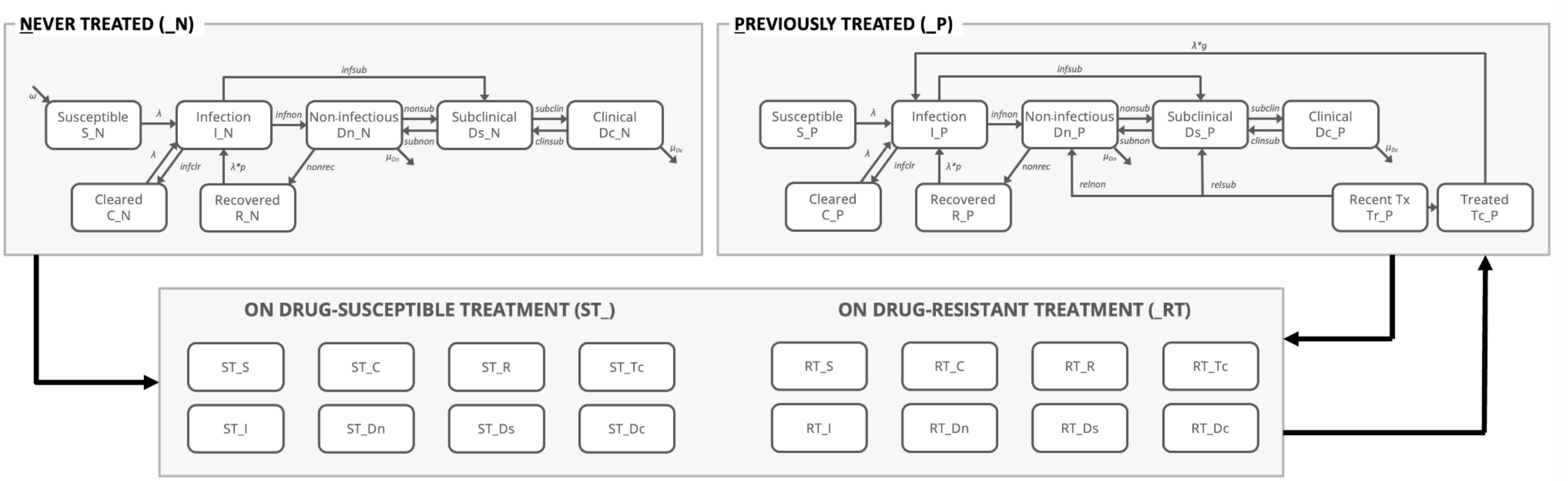

Figure S3. Diagnosis and treatment structure. TB disease state and parameter naming follow conventions from research over the past decade; results are reported using current WHO terminology.

**Figure S4** provides further detail on the flows of treatment initiations for each state. Boxes in light grey and dashed borders are duplicates (i.e. not repeated in each treatment history state), but shown here to help representation of state flows.

- There are separate on-treatment compartments for each TB state. Individuals initiate treatment standard treatment (ST_), following blue dashed lines, or drug-resistant treatment (RT_), following yellow dashed lines. Treatment initiation is determined by a treatment initiation rate (*tinit*), a country-specific factor that controls the overall treatment initiation rate (*eta*), and a relative factor (*relinit*), which varies across TB states.
- Initiation of drug-resistant treatment is defined by access to drug susceptibility testing (DST), which differs based on treatment history (*dst_n, dst_p*) and probability of a positive DST result, which is determined by drug resistance status (*dst_s, dst_r*).
- Recently treated individuals (Tr_P) are not placed on treatment due to recent treatment history.

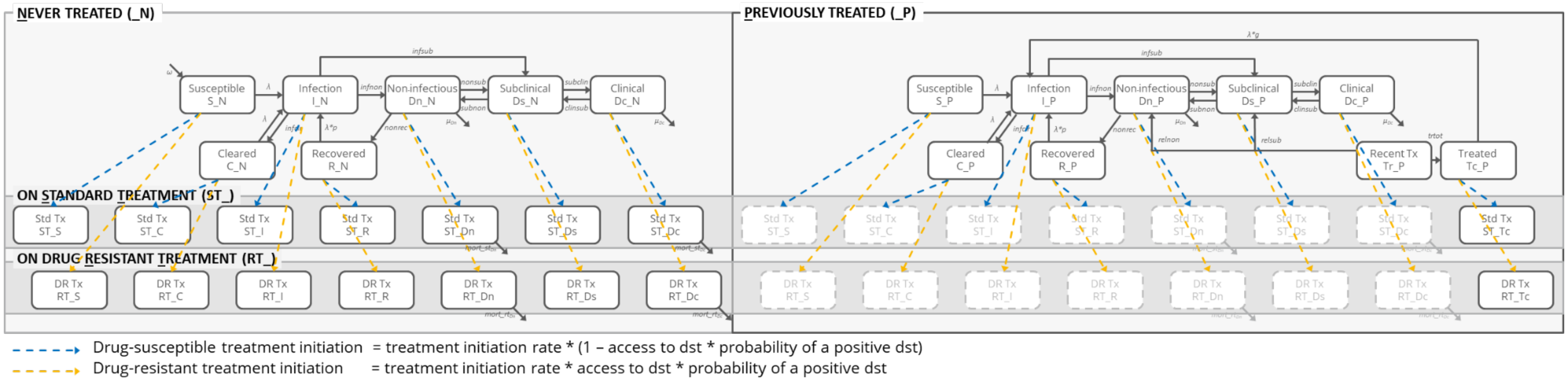

Figure S4. Treatment initiation in diagnosis and treatment structure. TB disease state and parameter naming follow conventions from research over the past decade; results are reported using current WHO terminology.

**Figure S5** provides detail on the flows of treatment completion for each state. Boxes in light grey and dashed borders are duplicates (i.e. not repeated in each treatment history state), but shown here to help representation of state flows.

- Individuals who are placed on treatment from susceptible (S), cleared (C), recovered (R), or treated (Tc) states move to the same state in the previously treated sub-dimension (_P) according to the rate of treatment completion (inverse of treatment duration), which is specific to the treatment regimen (*comp_st, comp_rt*). These transitions are not shown in the figure below.
- Individuals complete treatment successfully (green solid lines), or unsuccessfully (orange dotted lines). Successful treatment completion is defined by the rate of treatment completion (inverse of treatment duration), which is specific to the treatment regimen (*comp_st, comp_rt*), and the probability of treatment success (succ_st, succ_rt), which acknowledges a relative treatment success for individuals with drug-resistant TB who are placed on standard treatment (succ_strs).
    - Infected individuals (I) move to the previously treated cleared state (C_P) if treatment is successful and the previously treated infection state (I_P) if treatment is unsuccessful.
    - Individuals with non-infectious disease (Dn) move to the previously treated recovered state (R_P) if treatment is successful and the previously treated non-infectious disease state (Dn_P) if treatment is unsuccessful.
    - Individuals with asymptomatic disease (Ds) or symptomatic disease (Dc) move to the previously treated recent treatment state (Tr_P) if treatment is successful and the previously treated asymptomatic disease state (Ds_P) or the previously treated symptomatic disease state (Dc_P) if treatment is unsuccessful.
- Individuals with non-infectious disease (Dn) or symptomatic disease (Dc) are also at risk of mortality whilst on treatment, which is specific to the treatment regimen (*mort_st, mort_rt*).

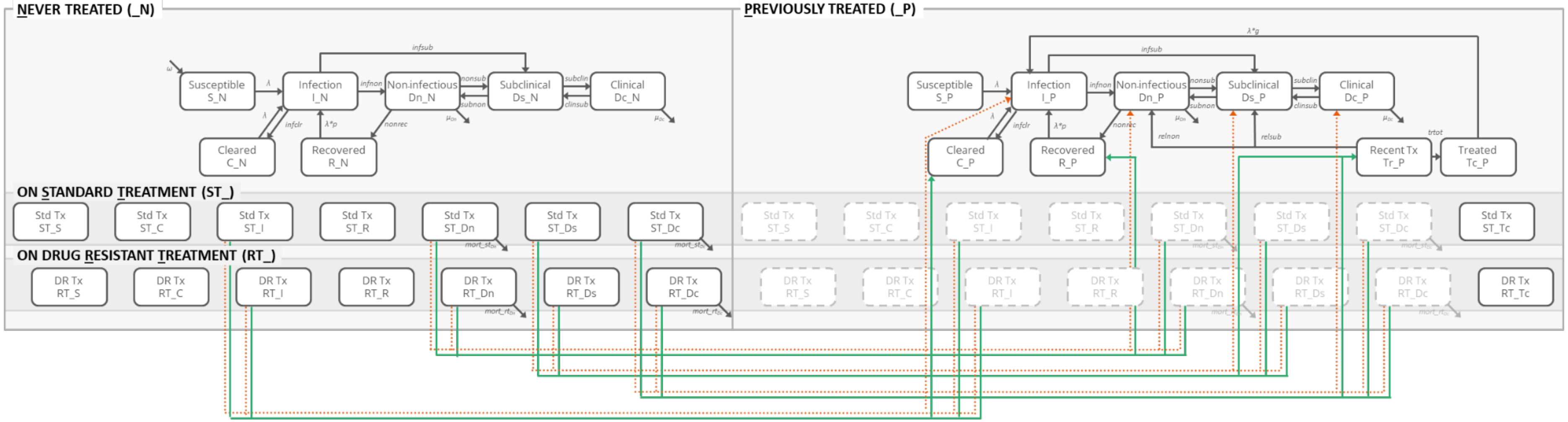

Successful treatment = completion rate * treatment success
Unsuccessful treatment = completion rate * (1 – treatment success – mortality on treatment)
Mortality on treatment = completion rate * mortality on treatment

Figure S5. Treatment completion in diagnosis and treatment structure. TB disease state and parameter naming follow conventions from research over the past decade; results are reported using current WHO terminology.

##### 1.2.3 DRUG RESISTANCE

This section aims to describe in detail the stratification into strain susceptibility.

**Figure S6** adds further stratification of the TB natural history by introducing strain susceptibility on infection and disease states. The figure shows duplicate states (shown in light grey with dashed outlines) which are placed for convenient representation of structure.

- Drug resistance status is modelled using two nearly parallel sub-strata, one for individuals with drug-susceptible strains (_s) and one for individuals with drug-resistant (i.e., rifampicin-resistant) strains (_r).
- Infected (I), recently treated (Tr) and all three disease states (Dn, Ds, Dc) are subdivided based on strain susceptibility.
- Both transmission of drug-resistant strains and acquisition of drug resistance during treatment are modelled.
- Relapse rates are disaggregated by drug-resistance strain (*relnon_s*, *relsub_s*, *relnon_r*, *relsub_r*)

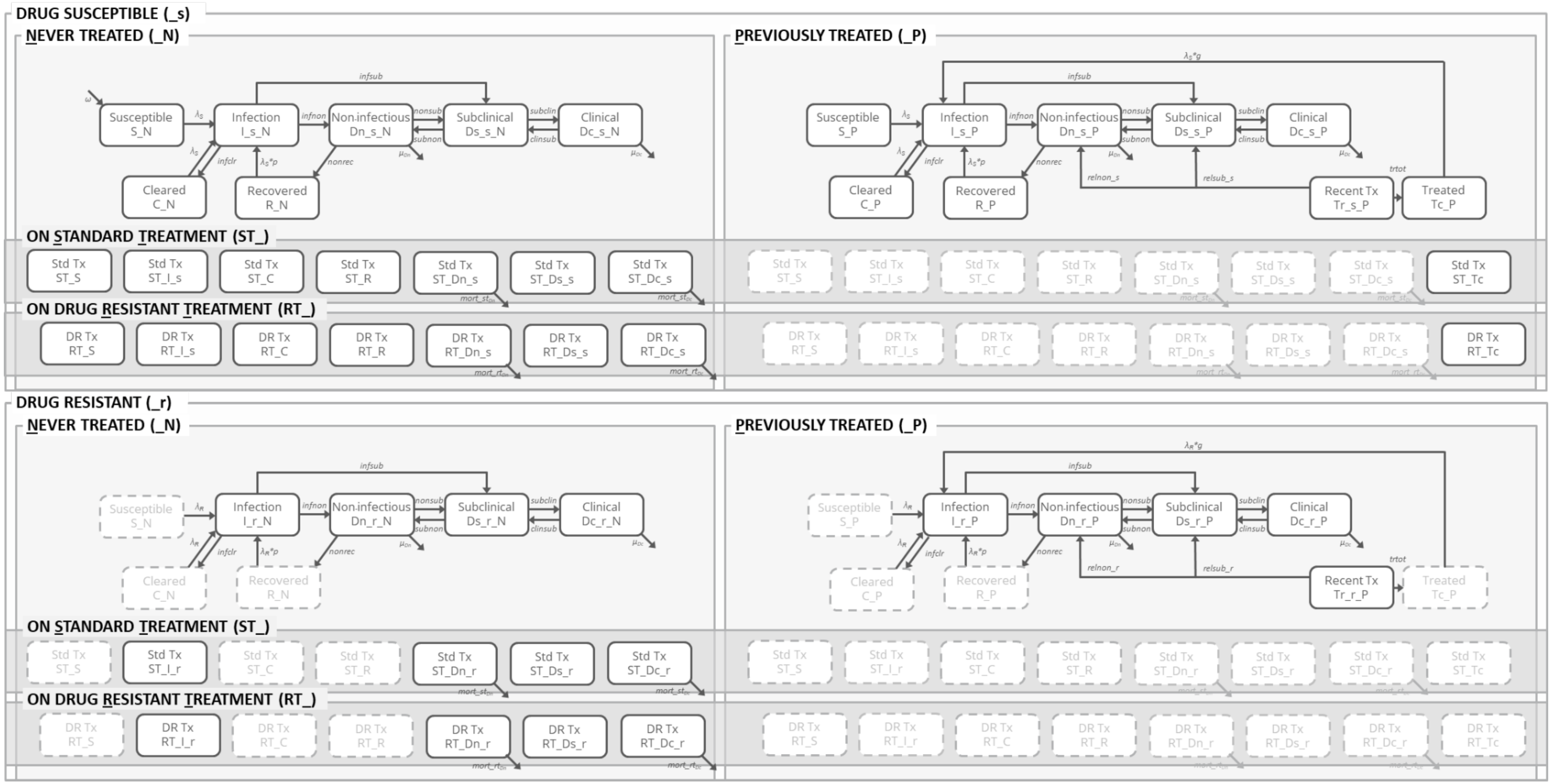

Figure S6. Drug resistance structure. TB disease state and parameter naming follow conventions from research over the past decade; results are reported using current WHO terminology.

**Figure S7** shows the transmission of drug susceptible and drug resistant strains. The figure shows duplicate states (shown in light grey with dashed outlines) which are placed for convenient representation of structure.

- Individuals in susceptible (S), cleared (C), recovered (R), and treatment (Tc) compartments can be infected with drug-susceptible or drug-resistant strains.

- Individuals with drug-susceptible infections (I_s) can be superinfected with drug-resistant strains (solid orange lines); however, it is assumed that there is no superinfection with drug-resistant strains among individuals with established drug-susceptible disease (Dn_s, Ds_s, Dc_s). Superinfection with drug-susceptible strains among individuals with drug-resistant infection (I_r) or disease (Dn_r, Ds_r, Dc_r) is not modelled.
- Transmission of drug-resistant strains acknowledges a fitness cost due to the genetic mutations that confer resistance (see Section 1.2.1 above).
- A fixed proportion of individuals with drug-susceptible infections (I_s) and with drug-susceptible disease (Dn_s, Ds_s, Dc_s) acquire resistance whilst on standard treatment at rate *acqres* (shown in dashed orange lines).

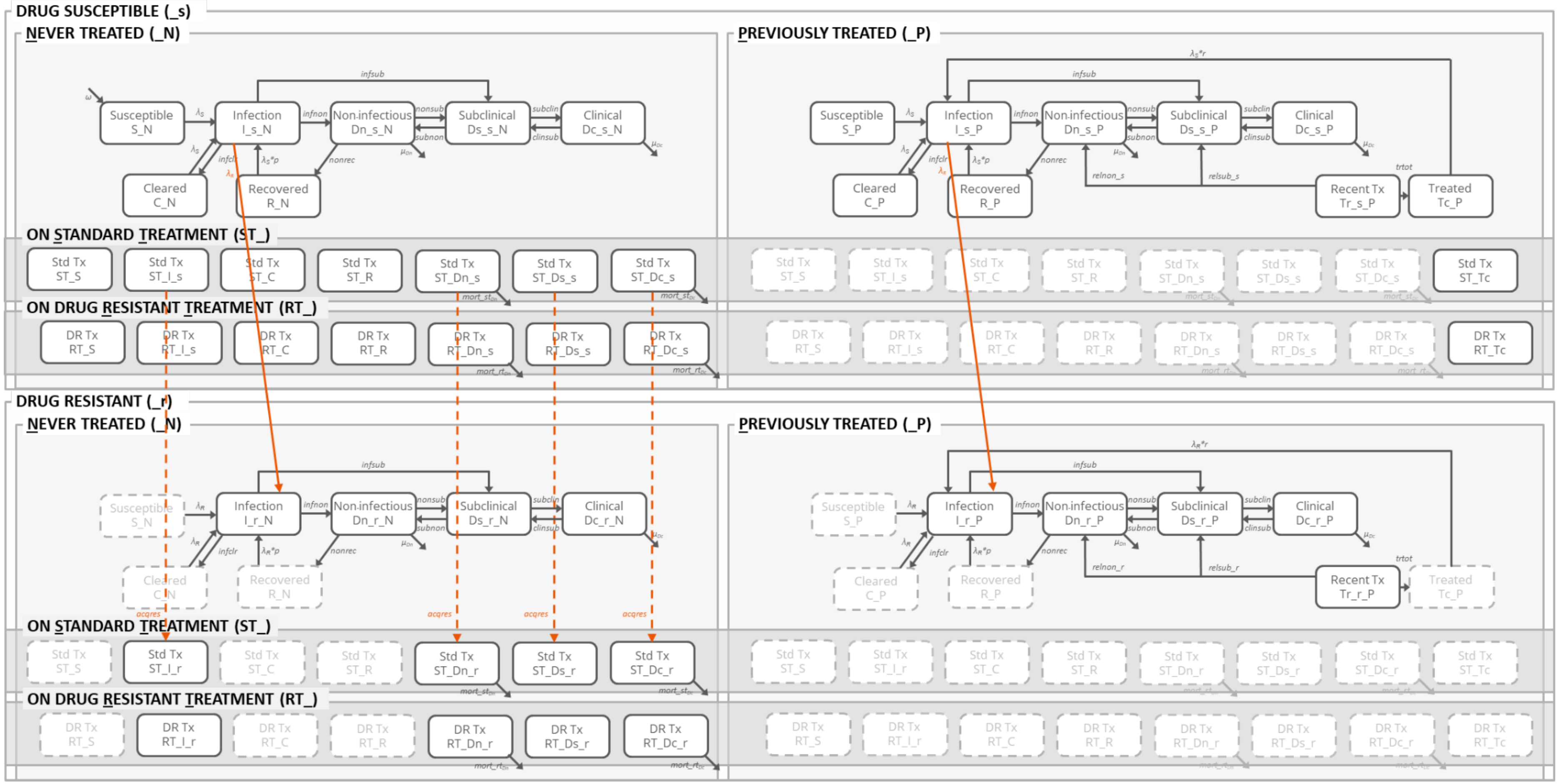

Figure S7. Drug resistance transmission and acquisition in drug resistance structure. TB disease state and parameter naming follow conventions from research over the past decade; results are reported using current WHO terminology.

#### 1.3 PARAMETERS

The following section will look into the parameters in detail, describing their use, their values or ranges and whether or not they are age or time-dependent.

Table S1. TB dimension parameters.

| Parameter | Description (units) | Prior/Value | Age-dependent | Time-dependent | Reference |
|---|---|---|---|---|---|
| pT | Probability of transmission per contact per day | 0.0013<br>See note 1 | No | No | Estimated from data |
| cscal | Multiplier to scale contacts to fit to burden data | ZAF/IND: 0-4<br>BRA: 0-2.5 | No | No | Assumption |
| relfit | Relative fitness of drug-resistant strains relative to drug-susceptible strains | 0.4-0.9 | No | No | (Kendall et al., 2017) |
| k | Relative infectiousness of children compared to adults | 0.25-1.00 | Yes | No | Assumption |
| t | Relative infectiousness of asymptomatic compared to symptomatic | 0.62-1.00 | No | No | (Emery et al., 2023)<br>Range truncated at 1.00 |
| p | Relative risk of infection following recovery from non-infectious | 0.14-0.30 | No | No | (Andrews et al., 2012) |
| g | Relative risk of infection following treatment completion | 2.14-4.27 | No | No | (Glynn et al., 2010; Verver et al., 2005) |
| infclr | Clearance from infection per year | 0.93-3.30 | No | No | (Horton et al., 2023) |
| infnon | Progression from infection to non-infectious per year | 0.04-0.23 | No | No | (Horton et al., 2023) |
| infsub | Progression from infection to asymptomatic per year | 0.01-0.10 | No | No | (Horton et al., 2023) |
| nonrec | Recovery from non-infectious per year | 0.14-0.23 | No | No | (Horton et al., 2023) |
| nonsub | Progression from non-infectious to asymptomatic per year | 0.21-0.28 | Yes | No | (Horton et al., 2023) |
| subnon | Regression from asymptomatic to non-infectious per year | 1.24-2.03 | No | No | (Horton et al., 2023) |
| subclin | Progression from asymptomatic to symptomatic per year | 0.56-0.94 | Yes | No | (Horton et al., 2023) |
| clinsub | Regression from symptomatic to asymptomatic per year | 0.46-0.72 | No | No | (Horton et al., 2023) |
| $\mu_{Dn}$ | Non-infectious TB mortality per year | 0.00-0.45<br>See note 2 | Yes | No | Assumption |
| $\mu_{Dc}$ | Symptomatic TB mortality per year | 0.017-0.45 | No | No | (Ragonnet et al., 2021) |
| tinit | Sigmoidal curve describing rate of treatment initiation | See note 3 | Yes | Yes | Assumption |
| midx | Parameter to define *tinit* | See note 3 | No | No | Assumption |

| | | | | | |
|---|---|---|---|---|---|
| sh | Parameter to define *tinit* | See note 3 | No | No | Assumption |
| eta | Country-specific access factor | ZAF/BRA: 0-1<br>IND: 0-1.5<br>See note 3 | No | No | Assumption |
| relinit | Relative access to TB screening by core TB state | See note 3 | No | No | See note 3 |
| dst_n | Access to drug-susceptibility testing in never treated individuals | See note 4 | No | Yes | (World Health Organization, 2023)<br>WHO case notifications<br>WHO drug resistance testing |
| dst_p | Access to drug-susceptibility testing in previously treated individuals | See note 4 | No | Yes | (World Health Organization, 2023)<br>WHO case notifications<br>WHO drug resistance testing |
| dst_s | Probability of a positive drug-susceptibility test result for uninfected individuals and individuals with drug susceptible TB | 0.01<br>See note 4 | No | No | (Zifodya et al., 2021) |
| dst_r | Probability of a positive drug-susceptibility test result for individuals with drug resistant TB | 0.95<br>See note 4 | No | No | (Zifodya et al., 2021) |
| comp_st | Treatment completion rate for standard treatment per year (1/duration of treatment) | 2.0 | No | Yes | (World Health Organization, 2022b) |
| comp_rt | Treatment completion rate for drug resistant treatment per year (1/duration of treatment) | ZAF/IND: 1.3-2.0<br>BRA: 0.67-2.0 | No | Yes | (World Health Organization, 2022a) |
| succ_st | Probability of treatment success for standard treatment of drug susceptible TB | See note 5 | No | Yes | (World Health Organization, 2023)<br>WHO treatment outcomes |
| succ_rt | Probability of treatment success for drug resistant treatment of drug resistant TB | See note 5 | No | Yes | (World Health Organization, 2023)<br>WHO treatment outcomes |
| mort_st | Probability of death on drug susceptible treatment | See note 5 | No | Yes | (World Health Organization, 2023)<br>WHO treatment outcomes |
| mort_rt | Probability of death on drug resistant treatment | See note 5 | No | Yes | (World Health Organization, 2023)<br>WHO treatment outcomes |
| succ_strs | Relative treatment success of standard treatment for drug resistant TB | 0.35-0.70 | No | No | (Espinal et al., 2000)<br>Range used is lower limit of previously treated and upper |

| | | | | | |
|---|---|---|---|---|---|
| | | | | | limit of never treated |
| relnon_s | Relapse from recently treated to non-infectious after standard treatment | 0.00-0.01 | No | No | Assumption |
| relsub_s | Relapse from recently treated to asymptomatic after standard treatment | 0.00-0.01 | No | No | Assumption |
| relnon_r | Relapse from recently treated to non-infectious after standard treatment | 0.01-0.06 | No | No | (Kendall et al., 2017) |
| relsub_r | Relapse from recently treated to asymptomatic after standard treatment | 0.01-0.06 | No | No | (Kendall et al., 2017) |
| trtot | Transition from recently treated to treated per year | 1<br>See note 6 | No | No | Assumption |
| acqres | Rate of acquisition of rifampicin resistance during standard treatment per year | 0.003-0.012 | No | No | (Kendall et al., 2017) |

Note 1: Probability of transmission per contact (pT)

The daily probability of transmission per respiratory contact between an infectious and healthy individual has a fixed value of 0.0013 (0.0002-0.0048). This value was estimated from the daily mean number of infectious contacts ((Prem et al., 2021; World Health Organization, 2023)) multiplied by the reversion-adjusted annual risk of infection from TST immunoreactivity surveys (Cauthen et al., 2002; Houben & Dodd, 2016).

$$pT = (mC[m,y] * TBprev * (ARI * rev))/365.25$$

where *mC[m,y]* indicates the mean number of age-specific contacts per year
*TBprev* indicates the country specific TB prevalence (%) from WHO
*ARI* indicates annual risk of infection from TST immunoreactivity surveys
*rev* indicates reversion-underestimation factor of ARI (Schwalb et al., 2023)

The parameter *cscal* is a calibration variable that acts upon *pT* and it ranges from 0 to 4 for IND and ZAF, and 0 to 2.5 for BRA.

Note 2: Non-infectious disease mortality per year

Non-infectious disease mortality is included in recognition that non-infectious disease includes both asymptomatic and symptomatic disease (as per ICE-TB framework, (Coussens et al., 2024)) and is particularly severe amongst children. The prior parameter range is defined from 0 to the upper bound of the mortality rate for symptomatic infectious disease (Ragonnet et al., 2021).

Note 3: Treatment initiation rate

Treatment initiation rate (*init*) is defined as follows:

$$init = tinit * eta * relinit$$

where *tinit* indicates a sigmoid curve symbolising the scale-up of treatment access
*eta* indicates a country-specific calibration factor (between 0 and 1)
*relinit* indicates a TB state-specific factor

Tuberculosis treatment was assumed to start in 1960, aligned roughly with the discovery and widespread use of rifampicin, and increase following a sigmoid curve (**Figure S8**).

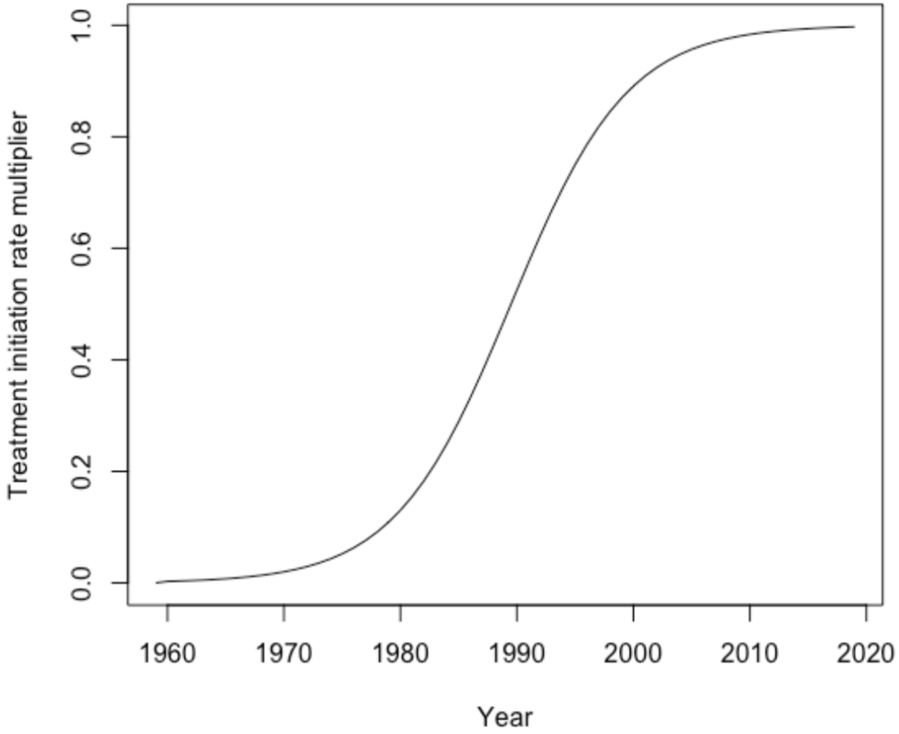

Figure S8. Sigmoid curve representing the scale-up in tuberculosis treatment.

The sigmoid curve *tint* is defined as follows:

$$tinit = \frac{1}{1 + e^{(-sh*(x-midx))}}$$

where *sh* indicates the steepness of the curve
*midx* indicates the midpoint of the curve

These parameters are fixed values defined by each country team.

Table S2. Country-specific sigmoid curve parameters

| Parameter | South Africa | India | Brazil |
|---|---|---|---|
| sh | 0.2 | 0.33 | 0.1 |
| midx | 2003.4 | 1970 | 2000 |

The TB state-specific factor (*relinit*) is used to recognise variability in treatment initiation.To enable adjustment of screening, diagnosis and treatment initiation between TB states each of these is specified separately, informed pre-baseline by:

- Relative access to diagnosis pathway (*access)*: Values are based on screening positivity of prolonged cough, using Dc as reference. We assume a relative access increase of 50% for those previously treated (_P).
- Probability of a positive test (*testpos*): Values are based on country-specific proportions of bacteriologically confirmed and clinically diagnosed notifications, according to WHO datasets, combined with test positivity of Xpert MTB/RIF, smear or culture, and clinical diagnosis.
- Proportion who accept treatment (*accept)*: Values are assumptions, using Dc as reference.

During interventions, values are adjusted to match intervention scale-up.

Table S3. Baseline relative treatment initiation by state

Relinit is equal to access*testpos*accept

| State | Relative access to diagnostic pathway (access) | Probability of a positive test (testpos) | | |
|---|---|---|---|---|
| | | ZAF | IND | BRA |
| S_N | 0.06 | 0.0251 | 0.0294 | [illegible] |
| C_N | 0.06 | 0.0251 | 0.0294 | [illegible] |
| R_N | 0.06 | 0.067 | 0.0821 | [illegible] |
| I_s_N | 0.06 | 0.0251 | 0.0294 | [illegible] |
| I_r_N | 0.06 | 0.0251 | 0.0294 | [illegible] |
| Dn_s_N | 0.19 | 0.0882 | 0.1307 | [illegible] |
| Dn_r_N | 0.19 | 0.0882 | 0.1307 | [illegible] |
| Ds_s_N | 0.06 | 0.4481 | 0.3549 | [illegible] |
| Ds_r_N | 0.06 | 0.4481 | 0.3549 | [illegible] |

| | | | |
|---|---|---|---|
| Dc_s_N | 1.00 | 0.7922 | 0.7599 |
| Dc_r_N | 1.00 | 0.7922 | 0.7599 |
| S_P | 0.09 | 0.0251 | 0.0294 |
| C_P | 0.09 | 0.0251 | 0.0294 |
| R_P | 0.09 | 0.067 | 0.0821 |
| I_s_P | 0.09 | 0.0251 | 0.0294 |
| I_r_P | 0.09 | 0.0251 | 0.0294 |
| Dn_s_P | 0.29 | 0.0882 | 0.1307 |
| Dn_r_P | 0.29 | 0.0882 | 0.1307 |
| Ds_s_P | 0.09 | 0.4481 | 0.3549 |
| Ds_r_P | 0.09 | 0.4481 | 0.3549 |
| Dc_s_P | 1.50 | 0.7922 | 0.7599 |
| Dc_r_P | 1.50 | 0.7922 | 0.7599 |
| Tc_P | 0.09 | 0.2366 | 0.2778 |

Note 4: Drug susceptibility testing (DST)

DST access is parameterised using data from the WHO Global Tuberculosis Programme databases as reported in the diagnosis, notification and treatment of rifampicin-resistant TB dashboard (World Health Organization, 2023). The proportion of individuals tested for rifampicin-resistant resistant is multiplied by the proportion of notifications that are bacteriologically-confirmed.

Based on the sensitivity and specificity of Xpert Mtb/RIF and Xpert Ultra for the detection of rifampicin resistance (Zifodya et al., 2021), we assume that 1% of individuals who do not have drug resistant infection or disease and 95% of individuals who do have drug resistant infection or disease will receive a positive result on drug susceptibility testing.

Table S4. Country-specific DST access by treatment history.

| State | South Africa | India | Brazil |
|---|---|---|---|
| | **Value** | **Value** | **Value** |
| dst_n | 0.621 | 0.4 | 0.318 |

| | | | |
|---|---|---|---|
| dst_p | 0.618 | 0.67 | 0.341 |

Note 5: Treatment success and mortality

This section will describe the parameters used to account for treatment success and mortality.

Treatment success is only relevant to individuals with infection, non-infectious TB, asymptomatic TB, or symptomatic TB, and is parameterised using data from the WHO Global Tuberculosis Programme database on treatment outcomes (World Health Organization, 2023). Transitions following treatment for susceptible, cleared, recovered, and treated individuals are not distinguished by treatment success, and recently treated individuals are not placed on treatment.Treatment success is expected to be higher among infected individuals and individuals with non-infectious TB than among those with bacteriologically positive asymptomatic or symptomatic TB. Only individuals with non-infectious TB and symptomatic TB are at risk of mortality on treatment; mortality is considered as unsuccessful treatment.

Table S5. Definitions of treatment success and mortality by disease state

| State | Treatment success | Mortality on treatment | Un[illegible] |
|---|---|---|---|
| Infected | 1 | 0 | |
| Non-infectious TB [Children] | (1 - mortality on treatment) * Treatment success[1] (defined below) | Mortality on treatment (defined below) | (1 - mortality on [illegible] |
| Non-infectious TB [Adults] | Midpoint between adjusted treatment success[1] (defined below) and 1 | 0 | (1[illegible] |
| Asymptomatic TB | Treatment success (defined below) | 0 | (1[illegible] |
| Symptomatic TB | (1 - mortality on treatment) * Treatment success[1] (defined below) | Mortality on treatment (defined below) | (1 - mortality on [illegible] |

[1] Treatment success of non-infectious and symptomatic TB was adjusted to account for mortality on treatment.

For drug susceptible TB, treatment success is defined as the total number of treatment success outcomes (cured or treatment completed) among new, relapse, and previously treated individuals, divided by the total cohort size of new, relapse, and previously treated individuals:

*Treatment success = (newrel_succ + ret_nrel_succ) / (newrel_coh + ret_nrel_coh)*

Mortality on treatment is defined as the number of deaths on treatment, plus half the number of individuals who are reported as lost to follow-up, among new, relapse, and previously treated individuals, divided by the total cohort size of new, relapse, and previously treated individuals:

*Mortality on treatment = (newrel_died + 0.5*newrel_lost + ret_nrel_died + 0.5*ret_nrel_lost) / (newrel_coh + ret_nrel_coh)*

For drug resistant TB, treatment success is defined as the total number of treatment success outcomes (cured or treatment completed) among individuals with MDR-TB, divided by the total cohort size of individuals with MDR-TB:

*Proportion success treatment* = (mdr_succ) / (mdr_coh)

Mortality on treatment is defined as the number of deaths on treatment, plus half the number of individuals who are reported as lost to follow-up, among individuals with MDR-TB, divided by the total cohort size of individuals with MDR-TB:

*Proportion deaths on treatment* = (mdr_died + 0.5*mdr_lost) / (mdr_coh)

Unsuccessful treatment is defined as the outcome for remaining individuals with MDR-TB:

*Proportion unsuccessful treatment = 1 - (proportion successful treatment + proportion deaths on treatment)*

Table S6. Country-specific treatment success and mortality

| State | South Africa | India | Brazil |
|---|---|---|---|
| | Value | Value | Value |
| succ_stDn / succ_stDc | 0.875 | 0.8617021 | 0.7882353 |
| succ_stDs | 0.77 | 0.81 | 0.67 |
| mort_stDn / mort_stDc | 0.12 | 0.062 | 0.15 |
| succ_rtDn / succ_rtDn | 0.7222222 | 0.7027027 | 0.6875 |
| succ_rtDs | 0.52 | 0.52 | 0.55 |
| mort_rtDn / mort_rtDc | 0.28 | 0.26 | 0.20 |

The value used for treatment success is different between non-infectious and symptomatic TB compared to asymptomatic TB to account for mortality on treatment.

Note 6: Relapse risk

We assume that individuals are at risk of relapse for a period of one year following treatment (Crampin et al., 2010; Marx et al., 2014).

#### 1.4 DIFFERENTIAL EQUATIONS

This section aims to provide the differential equations which operate the model structure described above. Each differential equation describes flows and transitions experienced in each 49 states of TBMod.

##### 1.4.1 EXPRESSIONS

**lambda_all** = pT*cscal*k*(t*(Ds_s_N + Ds_s_P) + Dc_s_N + Dc_s_P + relfit*(t*(Ds_r_N + Ds_r_P) + Dc_r_N + Dc_r_P)) / N

**lambda_s** = pT*cscal*k*(t*(Ds_s_N + Ds_s_P) + Dc_s_N + Dc_s_P) / N

**lambda_r** = pT*cscal*k*relfit*(t*(Ds_r_N + Ds_r_P) + Dc_r_N + Dc_r_P) / N

**treat_init** = tinit*eta*(access*testpos*accept)*dx_age

##### 1.4.2 NEVER TREATED

**dS_N/dt** = -(treat_init + lamda_all)*S_N

**dC_N/dt** = infclr*(I_s_N + I_r_N) - (treat_init + lambda_all)*C_N

**dR_N/dt** = nonrec*(1/prog_age)*(Dn_s_N + Dn_r_N) - (treat_init + lambda_all*p)*R_N

**dI_s_N/dt** = lambda_s*(S_N + C_N + p*R_N) - (infclr + prog_age*(infnon + infsub))*I_s_N - (treat_init + lambda_r)*I_s_N

**dI_r_N/dt** = lambda_r*(S_N + C_N + p*R_N + I_s_N) - (infclr + prog_age*(infnon + infsub) + treat_init)*I_r_N

**dDn_s_N/dt** = infnon*prog_age*I_s_N + subnon*(1/prog_age)*Ds_s_N - (nonrec*(1/prog_age) + nonsub*prog_age + muDn*mort_age + treat_init)*Dn_s_N

**dDn_r_N/dt** = infnon*prog_age*I_r_N + subnon*(1/prog_age)*Ds_r_N - (nonrec*(1/prog_age) + nonsub*prog_age + muDn*mort_age + treat_init)*Dn_r_N

**dDs_s_N/dt** = infsub*prog_age*I_s_N + nonsub*prog_age*Dn_s_N + clinsub*(1/prog_age)*Dc_s_N - (subnon*(1/prog_age) + subclin*prog_age + treat_init)*Ds_s_N

**dDs_r_N/dt** = infsub*prog_age*I_r_N + nonsub*prog_age*Dn_r_N + clinsub*(1/prog_age)*Dc_r_N - (subnon*(1/prog_age) + subclin*prog_age + treat_init)* Ds_r_N

**dDc_s_N/dt** = subclin*prog_age*Ds_s_N - (clinsub*(1/prog_age) + muDc*mort_age + treat_init)*Dc_s_N

**dDc_r_N/dt** = subclin*prog_age*Ds_r_N - (clinsub*(1/prog_age) + muDc*mort_age + treat_init)*Dc_r_N

##### 1.4.3 PREVIOUSLY TREATED

**dS_P/dt** = comp_st*ST_S + comp_rt*RT_S - (treat_init + lambda_all)*S_P

**dC_P/dt** = infclr*(I_s_P + I_r_P) + comp_st*(ST_C + succ_stI*(ST_I_s + succ_strs*ST_I_r)) + comp_rt*(RT_C + succ_rtI*(RT_I_s + RT_I_r)) - (treat_init + lambda_all)*C_P

**dR_P/dt** = nonrec*(1/prog_age)*(Dn_s_P + Dn_r_P) + comp_st*(ST_R + succ_stDn*(ST_Dn_s + succ_strs*ST_Dn_r)) + comp_rt*(RT_R + succ_rtDn*(RT_Dn_s + RT_Dn_r)) - (treat_init + lambda_all*p)*R_P

**dI_s_P/dt** = comp_st*(1-succ_stI)*ST_I_s + comp_rt*(1-succ_rtI)*RT_I_s + lambda_s*(S_P + C_P + p*R_P + r*Tc_P) - (infclr + prog_age*(infnon + infsub*) + treat_init + lambda_r)*I_s_P

**dI_r_P/dt** = comp_st*(1-succ_stI*succ_strs)*ST_I_r + comp_rt*(1-succ_rtI)*RT_I_r + lambda_r*(S_P + C_P + p*R_P + I_s_P + r*Tc_P) - (infclr + prog_age*(infnon + infsub) + treat_init)*I_r_P

**dDn_s_P/dt** = infnon*prog_age*I_s_P + subnon*(1/prog_age)*Ds_s_P + comp_st*(((1-mort_stDn)*mort_age*(1-succ_stDn))+((1-mort_age)*(1-((1+succ_stDn)/2))))*ST_Dn_s + comp_rt*(((1-mort_rtDn)*mort_age*(1-succ_rtDn))+((1-mort_age)*(1-

((1+succ_rtDn)/2)))*RT_Dn_s + relnon_s*Tr_s_P - (nonrec*(1/prog_age) + nonsub*prog_age + muDn*mort_age + treat_init)*Dn_s_P

**dDn_r_P/dt** = infnon*prog_age*I_r_P + subnon*(1/prog_age)*Ds_r_P + comp_st*(((1-mort_stDn)*mort_age*((1-succ_stDn)*succ_strs))+((1-mort_age)*(1-(((1+succ_stDn)/2)*succ_strs)))*ST_Dn_r + comp_rt*(((1-mort_rtDn)*mort_age*(1-succ_rtDn))+((1-mort_age)*(1-((1+succ_rtDn)/2)))*RT_Dn_r + relnon_r*Tr_r_P - (nonrec*(1/prog_age) + nonsub*prog_age + muDn*mort_age + treat_init)*Dn_r_P

**dDs_s_P/dt** = prog_age*(infsub*I_s_P + nonsub*Dn_s_P) + clinsub*(1/prog_age)*Dc_s_P + comp_st*(1-succ_stDs)*ST_Ds_s + comp_rt*(1-succ_rtDs)*RT_Ds_s + relsub_s*Tr_s_P - (subnon*(1/prog_age) + subclin*prog_age + treat_init)*Ds_s_P

**dDs_r_P/dt** = prog_age*(infsub*I_r_P + nonsub*Dn_r_P) + clinsub*(1/prog_age)*Dc_r_P + comp_st*(1-succ_stDs*succ_strs)*ST_Ds_r + comp_rt*(1-succ_rtDs)*RT_Ds_r + relsub_r*Tr_r_P - (subnon*(1/prog_age) + subclin*prog_age + treat_init)*Ds_r_P

**dDc_s_P/dt** = subclin*prog_age*Ds_s_P + comp_st*(1-(mort_stDc)*(1-succ_stDc)*ST_Dc_s + comp_rt*(1-(mort_rtDc)*(1-succ_rtDc)*RT_Dc_s - (clinsub*(1/prog_age) + muDc*mort_age + treat_init)*Dc_s_P

**dDc_r_P/dt** = subclin*prog_age*Ds_r_P + comp_st*(1-(mort_stDc)*(1-succ_stDc*succ_strs)*ST_Dc_r + comp_rt*(1-(mort_rtDc)*(1-succ_rtDc)*RT_Dc_r - (clinsub*(1/prog_age) + muDc*mort_age + treat_init)*Dc_r_P

**dTr_s_P/dt** = comp_st*(succ_stDs*ST_Ds_s + succ_stDc*ST_Dc_s) + comp_rt*(succ_rtDs*RT_Ds_s + succ_rtDc*RT_Dc_s) - (trtot + relnon_s + relsub_s)*Tr_s_P

**dTr_r_P/dt** = comp_st*succ_strs*(succ_stDs*ST_Ds_r + succ_stDc*ST_Dc_r) + comp_rt*(succ_rtDs*RT_Ds_r + succ_rtDc*RT_Dc_r) - (trtot + relnon_r + relsub_r)*Tr_r_P

**dTc_P/dt** = comp_st*ST_Tc + comp_rt*RT_Tc + trtot*(Tr_s_P + Tr_r_P) - (treat_init + lambda_all*g)*Tc_P

###### 1.4.4 ON TREATMENT

**dST_S/dt** = treat_init*((1-dst_n*dst_s)*S_N + (1-dst_p*dst_s)*S_P) - comp_st*ST_S

**dRT_S/dt** = treat_init*dst_s*(dst_n*S_N + dst_p*S_P) - comp_rt*RT_S

**dST_C/dt** = treat_init*((1-dst_n*dst_s)*C_N + (1-dst_p*dst_s)*C_P) - comp_st*ST_C

**dRT_C/dt** = treat_init*dst_s*(dst_n*C_N + dst_p*C_P) - comp_rt*RT_C

**dST_R/dt** = treat_init((1-dst_n*dst_s)*R_N + (1-dst_p*dst_s)*R_P) - comp_st*ST_R

**dRT_R/dt** = treat_init*dst_s*(dst_n*R_N + dst_p*R_P) - comp_rt*RT_R

**dST_I_s/dt** = treat_init*((1-dst_n*dst_s)*I_s_N + (1-dst_p*dst_s)*I_s_P) - (comp_st + acqres)*ST_I_s

**dRT_I_s/dt** = treat_init*dst_s*(dst_n*I_s_N + dst_p*I_s_P) - comp_rt*RT_I_s

**dST_I_r/dt** = acqres*ST_I_s + treat_init*((1-dst_n*dst_r)*I_r_N + (1-dst_p*dst_r)*I_r_P) - comp_st*ST_I_r

**dRT_I_r/dt** = treat_init*dst_r*(dst_n*I_r_N + dst_p*I_r_P) - comp_rt*RT_I_r

**dST_Dn_s/dt** = treat_init*((1-dst_n*dst_s)*Dn_s_N + (1-dst_p*dst_s)*Dn_s_P) - (comp_st + acqres )*ST_Dn_s

**dRT_Dn_s/dt** = treat_init*dst_s*(dst_n*Dn_s_N + dst_p*Dn_s_P) - comp_rt*RT_Dn_s

**dST_Dn_r/dt** = acqres*ST_Dn_s + treat_init*((1-dst_n*dst_r)*Dn_r_N + (1-dst_p*dst_r)*Dn_r_P) - comp_st*ST_Dn_r

**dRT_Dn_r/dt** = treat_init*dst_r*(dst_n*Dn_r_N + dst_p*Dn_r_P) - comp_rt*RT_Dn_r

**dST_Ds_s/dt** = treat_init*((1-dst_n*dst_s)*Ds_s_N + (1-dst_p*dst_s)*Ds_s_p) - (comp_st + acqres)*ST_Ds_s

**dRT_Ds_s/dt** = treat_init*dst_s*(dst_n*Ds_s_N + dst_p*Ds_s_P) - comp_rt*RT_Ds_s

**dST_Ds_r/dt** = acqres*ST_Ds_s + treat_init*((1-dst_n*dst_r)*Ds_r_N + (1-dst_p*dst_r)*Ds_r_P) - comp_st*ST_Ds_r

**dRT_Ds_r/dt** = treat_init*dst_r*(dst_n*Ds_r_N + dst_p*Ds_r_P) - comp_rt*RT_Ds_r

**dST_Dc_s/dt** = treat_init*((1-dst_n*dst_s)*Dc_s_N + (1-dst_p*dst_s)*Dc_s_P) - (comp_st + acqres)*ST_Dc_s

**dRT_Dc_s/dt** = treat_init*dst_s*(dst_n*Dc_s_N + dst_p*Dc_s_P) - comp_rt*RT_Dc_s

**dST_Dc_r/dt** = acqres*ST_Dc_s + treat_init*((1-dst_n*dst_r)*Dc_r_N + (1-dst_p*dst_r)*Dc_r_P) - comp_st*ST_Dc_r

**dRT_Dc_r/dt** = treat_init*dst_r*(dst_n*Dc_r_N + dst_p*Dc_r_P) - comp_rt*RT_Dc_r

**dST_Tc/dt** = treat_init*(1-dst_p*dst_s)*Tc_P - comp_st*ST_Tc

**dRT_Tc/dt** = treat_init*dst_p*dst_s*Tc_P - comp_rt*RT_Tc

### 2. AGE DIMENSION

#### 2.1 OVERVIEW

- The TB dimension described in detail above is further divided into sixteen 5-year age groups.
- TBMod operates in the background with country and age-specific demographics informed by the UN World Population Prospects.

#### 2.2 STRUCTURE

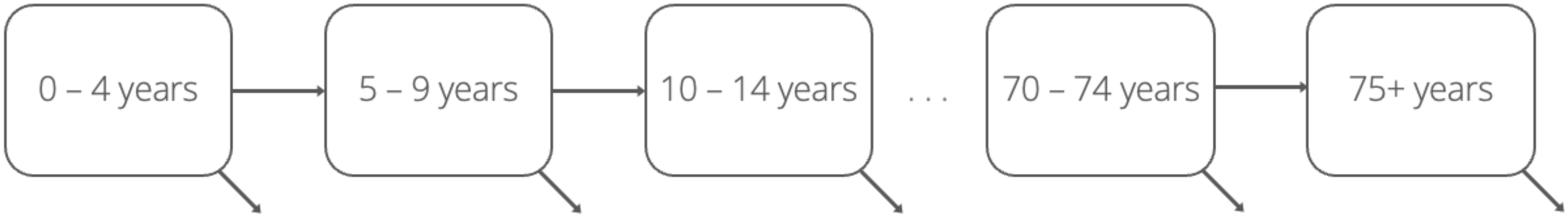

Figure S9. Age dimension structure.

#### 2.3 INTERACTION BETWEEN AGE AND TB DIMENSION

This section will describe how the interaction between age and TB is modelled. We assume that there are differing age-specific parameters in children (<15 years old, i.e. first three age groups) and in adults (≥15 years old, i.e. all other age groups). Particularly, children-specific parameters are either higher or lower when compared to their adult counterpart.

**Figure S10** shows where the age-specific adjustment factors are applied in the TB natural history structure. Parameters follow a naming convention where first it specifies where the parameter is acting upon (i.e., *prog* in progression flows) and then which dimension it refers to, in this case *age*. An additional age-specific adjustment (*dx_age*) is not shown but it acts upon treatment initiation.

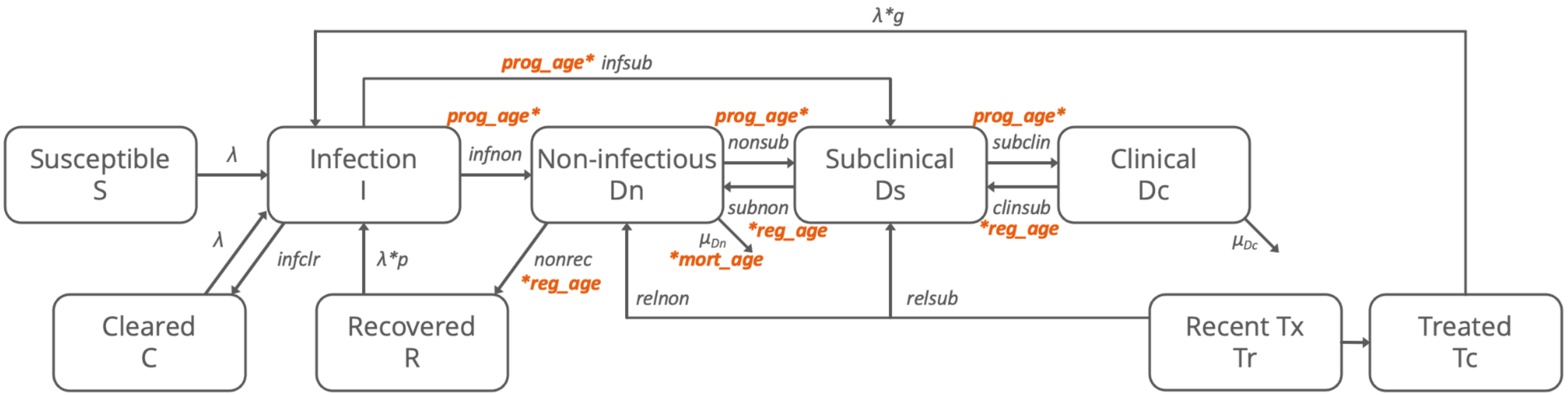

Figure S10. Age adjustments to transitions in the TB dimension. Detailed treatment pathways are shown in TB dimension. TB disease state and parameter naming follow conventions from research over the past decade; results are reported using current WHO terminology.

#### 2.4 EQUATIONS

Equations incorporating age are shown above in Section 1.3.

#### 2.5 PARAMETERS

Table S7. Age and TB interaction parameters

| Parameter | Description | Prior/Value | Age-dependent? | Time-dependent? | Reference |
|---|---|---|---|---|---|
| prog_age | Relative progression of disease in children | 0-14 years: 0.5 - 1.0 15+ years: 1.0 | Yes | No | Assumption |
| reg_age | Relative regression of disease in children | 1/prog_age | Yes | No | Assumption |
| mort_age | Relative non-infectious TB mortality in children | 0-14 years: 1.0 15+ years: 0.0 | Yes | No | Assumption |

| | | | | | |
|---|---|---|---|---|---|
| dx_age | Relative diagnostic rate in children | 0-14 years:<br>0.25 - 1.0<br>15+ years | Yes | No | Assumption |

- Decreased transmission in children is modelled with the use of age-specific contact matrices for each country.
- An adjustment factor (*prog_age*) is applied to parameters *nonsub* and *subclin* to account for the lower progression of disease from non-infectious (Dn) to asymptomatic disease (Ds) and from asymptomatic disease (Ds) to symptomatic disease (Dc) in children compared to adults.
- An adjustment factor (*mort_age)* is applied to the $\mu_{Dn}$ parameter to only account for non-infectious TB mortality in children. This is done to account for increased TB mortality in children despite not observing a higher number of symptomatic cases compared to adults.
- An adjustment factor (*dx_age*) is applied to the *access* parameter to account for lower care-seeking behaviour/access to diagnostics in children compared to adults.
- An adjustment factor (*k*) is applied to account for lower relative infectivity in asymptomatic (Ds) and symptomatic disease (Dc) in children compared to adults (see Section 1.2.1 for more detail).

### 4. RISK DIMENSION: HIV IN SOUTH AFRICA

We modelled an additional key risk factor for TB in our three model countries. For South Africa, this was HIV.

#### 4.1 OVERVIEW

- There are three compartments modelling HIV progression in this risk dimension: HIV uninfected (*HIV0*), HIV infected not on ART (*HIVnt*), and HIV infected on ART(*HIVart*) (shown in Figure S11).
- HIV transmission is not modelled. Instead, a HIV infection incidence data is used to simulate progression from HIV uninfected to HIV infected not on ART.  Increased risk of death if infected with HIV, mitigated by ART by 90%. Individuals infected with HIV and receiving ART can discontinue ART treatment and regress to HIV infected not on ART.
- Progression from one HIV state to another can happen at any state of the natural history of TB.
- Depending on the HIV state, some parameters modelling TB natural history and treatment may increase or decrease (more detail in section 4.3)

#### 4.2 STRUCTURE

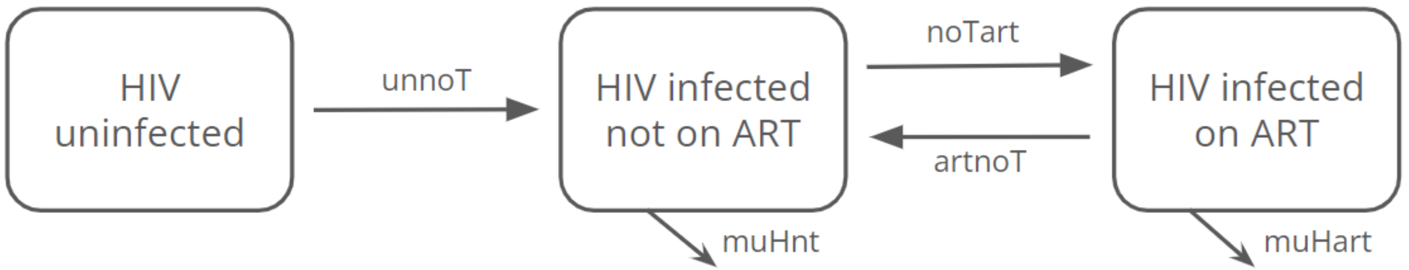

Figure S11. HIV dimension structure

When individuals are not infected with HIV, they are in the *HIV0* (HIV uninfected) compartment. They transition at rate *unnoT* to go to the *HIVnt* compartment, individuals that are HIV infected but are not on ART. In this compartment they can die at a rate *muHnt* or initiate treatment (*noTart*) and move into the *HIVart* compartment (HIV infected on ART). The individuals in this compartment can also die, but at a different rate (*muHart*), since being on ART decreases the mortality rate, or move back to the *HIVnt* compartment if there is discontinuation of treatment (*artnoT*).

- HIV parameters were mostly the same from previous paper using TBMod ((Sumner et al., 2024).
- HIV mortality in *HIVnt* was modified to be overall instead of by CD4 count. For that, the mean mortality rate was calculated to estimate HIV mortality for those not on treatment (*muHnt*) (Stover et al., 2019). The mean was assumed to be the maximum value and 0 assumed to be the minimum for the range 0 since the smallest values on the table from Stover et al. were close to it.
- *muHart* was calculated as the adjusted rate ratio of HIV mortality in *HIVart* compared to *HIVnt* times *muHnt*

#### 4.3 INTERACTION BETWEEN HIV AND TB DIMENSION

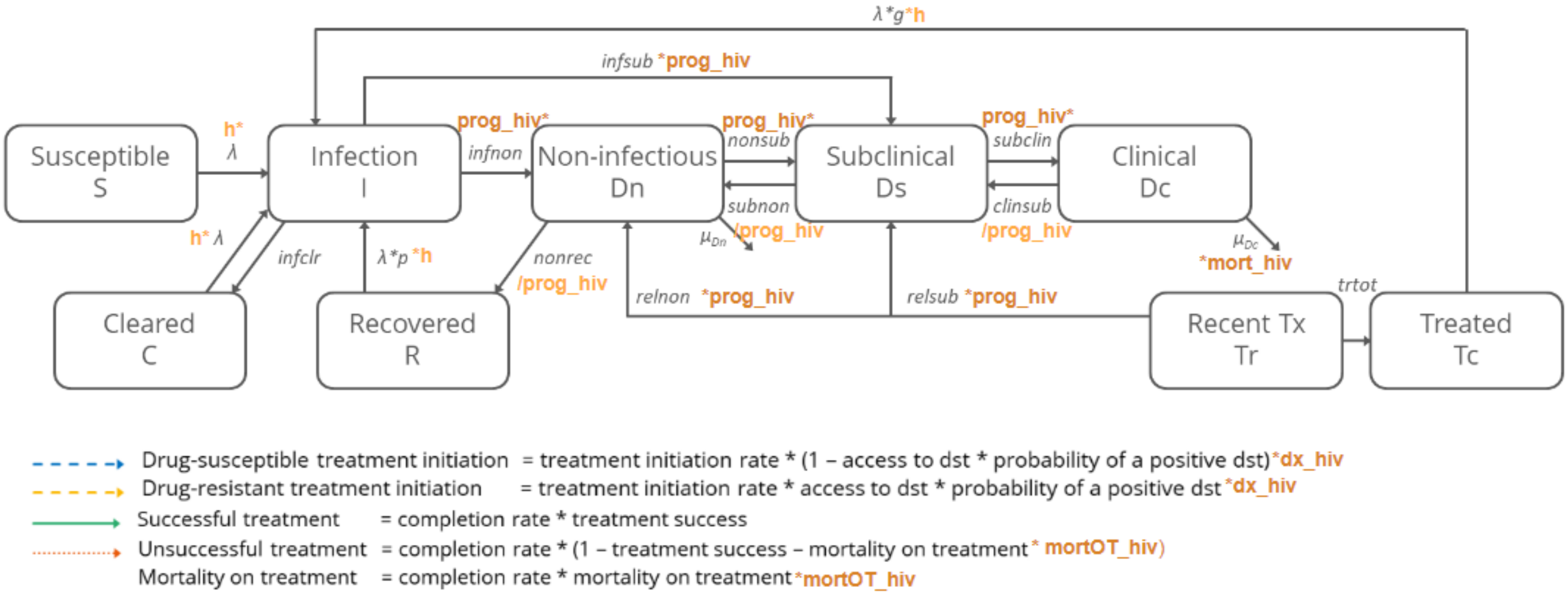

Figure S12. HIV adjustments to transitions in the TB dimension. The diagram shows an overview of which TB parameters are affected by HIV. In particular, there are two types of parameters: those that increase the TB parameter (dark orange) and those that decrease the value of the TB parameter (light orange). Note the coloured arrows in the bottom of the figure represent the treatment pathways in order to show the impact of HIV in them. TB disease state and parameter naming follow conventions from research over the past decade; results are reported using current WHO terminology.

- TB progression (*prog_hiv*):
  - Despite the impact of HIV on TB having been investigated in multiple studies, the exact changes HIV has on the natural history of TB have not been fully assessed. For this reason and simplicity, we decided to increase TB progression and decrease natural recovery by the same value. More details on this assumption can be found on the following bullet points.
    - TB progression (*prog_hiv*): It is used to increase all the rates from infected to symptomatic (and relapse).
      - Interaction equation: TB_parameter *(hiv0str + *prog_hiv* * *prog_hivstr*).
      - Note: *hiv0str* is a parameter with value 1 for *HIV0* and value 0 for *HIVnt* and *HIVart*. This way when *prog_hivstr* is 0 for *HIV0* the value of the TB parameter will not be affected, while still maintain the ratio between progression in *HIVnt* and progression in *HIVart*.
      - From non-infectious to symptomatic, because specific data to inform each transition were not found so the incidence rates found for incidence of disease were assumed to be the same for everyone.
      - In the parameters from infected to non-infectious and asymptomatic, it was assumed that, due to a weaker immune system, they would progress faster once infected. No data was found specifically for each progression parameter, so it was assumed to use the same relative rate as other progression parameters.
      - In risk of relapse, HIV infected were also assumed to have a weaker immune system and hence, a higher risk of relapsing.
    - Natural recovery (1/*prog_hiv*):
      - Interaction equation: TB_parameter/(hiv0str + *prog_hiv* * *prog_hivstr*)
      - HIV infected have weaker immune systems i.e., the rate at which they can recover naturally from the disease would be lower, hence we decrease this rate.
      - Since no information was available on this parameter, it was assumed to decrease by the same amount as TB progression increases.
- HIV mortality (*mort_hiv*):
  - Interaction equation: TB_parameter * (hiv0str + *mort_hiv* * *mort_hivstr*)
  - HIV infected have weaker immune systems i.e., Increased risk of death by TB compared to HIV uninfected.
  - It only affects *Dc* because in *Dn* only children die, and although it is assumed to increase the risk equally between children and

adults, the studies used to estimate the range only include adults, so we decided to assume this only for *Dc*.

- TB infectiousness (*h*):
  - Interaction equation: TB_parameter * (hiv0str + *h*)
  - Infectiousness is different between HIV+ and HIV- according to multiple papers (Crampin et al., 2006; Winter et al., 2020).
  - It was calculated assuming that HIV infected individuals were more likely to be smear negative and that smear negative TB was less infectious.
- TB diagnosis (*dx_hiv*):
  - Interaction equation: TB_parameter * (hiv0str + *dx_hiv* * *dx_hivstr*)
  - HIV infected individuals are more likely to be screened for TB and hence, more likely to start treatment.
- Mortality on TB treatment (*mortOT_hiv*):
  - Interaction equation: TB_parameter * (hiv0str + *mortOT_hiv* * *mortOT_hivstr*)
  - HIV infected individuals have a weaker immune system and, therefore, worse outcomes in TB treatment than HIV uninfected.
  - Both *HIVart* and *HIVnt* are assumed to be the same since in South Africa, the percentage of people on ART is higher than 80% and those that are not on ART and start treatment for TB are suggested to start also on ART between 2 and 8 weeks after initiation of TB treatment, thus, it was assumed for simplicity that both HIV infected states would have the same impact on treatment mortality (South Africa Department of Health, 2014).
- TB treatment success:
  - Not affected by HIV status
  - A study conducted in South Africa suggested that TB treatment success was not significantly different between HIV unifected and HIV infected (Nyasulu et al., 2022). On the other hand, a study in Kenya suggested that treatment success was significantly different between both groups (Owiti et al., 2015). Nonetheless, ART coverage in Kenya is much lower than in South Africa, and since neither study stratified by ART status, this would suggest that perhaps HIV infected not on ART may have worse outcomes but TB treatment success on HIV infected on ART is similar to HIV uninfected (Nyasulu et al., 2022; Owiti et al., 2015). As mentioned in “TB treatment mortality” section, ART coverage in South Africa is high and those not on ART when they initiate TB treatment are linked to HIV care (South Africa Department of Health, 2014). For these reasons and simplicity of the model, it was assumed that TB treatment success did not differ by HIV status.
- TB treatment completion
  - Not affected by HIV status
  - A study in South Africa showed similar loss of people in both HIV unifected and HIV infected groups (Naidoo et al., 2017).

#### 4.4 EQUATIONS

Below shows the equations used in the risk dimension: HIV in South Africa. On treatment equations are not included, due to there being no differences from core in section 1.4. Differences from core are noted using orange.

##### 4.4.1 EXPRESSIONS

**lambda_all =** pT * cscal* pEC * k * (hiv0str - h) * ((t*(Ds_s_N + Ds_s_P) + (Dc_s_N +Dc_s_P) + (relfit *( t*(Ds_r_N + Ds_r_P) + (Dc_r_N +Dc_r_P)))/N)

**lambda_s =** pT * cscal* pEC * k * (hiv0str - h) * ((t*(Ds_s_N + Ds_s_P) + (Dc_s_N +Dc_s_P)/N)

**lambda_r =** pT * cscal * pEC * k * (hiv0str - h) * (relfit * (t*(Ds_r_N + Ds_r_P) + (Dc_r_N +Dc_r_P))/N)

**treat_init =** relinit * tinit * eta* dx_age * (hiv0str + dx_hiv * dx_hivstr)

##### 4.4.2 HIV DIMENSION EQUATIONS

**dHIV0/dt =** unnoT * HIV0

**dHIVnt/dt =** + artnoT * HIVart - muHnt * HIVnt - age_noTart * noTart * HIVnt

**dHIVart/dt =** + age_noTart * noTart * HIVnt - artnoT * HIVart - muHnt * 0.1 * HIVart

##### 4.4.2 NEVER TREATED

**dS_N/dt =** -(treat_init + lamda_all)*S_N

**dC_N/dt =** infclr*(I_s_N + I_r_N) - (treat_init + lambda_all)*C_N

**dR_N/dt =** nonrec*(1/prog_age)/(hiv0str + prog_hiv*prog_hivstr)*(Dn_s_N + Dn_r_N) - (treat_init + lambda_all*p)*R_N

**dI_s_N/dt =** lambda_s*(S_N + C_N + p*R_N) - prog_age*(hiv0str + prog_hiv*prog_hivstr)*(infnon + infsub)*I_s_N - (treat_init + lambda_r+infclr)*I_s_N
**dI_r_N/dt =** lambda_r*(S_N + C_N + p*R_N + I_s_N) - prog_age*(hiv0str + prog_hiv*prog_hivstr)*(infnon + infsub)*I_r_N – (treat_init+infclr)*I_r_N

**dDn_s_N/dt =** infnon*prog_age*(hiv0str + prog_hiv*prog_hivstr)*I_s_N + subnon*(1/prog_age)/(hiv0str + prog_hiv*prog_hivstr)*Ds_s_N - nonrec*(1/prog_age)/(hiv0str + prog_hiv*prog_hivstr)*Dn_s_N - nonsub*prog_age*(hiv0str + prog_hiv*prog_hivstr)*Dn_s_N - muDn*mort_age*Dn_s_N - treat_init*Dn_s_N

**dDn_r_N/dt =** infnon*prog_age*(hiv0str + prog_hiv*prog_hivstr)*I_r_N + subnon*(1/prog_age)/(hiv0str + prog_hiv*prog_hivstr)*Ds_r_N - nonrec*(1/prog_age)/(hiv0str + prog_hiv*prog_hivstr)*Dn_r_N - nonsub*prog_age*(hiv0str + prog_hiv*prog_hivstr)*Dn_r_N - muDn*mort_age*Dn_r_N - treat_init*Dn_r_N

**dDs_s_N/dt =** infsub*prog_age*(hiv0str + prog_hiv*prog_hivstr)*I_s_N + nonsub*prog_age*(hiv0str + prog_hiv*prog_hivstr)*Dn_s_N + clinsub*(1/prog_age)/(hiv0str + prog_hiv*prog_hivstr)*Dc_s_N - subnon*(1/prog_age)/(hiv0str + prog_hiv*prog_hivstr)*Ds_s_N - subclin*prog_age*(hiv0str + prog_hiv*prog_hivstr)*Ds_s_N - treat_init*Ds_s_N

**dDs_r_N/dt =** infsub*prog_age*(hiv0str + prog_hiv*prog_hivstr)*I_r_N + nonsub*prog_age*(hiv0str + prog_hiv*prog_hivstr)*Dn_r_N + clinsub*(1/prog_age)/(hiv0str + prog_hiv*prog_hivstr)*Dc_r_N - subnon*(1/prog_age)/(hiv0str + prog_hiv*prog_hivstr)*Ds_r_N - subclin*prog_age*(hiv0str + prog_hiv*prog_hivstr)*Ds_r_N - treat_init*Ds_r_N

**dDc_s_N/dt =** subclin*prog_age*(hiv0str + prog_hiv*prog_hivstr)*Ds_s_N - clinsub*(1/prog_age)/(hiv0str + prog_hiv*prog_hivstr)*Dc_s_N - muDc*(hiv0str + mort_hiv*mort_hivstr)*Dc_s_N - treat_init*Dc_s_N

**dDc_r_N/dt =** subclin*prog_age*(hiv0str + prog_hiv*prog_hivstr)*Ds_r_N - clinsub*(1/prog_age)/(hiv0str + prog_hiv*prog_hivstr)*Dc_r_N - muDc*(hiv0str + mort_hiv*mort_hivstr)*Dc_r_N - treat_init*Dc_r_N

##### 4.4.2 PREVIOUSLY TREATED

**dS_P/dt =** comp_st*ST_S + comp_rt*RT_S - (treat_init + lambda_all)*S_P

**dC_P/dt =** infclr*(I_s_P + I_r_P) + comp_st*(ST_C + succ_stI*(ST_I_s + succ_strs*ST_I_r)) + comp_rt*(RT_C + succ_rtI*(RT_I_s + RT_I_r)) - (treat_init + lambda_all)*C_P

**dR_P/dt =** nonrec*(1/prog_age)/(hiv0str + prog_hiv*prog_hivstr)*(Dn_s_P + Dn_r_P) + comp_rt* + comp_st*(ST_R+succ_stDn*(ST_Dn_s + succ_strs*ST_Dn_r)) + comp_rt*(RT_R+succ_rtDn*(RT_Dn_s +RT_Dn_r)) - (treat_init + lambda_all*p)*R_P

**dI_s_P/dt =** (1-succ_stI)*(comp_st*ST_I_s + comp_rt*RT_I_s) +lambda_s*(S_P +C_P + p*R_P + g*Tc_P) - infclr*I_s_P - infnon*prog_age*(hiv0str + prog_hiv*prog_hivstr)*I_s_P - infsub*prog_age*(hiv0str + prog_hiv*prog_hivstr)*I_s_P – (treat_init + lambda_r)*I_s_P

**dI_r_P/dt =** comp_st*(1-succ_stI*succ_strs)*ST_I_r + comp_rt*(1-succ_rtI)*RT_I_r + lambda_r*(S_P + C_P + p*R_P + I_s_P + r*Tc_P) - (infclr + prog_age*(hiv0str + prog_hiv*prog_hivstr)*(infnon + infsub)*I_r_P + treat_init)*I_r_P

**dDn_s_P/dt =** infnon*prog_age*(hiv0str + prog_hiv*prog_hivstr)*I_s_P + subnon*(1/prog_age)/(hiv0str + prog_hiv*prog_hivstr)*Ds_s_P + comp_st*(((1-mort_stDn)*mort_age*(1-succ_stDn))+((1-mort_age)*(1-((1+succ_stDn)/2))))*ST_Dn_s + comp_rt*((1-mort_rtDn)*mort_age*(1-succ_rtDn))+((1-mort_age)*(1-((1+succ_rtDn)/2)))*RT_Dn_s + relnon_s*(hiv0str + prog_hiv*prog_hivstr)*Tr_s_P - nonrec*(1/prog_age)/(hiv0str + prog_hiv*prog_hivstr)*Dn_s_P - nonsub*prog_age*(hiv0str + prog_hiv*prog_hivstr)*Dn_s_P - muDn*mort_age*(hiv0str + mort_hiv*mort_hivstr)*Dn_s_P - treat_init*Dn_s_P

**dDn_r_P/dt =** infnon*prog_age*(hiv0str + prog_hiv*prog_hivstr)*I_r_P + subnon*(1/prog_age)/(hiv0str + prog_hiv*prog_hivstr)*Ds_r_P + comp_st*(((1-mort_stDn)*mort_age*((1-succ_stDn)*succ_strs))+((1-mort_age)*(1-(((1+succ_stDn)/2)*succ_strs)))*ST_Dn_r + comp_rt*(((1-mort_rtDn)*mort_age*(1-succ_rtDn))+((1-mort_age)*(1-((1+succ_rtDn)/2)))*RT_Dn_r + relnon_r*(hiv0str + prog_hiv*prog_hivstr)*Tr_r_P - nonrec*(1/prog_age)/(hiv0str + prog_hiv*prog_hivstr)*Dn_r_P - nonsub*prog_age*(hiv0str + prog_hiv*prog_hivstr)*Dn_r_P - muDn*mort_age*(hiv0str+ mort_hiv*mort_hivstr)*Dn_r_P - (treat_init)*Dn_r_P

**dDs_s_P/dt =** infsub*prog_age*(hiv0str + prog_hiv*prog_hivstr)*I_s_P + nonsub*prog_age*(hiv0str + prog_hiv*prog_hivstr)*Dn_s_P + clinsub*(1/prog_age)/(hiv0str + prog_hiv*prog_hivstr)*Dc_s_P + comp_st*(1-succ_stDs)*ST_Ds_s + comp_rt*(1-succ_rtDs)*RT_Ds_s + relsub_s*(hiv0str + prog_hiv*prog_hivstr)*Tr_s_P - subnon*(1/prog_age)/(hiv0str + prog_hiv*prog_hivstr)*Ds_s_P - subclin*prog_age*(hiv0str + prog_hiv*prog_hivstr)*Ds_s_P - treat_init*Ds_s_P

**dDs_r_P/dt =** infsub*prog_age*(hiv0str + prog_hiv*prog_hivstr)*I_r_P + nonsub*prog_age*(hiv0str + prog_hiv*prog_hivstr)*Dn_r_P + clinsub*(1/prog_age)/(hiv0str + prog_hiv*prog_hivstr)*Dc_r_P + comp_st*(1-succ_stDs*succ_strs)*ST_Ds_r + comp_rt*(1-succ_rtDs)*RT_Ds_r + relsub_r*(hiv0str + prog_hiv*prog_hivstr)*Tr_r_P + comp_st*(1-succ_stDs*succ_strs)*ST_Ds_r + comp_rt*(1-succ_rtDs)*RT_Ds_r - subnon*(1/prog_age)/(hiv0str + prog_hiv*prog_hivstr)*Ds_r_P - subclin*prog_age*(hiv0str + prog_hiv*prog_hivstr)*Ds_r_P - treat_init*Ds_r_P

**dDc_s_P/dt =** subclin*prog_age*(hiv0str + prog_hiv*prog_hivstr)*Ds_s_P + comp_st*(1-(mort_stDc*(hiv0str+mortOT_hiv*mortOT_hivstr)))*(1-succ_stDc)*ST_Dc_s + comp_rt*(1-(mort_rtDc*(hiv0str+mortOT_hiv*mortOT_hivstr)))*(1-succ_rtDc)*RT_Dc_s - clinsub*(1/prog_age)/(hiv0str + prog_hiv*prog_hivstr)*Dc_s_P - muDc*(hiv0str + mort_hiv*mort_hivstr)*Dc_s_P - treat_init*Dc_s_P

**dDc_r_P/dt =** subclin*prog_age*(hiv0str + prog_hiv*prog_hivstr)*Ds_r_P + comp_st*(1-(mort_stDc*(hiv0str+mortOT_hiv*mortOT_hivstr)))*(1-succ_stDc*succ_strs)*ST_Dc_r + comp_rt*(1-(mort_rtDc*(hiv0str+mortOT_hiv*mortOT_hivstr)))*(1-succ_rtDc)*RT_Dc_r - clinsub*(1/prog_age)/(hiv0str + prog_hiv*prog_hivstr)*Dc_r_P - muDc*(hiv0str + mort_hiv*mort_hivstr)*Dc_r_P - treat_init*Dc_r_P

**dTr_s_P/dt =** comp_st*succ_stDs*ST_Ds_s + comp_rt*succ_rtDs*RT_Ds_s + comp_st*succ_stDc*ST_Dc_s + comp_rt*succ_rtDc*RT_Dc_s - trtot*Tr_s_P - relnon_s*(hiv0str + prog_hiv*prog_hivstr)*Tr_s_P - relsub_s*(hiv0str + prog_hiv*prog_hivstr)*Tr_s_P

**dTr_r_P/dt =** comp_st*succ_stDs*succ_strs*ST_Ds_r + comp_rt*succ_rtDs*RT_Ds_r + comp_st*succ_stDc*succ_strs*ST_Dc_r + comp_rt*succ_rtDc*RT_Dc_r - trtot*Tr_r_P - relnon_r*(hiv0str + prog_hiv*prog_hivstr)*Tr_r_P - relsub_r*(hiv0str + prog_hiv*prog_hivstr)*Tr_r_P

**dTc_P/dt =** comp_st*ST_Tc + comp_rt*RT_Tc + trtot*(Tr_s_P +Tr_r_P) - (treat_init + lambda_all*r)*Tc_P

#### 4.5 PARAMETERS

The parameter table details the HIV dimension parameters, which include HIV and TB specific interaction parameters; categorised by prior/value, age and time dependency. References are given where applicable.

Table S8. HIV dimension parameters

| Parameter | Description (units) | Prior/Value | Age-dependent? | Time-dependent? | Reference |
|---|---|---|---|---|---|
| HIV parameters | | | | | |
| | Incidence rate of HIV (per person year) | Trend data | Yes | Yes | (Stover & Glaubius, 2024) |
| unnoT[1] | Multiplier of the incidence rate of HIV | 0 - 2.5 | No | No | Fitted in calibration |
| noTart | Rate of treatment initiation (per year) | 0.1 - 3 | Yes | Yes | |
| noTart2010[2] | Multiplier in 2010 of noTart | 0.1 - 3 | No | No | |
| age_noTart | Relative risk of treatment initiation by age | 0.75 | 0-14 | No | (Sumner et al., 2024) |
| | | 1 | 15+ | No | |
| artnoT | Rate of treatment discontinuation (per year) | 0.092 | 0-14 | No | (Mberi et al., 2015; Van Cutsem et al., 2011) |
| | | 0.074 | +15 | No | (Chandiwana et al., 2018; Fenner et al., 2010; Sengayi et al., 2013) |
| prog_hiv[3] | Rate ratio of TB progression | 2.48 - 8 | No | No | (Corbett et al., 2003; Sonnenberg et al., 2005) |
| 1/prog_hiv[3] | Natural recovery | Inverse of TB progression | No | No | Assumption |
| prog_hivstr | For TB transmission | *HIV0* = 0 | No | No | (Lawn et al., 2006) |
| | | *HIVnt* = 1 | | | |
| | | *HIVart* = 0.09-0.38 | | | |
| mort_hivstr | For TB mortality | *HIV0* = 0 | No | No | (Yang et al., 2023) |
| | | *HIVnt* = 1 | | | |
| | | *HIVart* = 0.19-0.82 | | | |
| mortOT_hivstr | For mortality on TB treatment | *HIV0* = 0 | No | No | Assumption |
| | | *HIVnt* = 1 | | | |
| | | *HIVart* = 1 | | | |
| dx_hivstr | For TB treatment | *HIV0* = 0 | No | No | Assumption |
| | | *HIVnt* = 1 | | | |

| | | |
|---|---|---|
| | | *HIVart* = 1 |

[1] HIV transmission is not modelled directly. Instead, a file containing the incidence rate of HIV (per person-year) across ages and time is introduced. This file is obtained from the model Spectrum. The purpose of unnoT is to multiply the rates in that file to adjust the HIV prevalence in the model to that observed in UNAIDS (Joint United Nations Programme on HIV/AIDS, 2021, 2023).
[2] Treatment initiation (*noTart*) varies by age and time. In order to model the changes in time of ART introduction, we added a multiplier called *noTartmul*. This multiplier has specified 3 timepoints: 2003, 2010 and 2020. For 2003 and 2020 the values are fixed. While in 2010, the value is fitted during calibration.
[3] Both TB progression and natural recovery use the same parameter name because prog_hiv is sampled and its inverse is used in the interaction with the respective TB parameter.
[4] The lower bound of dx_hiv is assumed to be lower than in the paper because the study only included places in South Africa with >400 notifications/100,000. Therefore, the range could be lower across the country since areas with less TB may be less focused on TB diagnosis in people living with HIV.

#### 5. RISK DIMENSION: NUTRITION IN INDIA

We modelled an additional key risk factor for TB in our three model countries. For India, this was undernutrition.

##### 5.1 OVERVIEW

- An increased risk of TB disease, unsuccessful TB treatment, and TB mortality due to undernutrition—based on previous modelling exercises (McQuaid et al., 2025)—was captured by dividing the risk dimension in TBMod into four Body Mass Index (BMI) states:
  - Overweight or obese (>25.0 kg/m$^2$)
  - Normal BMI (18.5-24.9 kg/m$^2$)
  - Mild undernutrition (17.0-18.4 kg/m$^2$)
  - Moderate or severe undernutrition(<17 kg/m$^2$)
- BMI distribution in the population from 1974 to 2016 was defined based on data from the Global Health Observatory and the India Demographic and Health Survey datasets (International Institute for Population Sciences, 2022; World Health Organization, 2025). Flows between BMI strata were not dynamically modelled in the framework.

##### 5.2 STRUCTURE

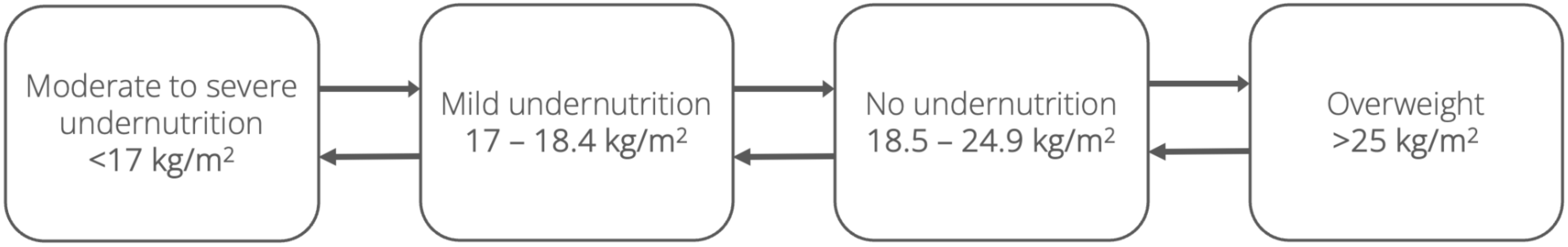

Figure S13. Nutrition dimension structure

#### 5.3 INTERACTION BETWEEN NUTRITION AND TB DIMENSION

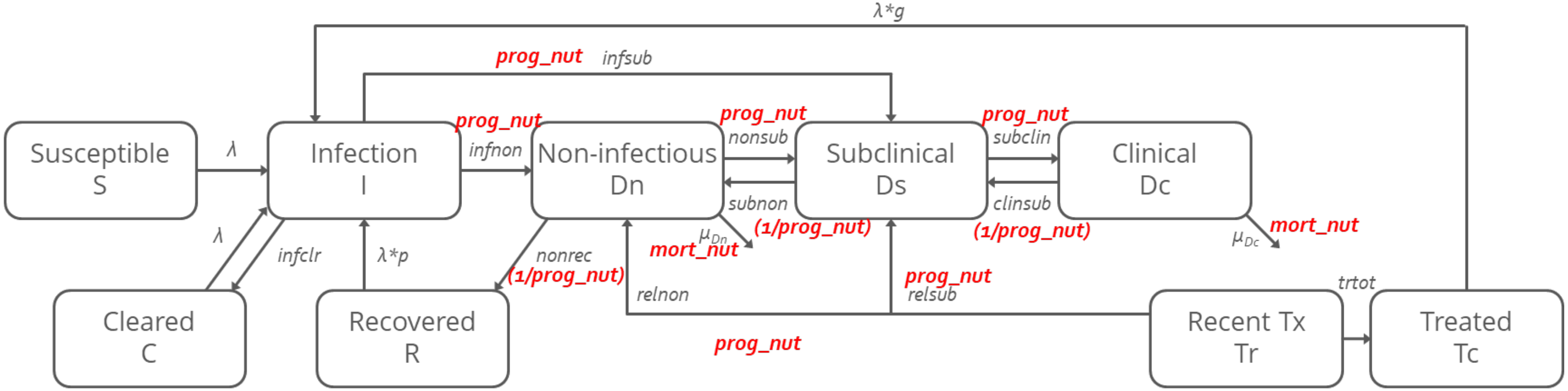

Drug-susceptible treatment initiation = treatment initiation rate * (1 – access to dst * probability of a positive dst)
Drug-resistant treatment initiation = treatment initiation rate * access to dst * probability of a positive dst
Successful treatment = completion rate * treatment success***tx_nut**
Unsuccessful treatment = completion rate * (1 – treatment success***tx_nut** – mortality on treatment ***mort_nut**)
Mortality on treatment = completion rate * mortality on treatment***mort_nut**

Figure S14. Nutrition adjustments to transitions in the TB dimension. TB disease state and parameter naming follow conventions from research over the past decade; results are reported using current WHO terminology.

- We assumed risk of progression to TB disease (*prog_nut*), treatment outcomes (*unsucc_nut*/*tx_nut*), and TB mortality (*mort_nut*) varied by BMI status. Adjustments are made by multiplying a given progression parameter that determines flow (e.g. *nonsub*) with the relative risks (*prog_nut*, relevant for nonsub, *tx_nut* or *mort_nut*, as applicable) experienced by people in particular BMI strata.

#### 5.4 EQUATIONS

Below shows the equations used in the risk dimension: Undernutrition in India. On treatment equations are not included, due to there being no differences from core in section 1.4. Differences from core are noted using orange.

##### 5.4.1 EXPRESSIONS

**lambda_all =** cscal * pT * k * C • (t*(Ds_s_N + Ds_s_P) + (Dc_s_N +Dc_s_P) + (relfit *( t*(Ds_s_N + Ds_s_P) + (Dc_s_N +Dc_s_P))))

**lambda_s =** cscal * pT * k * C • (t*(Ds_s_N + Ds_s_P) + (Dc_s_N +Dc_s_P))

**lambda_r =** cscal * pT * k * C • (relfit * (t*(Ds_s_N + Ds_s_P) + (Dc_s_N +Dc_s_P)))

**treat_init =** relinit * tinit * eta * dx_age

##### 5.4.2 NUTRITION DIMENSION EQUATIONS

**dover/dt =** NA

**dnormal/dt =** NA

**dmild/dt =** NA

**dmoderate/dt =** NA

##### 5.4.3 NEVER TREATED

**dS_N/dt =** - (treat_init + lambda_all) * S_N

**dC_N/dt =** infclr * (I_s_N + I_r_N) - (treat_init + lambda_all) * C_N

**dR_N/dt =** nonrec *(1/prog_age) * (1/prog_nut)* ( Dn_s_N + Dn_r_N ) – (treat_init + lambda_all * p) * R_N

**dI_s_N/dt =** lambda_s * (S_N + C_N + p*R_N) – (treat_init + infclr + lambda_r + (infnon + infsub * prog_nut) * prog_age) * I_s_N

**dI_r_N/dt =** lambda_r * (S_N + C_N + p*R_N + I_s_N) – (treat_init + infclr + prog_age * (infnon+ infsub * prog_nut ) * I_r_N

**dDn_s_N/dt =** infnon * prog_age * I_s_N + subnon * (1/prog_age) * (1/prog_nut) * Ds_s_N – (treat_init +nonrec * (1/prog_age) * (1/prog_nut) + nonsub * prog_age * prog_nut + muDn * mort_age * mort_nut) * Dn_s_N

**dDn_r_N/dt =** infnon*prog_age*I_r_N + subnon*(1/prog_age)* (1/prog_nut)*Ds_r_N – (treat_init + nonrec*(1/prog_age)* (1/prog_nut) + nonsub*prog_age* prog_nut + muDn * mort_age * mort_nut) * Dn_r_N

**dDs_s_N/dt =** prog_age* prog_nut*( infsub*I_s_N + nonsub *Dn_s_N – subclin * Ds_s_N) + (1/prog_age)* [(1/prog_nut)*( clinsub*Dc_s_N – subnon) - treat_init] * Ds_s_N

**dDs_r_N/dt =** prog_age* prog_nut*(infsub* I_r_N + nonsub* Dn_r_N – subclin * Ds_r_N )+ (1/prog_age)* [(1/prog_nut)*(clinsub*Dc_r_N – subnon) – treat_init ]*Ds_r_N)

**dDc_s_N/dt =** subclin*prog_age* prog_nut*Ds_s_N – (treat_init + (1/prog_nut)*clinsub*(1/prog_age) + mort_nut*muDc) * Dc_s_N

**dDc_r_N/dt =** subclin*prog_age* prog_nut*Ds_r_N – (treat_init + (1/prog_nut)*clinsub*(1/prog_age) + mort_nut*muDc )*Dc_r_N

##### 5.4.4 PREVIOUSLY TREATED

**dS_P/dt =** comp_st * ST_S + comp_rt * RT_S – (treat_init + lambda_all) * S_P

**dC_P/dt =** infclr*(I_s_P + I_r_P) + comp_st*(ST_C + succ_stI* (ST_I_s + succ_strs*ST_I_r)) + comp_rt*(RT_C + succ_rtI*(RT_I_s + RT_I_r)) – (treat_init + lambda_all) * C_P

**dR_P/dt =** nonrec*(1/prog_age)* (1/prog_nut)*(Dn_s_P + Dn_r_P) + comp_st*(ST_R + succ_stDn*(ST_Dn_s + succ_strs*ST_Dn_r)) + comp_rt*(RT_R + succ_rtDn*(RT_Dn_s + RT_Dn_r)) – (treat_init + lambda_all*p) * R_P

**dI_s_P/dt =** comp_st*(1-succ_stI*(1/unsucc_nut))*ST_I_s + comp_rt*(1-succ_rtI*(1/unsucc_nut))*RT_I_s + lambda_s*(S_P + C_P + p*R_P + g*Tc_P) – (infclr + treat_init + lambda_r + prog_age* (infnon + infsub* prog_nut)*I_s_P

**dI_r_P/dt =** comp_st*(1-succ_stI*succ_strs*(1/unsucc_nut))*ST_I_r + comp_rt*(1-succ_rtI*(1/unsucc_nut))*RT_I_r + lambda_r*(S_P + C_P + p*R_P + I_s_P + g*Tc_P) – (treat_init + infclr + prog_age*(infnon + infsub* prog_nut))*I_r_P

**dDn_s_P/dt =** prog_nut*infnon*prog_age*I_s_P + (1/prog_nut)*subnon*(1/prog_age)*Ds_s_P + comp_st*(1-mort_nut*mort_stDn-succ_stDn*(1/unsucc_nut))*ST_Dn_s + comp_rt*(1-mort_nut*mort_rtDn-succ_rtDn*(1/unsucc_nut))*RT_Dn_s + prog_nut*relnon_s*Tr_s_P - (nonrec*(1/prog_age)* (1/prog_nut) + nonsub*prog_age* prog_nut + muDn*mort_age* mort_nut + treat_init)*Dn_s_P

**dDn_r_P/dt =** prog_nut *infnon*prog_age*I_r_P + (1/prog_nut)*subnon*(1/prog_age)*Ds_r_P + comp_st*(1-mort_nut *mort_stDn-succ_stDn*succ_strs*(1/unsucc_nut))*ST_Dn_r + comp_rt*(1-mort_nut *mort_rtDn-succ_rtDn(1/unsucc_nut))*RT_Dn_r + prog_nut *relnon_r*Tr_r_P - (nonrec*(1/prog_age)* (1/prog_nut) + nonsub*prog_age* prog_nut + muDn*mort_age* mort_nut + treat_init)*Dn_r_P

**dDs_s_P/dt =** prog_nut *prog_age*(infsub *I_s_P + nonsub*Dn_s_P) + (1/prog_nut)*clinsub*(1/prog_age)*Dc_s_P + *comp_st*(1-succ_stDs*(1/unsucc_nut))*ST_Ds_s + comp_rt*(1-succ_rtDs*(1/unsucc_nut))*RT_Ds_s + prog_nut *relsub_s*Tr_s_P - ( (1/prog_nut)*subnon*(1/prog_age) + prog_nut *subclin*prog_age + treat_init)*Ds_s_P

**dDs_r_P/dt =** prog_nut *prog_age (*infsub *I_r_P + nonsub*Dn_r_P) + (1/prog_nut)*clinsub*(1/prog_age)*Dc_r_P + comp_st*(1-succ_stDs*succ_strs*(1/unsucc_nut))*ST_Ds_r + comp_rt*(1-succ_rtDs*(1/unsucc_nut))*RT_Ds_r + prog_nut *relsub_r*Tr_r_P - ( (1/prog_nut)*subnon*(1/prog_age) - prog_nut *subclin*prog_age+ treat_init) *Ds_r_P

**dDc_s_P/dt =** prog_nut *subclin*prog_age*Ds_s_P + comp_st*((1-(mort_stDc*mort_nut))*(1-(succ_stDc*(1/unsucc_nut)))*ST_Dc_s + comp_rt*((1-(mort_rtDc*mort_nut))*(1-(succ_rtDc*(1/unsucc_nut))))*RT_Dc_s - ( (1/prog_nut)*clinsub*(1/prog_age) + mort_nut *muDc + treat_init)*Dc_s_P

**dDc_r_P/dt =** prog_nut *subclin*prog_age*Ds_r_P + comp_st*((1-(mort_stDc*mort_nut))*(1-(succ_stDc*(1/unsucc_nut)*succ_strs)))*ST_Dc_r + comp_rt*((1-(mort_rtDc*mort_nut))*(1-(succ_rtDc*(1/unsucc_nut))))*RT_Dc_r - ( (1/prog_nut)*clinsub*(1/prog_age) + mort_nut *muDc + treat_init)*Dc_r_P

**dTr_s_P/dt =** comp_st*succ_stDs*(1/unsucc_nut)*ST_Ds_s + comp_rt*succ_rtDs*(1/unsucc_nut)*RT_Ds_s + comp_st*succ_stDc*(1/unsucc_nut)*ST_Dc_s + comp_rt*succ_rtDc*(1/unsucc_nut)*RT_Dc_s – (trtot + prog_nut *relnon_s + prog_nut *relsub_s)*Tr_s_P

**dTr_r_P/dt =** comp_st*succ_stDs*succ_strs*(1/unsucc_nut)*ST_Ds_r + comp_rt*succ_rtDs*(1/unsucc_nut)*RT_Ds_r + comp_st*succ_stDc*succ_strs*(1/unsucc_nut)*ST_Dc_r + comp_rt*succ_rtDc*(1/unsucc_nut)*RT_Dc_r – (trtot + prog_nut *relnon_r + prog_nut *relsub_r)*Tr_r_P

**dTc_P/dt =** comp_st*ST_Tc + comp_rt*RT_Tc + trtot*Tr_s_P + trtot*Tr_r_P – (treat_init + lambda_all*g)*Tc_P

#### 5.5 PARAMETERS

The relationship between BMI and TB incidence were replicated from the methods used by Clark et al., with minor revisions where necessary (Clark et al., 2025). The age-standardised proportion of the adult population >19 years within each strata was taken from the Global Health Observatory for the years 1975–2016 (World Health Organization, 2025). This consisted of the proportion of the population overweight (BMI ≥ 25.0 kg/m2) or underweight (BMI < 18.5 kg/m2), from which the proportion of normal weight was calculated. The proportion underweight was separated into moderate to severe underweigth and mild underweigth based on trends in the ratio of each in the India Demographic and Health Surveys from 2005–2006, 2015–2016 and 2019–2021, 2020–2022 weighted by the male:female ratio from UN population estimates and using the survey package in R. To estimate the proportion of children and adolescents in each BMI strata, we again used the Global Health Observatory (World Health Organization, 2025). We separated children aged 0–4 years into BMI strata based on standard deviations (SD) for weight-for-height of severe wasting (weight-for-height < -3SD), wasting (-3SD ≤ weight-for-height < - 2SD), normal (-2SD ≤ weight-for-height <

+2SD) and overweight (weight-for-height ≥ +2SD), aligning with our four BMI strata. We note that estimates were only available for normal, wasting and severe wasting for the years 1999, 2006, 2014, 2015 and 2017, while for overweight these were available for 1994–2019.

We re-estimated the relationship between BMI and TB incidence from Lönnroth et al. using updated data from Cegielski et al. and assuming a uniform distribution for BMI within each BMI strata (Cegielski et al., 2012; Lönnroth et al., 2010). We used this to fit a log-linear model with a dependent variable of TB incidence, an independent variable of BMI, and a categorical variable for the studies reported in Lönnroth et al., controlling for study-specific effects due to variation in study design and populations studied (Lönnroth et al., 2010).

Using the log-linear model, the BMI distributions from above, and estimated population-level TB incidence in 2020 (while 2005–2006 did not include primary sampling units in IPUMS and 1998–1999 did not include data for men), we estimated the expected TB incidence for each BMI strata weighted by the female:male ratio. We used this to calculate the risk ratio for TB in each strata compared to the strata with normal BMI, assuming that differences in risk of incident TB were a direct result of differences in rate of progression or reversion to TB disease. We estimated the mean and uncertainty intervals for this from 10,000 runs of the process.

Treatment outcome estimates were calculated using data collected from adults with microbiologically confirmed TB in five sites of the Regional Prospective Observational Research on Tuberculosis (RePORT) India consortium from 2015–2019.(Sinha et al., 2023) The primary exposure was BMI at treatment initiation and the outcome was a composite of death, treatment failure, and relapse/recurrence. We conducted multivariable analysis using Poisson regression, in which we used person-time as an offset to calculate adjusted incidence rate ratios (aIRR). We calculated person-time from the time of treatment initiation to the occurrence of the first mutually exclusive outcome of interest (failure/recurrence/death), loss to follow-up, or until right-censoring at 24 months of follow-up. In multivariable analysis, we included age, sex, and cough duration in the multivariable a priori and included other potential confounders based on prior literature that were significant at the p<0.2 level. The original publication calculated risk for different BMI categories than the ones required for this model. We recalculated the aIRR for the following BMI categories: moderately to severely thin (BMI < 17.0 kg/m$^2$), mildly thin (17.0 kg/m$^2$ ≤ BMI < 18.5 kg/m$^2$), normal (18.5 kg/m$^2$ ≤ BMI <25.0kg/m$^2$) and overweight to obese (BMI ≥ 25.0 kg/m$^2$). We assumed that the risk ratios for disease progression, reversion, and treatment outcomes were different by BMI strata, and were applied identically to adults, adolescents, and children.

The parameter table details the nutrition dimension parameters, which include nutrition and TB specific interaction parameters; categorised by prior/value, age and time dependency. References are given where applicable.

Table S9. Nutrition dimension parameters

| Parameter | Description | Prior/Value | Age-dependent? | Time-dependent? | Reference |
|---|---|---|---|---|---|
| Nutrition parameters | | | | | |
| Rates between<br>● Moderate-mild<br>● Over-normal<br>● Normal-over<br>● Normal-mild<br>● Mild-normal<br>● Mild-moderate | Rates of flow between BMI states per year (proportion of population moving from one state to the other) | Input from data | Yes | Yes | (International Institute for Population Sciences, 2022; World Health Organization, 2025) |
| prog_nut | Relative risk of TB progression | Moderate: 1.25- 1.84<br>Mild: 1.12-1.47<br>Normal: 1<br>Overweight: 0.23-0.66 | No | No | (International Institute for Population Sciences, 2022; McQuaid et al., 2025) |
| reg_nut | Relative risk of natural recovery | 1/prog_nut | No | No | Assumption |
| mort_nut | Relative risk of TB mortality | Moderate: 1.84–6.48<br>Mild: 1<br>Normal: 1<br>Overweight:1 | No | No | (McQuaid et al., 2025; Sinha et al., 2024) |
| unsucc_nut | Relative risk of treatment non-completion /unsuccessful treatment | Moderate: 1.09–2.31<br>Mild: 1<br>Normal: 1<br>Overweight: 1 | No | No | |
| tx_nut | Relative risk of treatment success and/or completion | 1/unsucc_nut | No | No | Assumption |

### 6. RISK DIMENSION: INCARCERATION IN BRAZIL

#### 6.1 OVERVIEW

There are three population risk strata that we are modelling TB transmission in: general population, currently incarcerated, and previously incarcerated. The model allows for movement and mixing between specific groups (described below).

- Prison: A correctional facility where individuals who are serving sentences, or awaiting trial are held.
- General population / community: Individuals who are not currently incarcerated, nor have been incarcerated within the last 7 years.
- Incarcerated persons: Individuals who are currently in prison. Incarcerated persons will have entered prison from the general population or from the previously incarcerated population.
- Previously incarcerated persons: Individuals who have been released from prison within the past 7 years. Previously incarcerated individuals will rejoin the general population after 7 years.
- Individuals can be incarcerated and released from prison at any stage of the TB pathway.
- To capture the higher risk of TB in prisons, currently incarcerated individuals will have an increased progression through disease states and a higher contact rate than the general population. The increased risk will be distributed equally between progression and contact rates to combat the uncertainty regarding the exact source of the elevated risk. Previously incarcerated individuals will have equal progression to the currently incarcerated individuals to account for their increased progression compared to the general population.

#### 6.2 STRUCTURE

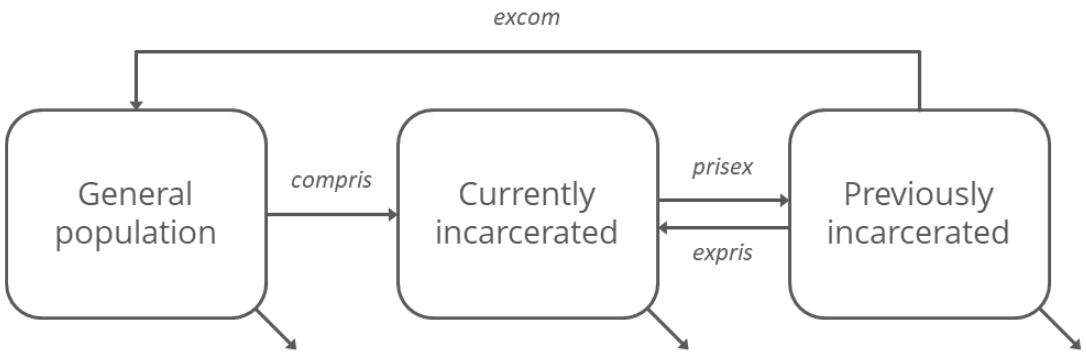

Figure S15. Prison dimension structure

#### 6.3 INTERACTION OF PRISONS AND TB DIMENSION

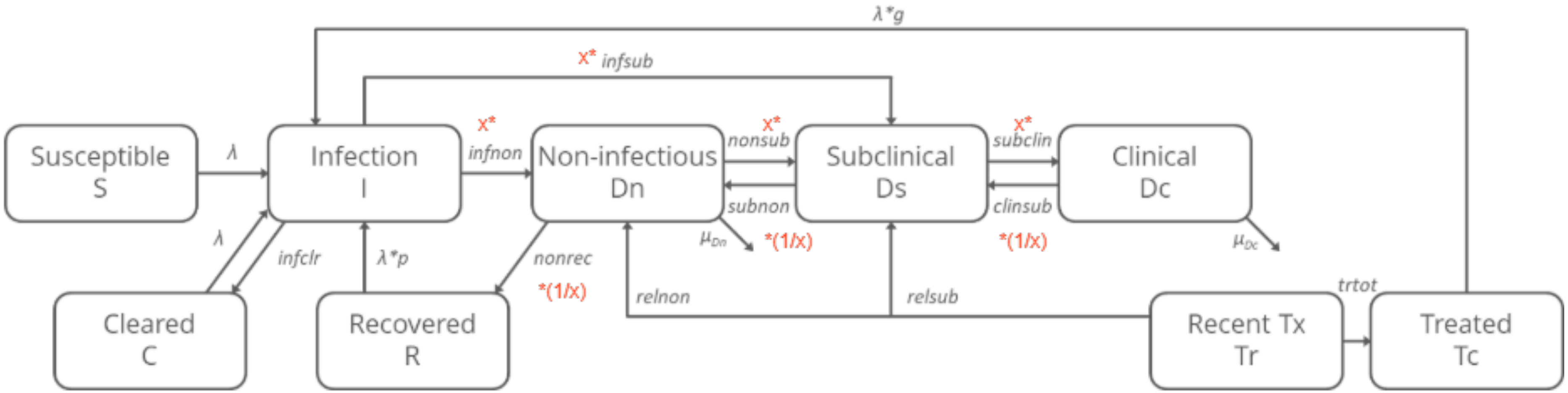

Figure 15. Prison adjustments to transitions in the TB dimension. The diagram shows an overview of which TB parameters are affected by incarceration. There are two types of parameters: those that increase the TB parameter and those that decrease the value of the TB parameter .Detailed treatment pathways are shown in Figure 4 and Figure 5.
X = prog_not_inprison+inprison*prog_inprison*prog_contRate_pri

#### 6.4 EQUATIONS

Below shows the equations used in the risk dimension: Incarceration in Brazil. On treatment equations are not included, due to there being no differences from core in section 1.4. Differences from core are noted using yellow.

##### 6.4.1 EXPRESSIONS

**lambda_all =** cscal * $C_{commexpris}$ * pT * mix_pri_cx * k * ((t*(Ds_s_N + Ds_s_P) + (Dc_s_N +Dc_s_P) + (relfit *( t*(Ds_r_N + Ds_r_P) + (Dc_r_N +Dc_r_P)))) + contactRate_pri_p * $C_{prison}$ * pT * mix_pri_p * k * ((t*(Ds_s_N + Ds_s_P) + (Dc_s_N +Dc_s_P) + (relfit *( t*(Ds_r_N + Ds_r_P) + (Dc_r_N +Dc_r_P)))/N)

**lambda_s_communityexpris =** cscal * $C_{commexpris}$ * pT * mix_pri_cx*k * ((t*(Ds_s_N + Ds_s_P) + (Dc_s_N +Dc_s_P)/N)

**lambda_r_communityexpris =** cscal * $C_{commexpris}$ * pT * mix_pri_cx* k * (relfit * (t*(Ds_r_N + Ds_r_P) + (Dc_r_N +Dc_r_P))/N)

**lambda_s_prison =** contactRate_pri_p * $C_{prison}$ * pT * mix_pri_cx*k * ((t*(Ds_s_N + Ds_s_P) + (Dc_s_N +Dc_s_P)/N)

**lambda_r_prison =** contactRate_pri_p * $C_{prison}$ * pT * mix_pri_cx* k * (relfit * (t*(Ds_r_N + Ds_r_P) + (Dc_r_N +Dc_r_P))/N)

treat_init = relinit*tinit * eta* dx_age

##### 6.4.2 PRISON DIMENSION EQUATIONS

**dcommunity/dt =** excom * exprsn - 1 * f * community

**dexprsn/dt =** exmortmult * 0.01696 * exprsn + 1 * prison - (excom + expris) * exprsn

**dprison/dt =** + expris*exprsn + 1*f*community - 1*prison

##### 6.4.3 NEVER TREATED

**dS_N/dt =** - (treat_init + lamda_all)*S_N

**dC_N/dt =** infclr*(I_s_N + I_r_N) - (treat_init + lambda_all)*C_N

**dR_N/dt =** (nonrec*(1/prog_age)*(1/(prog_not_inprison+inprison*prog_inprison*prog_contRate_pri ))** (Dn_s_N + Dn_r_N) - (treat_init + lambda_all*p)*R_N

**dI_s_N/dt =** lambda_s*(S_N + C_N + p*R_N) - infclr*I_s_N -
infnon*prog_age*(prog_not_inprison+inprison*prog_inprison*prog_contRate_pri)*I_s_N -
infsub*prog_age*(prog_not_inprison+inprison*prog_inprison*prog_contRate_pri)*I_s_N - (treat_init + lambda_r) *I_s_N

**dI_r_N/dt =** lambda_r*(S_N + C_N + p*R_N + I_s_N) - infclr*I_r_N -
infnon*prog_age*(prog_not_inprison+inprison*prog_inprison*prog_contRate_pri)*I_r_N -
infsub*prog_age*(prog_not_inprison+inprison*prog_inprison*prog_contRate_pri)*I_r_N - treat_init *I_r_N

**dDn_s_N/dt =** infnon*prog_age*(prog_not_inprison+inprison*prog_inprison*prog_contRate_pri)*I_s_N +
subnon*(1/prog_age)*(1/(prog_not_inprison+inprison*prog_inprison*prog_contRate_pri ))**Ds_s_N - nonrec*(1/prog_age)
*(1/(prog_not_inprison+inprison*prog_inprison*prog_contRate_pri ))**Dn_s_N -
nonsub*prog_age*(prog_not_inprison+inprison*prog_inprison*prog_contRate_pri)*Dn_s_N - muDn*mort_age*Dn_s_N -
treat_init*Dn_s_N

**dDn_r_N/dt =** infnon*prog_age*(prog_not_inprison+inprison*prog_inprison*prog_contRate_pri)*I_r_N +
subnon*(1/prog_age)*(1/(prog_not_inprison+inprison*prog_inprison*prog_contRate_pri ))**Ds_r_N - nonrec*(1/prog_age)
*(1/(prog_not_inprison+inprison*prog_inprison*prog_contRate_pri ))** Dn_r_N -
nonsub*prog_age*(prog_not_inprison+inprison*prog_inprison*prog_contRate_pri)*Dn_r_N - muDn*mort_age*Dn_r_N - treat_init
*Dn_r_N

**dDs_s_N/dt =** infsub*prog_age*(prog_not_inprison+inprison*prog_inprison*prog_contRate_pri)*I_s_N +
nonsub*prog_age*(prog_not_inprison+inprison*prog_inprison*prog_contRate_pri)*Dn_s_N +
clinsub*(1/prog_age)*(1/(prog_not_inprison+inprison*prog_inprison*prog_contRate_pri ))**Dc_s_N -
subnon*(1/prog_age)*(1/(prog_not_inprison+inprison*prog_inprison*prog_contRate_pri ))**Ds_s_N -
subclin*prog_age*(prog_not_inprison+inprison*prog_inprison*prog_contRate_pri)*Ds_s_N - treat_init *Ds_s_N

**dDs_r_N/dt =** infsub*prog_age*(prog_not_inprison+inprison*prog_inprison*prog_contRate_pri)*I_r_N +
nonsub*prog_age*(prog_not_inprison+inprison*prog_inprison*prog_contRate_pri)*Dn_r_N +
clinsub*(1/prog_age)*(1/(prog_not_inprison+inprison*prog_inprison*prog_contRate_pri ))**Dc_r_N -
subnon*(1/prog_age)*(1/(prog_not_inprison+inprison*prog_inprison*prog_contRate_pri ))**Ds_r_N -
subclin*prog_age*(prog_not_inprison+inprison*prog_inprison*prog_contRate_pri)*Ds_r_N - treat_init *Ds_r_N

**dDc_s_N/dt =** subclin*prog_age*(prog_not_inprison+inprison*prog_inprison*prog_contRate_pri)*Ds_s_N - clinsub*(1/prog_age)*(1/(prog_not_inprison+inprison*prog_inprison*prog_contRate_pri ))**Dc_s_N - muDc*Dc_s_N - treat_init *Dc_s_N

**dDc_r_N/dt =** subclin*prog_age*(prog_not_inprison+inprison*prog_inprison*prog_contRate_pri)*Ds_r_N - clinsub*(1/prog_age)*(1/(prog_not_inprison+inprison*prog_inprison*prog_contRate_pri ))**Dc_r_N - muDc*Dc_r_N - treat_init *Dc_r_N

##### 6.4.4 PREVIOUSLY TREATED

**dS_P/dt =** comp_st*ST_S + comp_rt*RT_S - (treat_init + lambda_all)*S_P

**dC_P/dt =** infclr*(I_s_P + I_r_P) + comp_st*(ST_C + succ_stI*(ST_I_s + succ_strs*ST_I_r)) + comp_rt*(RT_C + succ_rtI*(RT_I_s + RT_I_r)) - (treat_init + lambda_all)*C_P

**dR_P/dt =** nonrec*(1/prog_age)*(1/(prog_not_inprison+inprison*prog_inprison*prog_contRate_pri ))** (Dn_s_P + Dn_r_P) + comp_st*(ST_R + succ_stDn*(ST_Dn_s + succ_strs*ST_Dn_r)) + comp_rt*(RT_R + succ_rtDn*(RT_Dn_s + RT_Dn_r)) - (treat_init + lambda_all*p)*R_P

**dI_s_P/dt =** comp_st*(1-succ_stI)*ST_I_s + comp_rt*(1-succ_rtI)*RT_I_s + lambda_s*(S_P + C_P + p*R_P + r*Tc_P) - (infclr+ infnon*prog_age*(prog_not_inprison+inprison*prog_inprison*prog_contRate_pri))*I_s_P - infsub*prog_age*(prog_not_inprison+inprison*prog_inprison*prog_contRate_pri)*I_s_P - (treat_init +lambda_r)*I_s_P

**dI_r_P/dt =** comp_st*(1-succ_stI*succ_strs)*ST_I_r + comp_rt*(1-succ_rtI)*RT_I_r + lambda_r*(S_P + C_P + p*R_P + I_s_P + r*Tc_P) - infnon*prog_age*(prog_not_inprison+inprison*prog_inprison*prog_contRate_pri)*I_r_P - infsub*prog_age*(prog_not_inprison+inprison*prog_inprison*prog_contRate_pri)*I_r_P - treat_init *I_r_P

**dDn_s_P/dt =** infnon*prog_age*(prog_not_inprison+inprison*prog_inprison*prog_contRate_pri)*I_s_P + subnon*(1/prog_age)*(1/(prog_not_inprison+inprison*prog_inprison*prog_contRate_pri ))**Ds_s_P + comp_st*(((1-mort_stDn)*mort_age*(1-succ_stDn))+((1-mort_age)*(1-((1+succ_stDn)/2))))*ST_Dn_s + comp_rt*((1-mort_rtDn)*mort_age*(1-

succ_rtDn))+((1-mort_age)*(1-((1+succ_rtDn)/2)))*RT_Dn_s + relnon_s*Tr_s_P - nonrec*(1/prog_age)*(1/(prog_not_inprison+inprison*prog_inprison*prog_contRate_pri ))**Dn_s_P - nonsub*prog_age*(prog_not_inprison+inprison*prog_inprison*prog_contRate_pri)*Dn_s_P - muDn*mort_age*Dn_s_P- treat_init*Dn_s_P

**dDn_r_P/dt =** prog_age*(prog_not_inprison+inprison*prog_inprison*prog_contRate_pri)*(infnon*I_r_P - nonsub*Dn_r_P) + subnon*(1/prog_age)*(1/(prog_not_inprison+inprison*prog_inprison*prog_contRate_pri ))**Ds_r_P + comp_st*(((1-mort_stDn)*mort_age*((1-succ_stDn)*succ_strs))+((1-mort_age)*(1-(((1+succ_stDn)/2)*succ_strs)))*ST_Dn_r + comp_rt*(((1-mort_rtDn)*mort_age*(1-succ_rtDn))+((1-mort_age)*(1-((1+succ_rtDn)/2)))*RT_Dn_r + relnon_r*Tr_r_P - nonrec*(1/prog_age)*(1/(prog_not_inprison+inprison*prog_inprison*prog_contRate_pri ))**Dn_r_P - muDn*mort_age*Dn_r_P - treat_init *Dn_r_P

**dDs_s_P/dt =** prog_age*(prog_not_inprison+inprison*prog_inprison*prog_contRate_pri)*(infsub*I_s_P + nonsub*Dn_s_P - subclin*Ds_s_P) + (1/prog_age)*(1/(prog_not_inprison+inprison*prog_inprison*prog_contRate_pri ))** (clinsub*Dc_s_P - subnon*Ds_s_P) + comp_st*(1-mort_stDc)*(1-succ_stDc)*ST_Ds_s + comp_rt*(1-succ_rtDs)*RT_Ds_s + relsub_s*Tr_s_P -treat_init*Ds_s_P

**dDs_r_P/dt =** prog_age*(prog_not_inprison+inprison*prog_inprison*prog_contRate_pri)* (infsub*I_r_P + nonsub*Dn_r_P - subclin*Ds_r_P) + (1/prog_age)*(1/(prog_not_inprison+inprison*prog_inprison*prog_contRate_pri ))** (clinsub*Dc_r_P - subnon*Ds_r_P) + comp_st*(1-succ_stDs*succ_strs)*ST_Ds_r + comp_rt*(1-succ_rtDs)*RT_Ds_r + relsub_r* - treat_init *Ds_r_P

**dDc_s_P/dt =** subclin*prog_age*(prog_not_inprison+inprison*prog_inprison*prog_contRate_pri)*Ds_s_P + comp_st*(1-mort_stDc)*(1-succ_stDc)*ST_Dc_s + comp_rt*(1-mort_rtDc)*(1-succ_rtDc)*RT_Dc_s - clinsub*(1/prog_age)*(1/(prog_not_inprison+inprison*prog_inprison*prog_contRate_pri ))**Dc_s_P - (muDc + treat_init)*Dc_s_P

**dDc_r_P/dt =** subclin*prog_age*(prog_not_inprison+inprison*prog_inprison*prog_contRate_pri)*Ds_r_P + comp_st*(1-mort_stDc)*(1-succ_stDc*succ_strs)*ST_Dc_r + comp_rt*((1-mort_rtDc)*(1-succ_rtDc)*RT_Dc_r - clinsub*(1/prog_age)*(1/(prog_not_inprison+inprison*prog_inprison*prog_contRate_pri ))**Dc_r_P - muDc*Dc_r_P - treat_init *Dc_r_P

**dTr_s_P/dt =** comp_st*(succ_stDs*ST_Ds_s + succ_stDc*ST_Dc_s) + comp_rt*(succ_rtDs*RT_Ds_s + succ_rtDc*RT_Dc_s) - (trtot + relnon_s + relsub_s)*Tr_s_P

**dTr_r_P/dt =** comp_st*succ_strs*(succ_stDs*ST_Ds_r + succ_stDc*ST_Dc_r) + comp_rt*(succ_rtDs*RT_Ds_r + succ_rtDc*RT_Dc_r) - (trtot + relnon_r + relsub_r)*Tr_r_P

**dTc_P/dt =** comp_st*ST_Tc + comp_rt*RT_Tc + trtot*(Tr_s_P + Tr_r_P) - (treat_init + lambda_all*r)*Tc_P

#### 6.5 PARAMETERS

The parameter table details the incarceration dimension parameters, which include general incarceration and TB specific interaction parameters; categorised by prior/value, age and time dependency. References are given where applicable.

Table S10. Incarceration dimension parameters

| Parameter | Description (units) | Prior/Value | Age-dependent? | Time-dependent? | Reference |
|---|---|---|---|---|---|
| **Incarceration parameters** | | | | | |
| compris | Admission rate to prison from the general population (per year) | 0 | Yes: 0-14 | No | (Liu et al., 2024) |
| | | 0.00018 - 0.00054 | Yes: 15+ | Yes (from 1990) | |
| | | 0.00122 - 0.00294 | | Yes (from 2020) | |
| | | 0.00084 - 0.00261 | | Yes (from 2024) | |
| expris | Admission to prison from previously incarcerated (per year) | 0.126 - 0.325 | No | No | (Liu et al., 2024) |
| prisex | Release from prison (per year) | 0.479 - 1.201 | No | Yes (from 1990) | (Liu et al., 2024) |
| | | 0.452 - 1.064 | | Yes (from 2015) | |
| | | 0.476 - 1.106 | | Yes (from 2024) | |
| excom | Ex-prison to the community (per year) | 0.143 | No | No | (Liu et al., 2024) |
| exmortmult | Multiplier on background mortality to allow previously incarcerated to have a higher mortality rate | 0 - 0.3974 | No | No | (Liu et al., 2024) |
| **TB-incarceration interaction parameters** | | | | | |
| prog_not_inprison | TB progression multiplier. This works in conjunction with prog_inprison to ensure TB progression in the community is 1*progression | *Community = 1*<br>*Prison = 0*<br>*Exprsn = 0* | No | No | Assumption |

| | | | | | |
|---|---|---|---|---|---|
| prog_inprison | TB progression multiplier, applied to all populations. This is then multiplied by the inprison parameter to ensure this additional progression is applied only to previously incarcerated and incarcerated individuals | *1 - 2.5* | No | No | Assumption |
| 1/prog_inprison | Natural recovery multiplier applied to all populations. This is then multiplied by the inprison parameter to ensure this reduced regression is applied only to previously incarcerated and incarcerated individuals | *Inverse of TB progression* | No | No | Assumption |
| inprison | Multiplier applied to TB progression to ensure increased progression and regression only affects the previously incarcerated and incarcerated populations | *Community = 0*<br>*Prison = 1*<br>*Exprsn = 1* | No | No | Assumption |
| prog_contRate | Multiplier applied to progression and contact rate in prisons equally. This is to ensure progression and contact rate in prisons change by the same proportion to spread the risk equally. | *1 - 2.5* | No | No | Assumption |
| inf_r | Contact rate for prisons | Prison = ((-155*(prog_inprison^2))+(583*prog_inprison)-410)*prog_contRate_pri<br>Community = 1<br>Exprsn = 1 | No | No | Assumption |

- Both TB progression and natural recovery use the same parameter name because *prog_not_inprison* and *prog_inprison* are sampled and its inverse is used in the interaction with the respective TB parameter.
- Prison dimension specific parameters include admission and release from prison. Individuals can move into the prison strata from the general population rate *compris*. Individuals in the previously incarcerated stratus can either be reincarcerated or eventually move back to the general population strata at rates and *expris* and *excom* respectively
- For the prison dimension, the background mortality parameter (*mu*) will be varied to capture the different mortality risk across the three populations: increased mortality in the previously incarcerated population compared to the community and currently incarcerated populations.
- TB mortality is constant across all population strata.
- The probability of transmission pT is the same in and out of prisons. Increased TB incidence in prison is accounted for by the increased mixing through the contact rate *contrate_p* which varies during calibration, as well as a multiplier (*prog_inprison*) and (*prog_not_inprison*) which has been added to the TB progression cascade to indicate increased TB progression in prisoners and ex prisoners. Similarly, to indicate slower TB clearance/recovery (*reg_inprison = 1/prog_inprison*) and (*reg_not_inprison = 1/prog_not_inprison*). A detailed description of mixing in and out of prison is provided below.

Mixing

In the Brazil country model, there are three population strata: community, currently incarcerated individuals, and individuals previously incarcerated within the last seven years. The community comprises individuals not currently incarcerated, nor incarcerated within the last seven years. Community and previously incarcerated individuals are able to mix with each other and this mixing is age dependent, while incarcerated individuals can only mix with each other and this mixing is homogeneous.

The force of infection between community and previously incarcerated individuals is expressed by the following:

(Eq1) $\lambda = cscal * pT * \boldsymbol{C} \bullet \boldsymbol{inf}$

with ***C* • *inf*** the matrix product of the contact matrix ***C*** [with elements $c_{r,i,j}$ for risk group r and age groups *i* and *j*] and a vector ***inf*** [with elements $inf_{r,i} = I_{r,i}/N_r$, with I infectious individuals (Ds + Dc) of risk group *r* and age group *i* and $N_r$ the total number in risk group *r*]. Elements of both ***C*** and ***inf*** are ordered by risk group first and age group second (i.e. first 16 age groups for community and the 16 age groups for formerly incarcerated).

$$\begin{pmatrix} c_{comm,1,1} & \cdots & c_{comm,1,16} \\ \vdots & \ddots & \vdots \\ c_{comm,16,1} & \cdots & c_{comm,16,16} \\ c_{expris,1,1} & \cdots & c_{expris,1,16} \\ \vdots & \ddots & \vdots \\ c_{expris,16,1} & \cdots & c_{expris,16,16} \end{pmatrix} \begin{pmatrix} I_{comm,1}/N_{comm,1} \\ \vdots \\ I_{comm,16}/N_{comm,16} \\ I_{expris,1}/N_{expris,1} \\ \vdots \\ I_{expris,16}/N_{expris,16} \end{pmatrix} = \begin{pmatrix} c_{comm,1,1}I_{comm,1}/N_{comm,1} + \dots + c_{comm,1,16}I_{comm,16}/N_{comm,16} \\ \vdots \\ c_{comm,16,1}I_{comm,1}/N_{comm,1} + \dots + c_{comm,16,16}I_{comm,16}/N_{comm,16} \\ c_{expris,1,1}I_{expris,1}/N_{expris,1} + \dots + c_{expris,1,16}I_{expris,16}/N_{expris,16} \\ \vdots \\ c_{expris,16,1}I_{expris,1}/N_{expris,1} + \dots + c_{expris,16,16}I_{expris,16}/N_{expris,16} \end{pmatrix}$$

*cscal* is a contact scaling factor and will vary during calibration, while *pT* is the probability of transmission per respiratory contact between an infectious and healthy individual. *pT* has a fixed value of 0.0013 (0.008-0.0018) by the average number of social contacts per year (see note 1 in section 1.3).

The incarcerated population is modelled with homogenous mixing to reflect how interactions are unlikely age dependent, due to shared facilities, thereby assuming every inmate has an equal chance of interacting with another inmate. The incarcerated population can only mix with themselves. Contact with staff and the wider community has not been modelled for the incarcerated population because most interactions occur between incarcerated individuals, therefore external interactions would provide limited additional accuracy. The force of infection in prisons is estimated by the following:

(Eq2) $$\lambda_p = contrate_p * pT * I_p/N_p$$

whereby $contrate_p$ is the contact rate in prisons, $pT$ the probability of infection per contact, $I_p$ the infected population within prisons and $N_p$ the overall population in prisons. This reflects homogenous mixing.

### CALIBRATION

Calibration was performed using approximate Bayesian computation (ABC) Markov chain Monte Carlo (MCMC), seeded with manual calibrations, for each country. MCMC chains were run to at least 25,000 parameter sets and then thinned to select 500 parameter sets with posterior estimates consistent with all calibration targets.

#### 7 CALIBRATION TARGETS

Table S11. Calibration targets for each country

| Target | Description (units) | Year | Pop | Age | Country | | | Comments | Reference |
|---|---|---|---|---|---|---|---|---|---|
| | | | | | South Africa | India | Brazil | | |
| **TB targets** | | | | | | | | | |
| TB incidence | Estimated incidence (all forms) per 100 000 population [subclin*Ds / population] (with adjustment for double counting) | 2000 | All | All | 703 (387-1117) | 264 (90-527) | n/a | Symptomatic incidence is currently closest to WHO estimates. Targets were adjusted to not include extrapulmonary TB.<br><br>Can apply a standard reduction to account for repeat episodes within the same individual/infection if needed. | (World Health Organization, 2023) [e_inc_100k] |
| | | | HIV+ | All | 529 (271-870) | n/a | n/a | | (World Health Organization, 2023) [e_inc_tbhiv_100k] |
| | | 2007 | Prison | All | n/a | n/a | 1493 (1069-2118) | | (Liu et al., 2024) |
| | | 2010 | All | All | n/a | n/a | 40(34-46) | | (World Health Organization, 2023) [e_inc_100k] |
| | | 2019 | Prison | All | n/a | n/a | 1963 (1437-2677) | | (Liu et al., 2024) |
| | | 2022 | All | All | 432 (280-614) | 163 (139-189) | 44 (37-51) | | (World Health Organization, 2023) [e_inc_100k] |
| | | | | 0-14 | 146 (81-205) | 74 (62-88) | 10.5 (8.6-11.9) | | (United Nations, n.d.; World Health Organization, 2023) |

| | | | | | | | | | |
|---|---|---|---|---|---|---|---|---|---|
| | | | | | | | | | [estimates disaggregated by age [best] / UN population (0-14)] |
| | | | | 15+ | 551 (316-783) | 194 (161-228) | 53 (45-61) | | (United Nations, n.d.; World Health Organization, 2023) [estimates disaggregated by age [best] / UN population (15-99)] |
| | | | HIV+ | All | 235 (153-334) | n/a | n/a | | (World Health Organization, 2023) [e_inc_tbhiv_100k] |
| TB mortality | Estimated mortality of TB cases (all forms) per 100 000 population [(muDn*Dn + muDc*Dc) / population] | 2000 | All | All | 317 (199-461) | n/a | n/a | | (World Health Organization, 2023) [e_mort_100k] |
| | | | HIV+ | All | 258 (144-405) | n/a | n/a | | (World Health Organization, 2023) [e_mort_tbhiv_100k] |
| | | 2010 | All | All | n/a | n/a | 4.2 (3.7-4.7) | | (World Health Organization, 2023) [e_mort_100k] |
| | | 2015 | All | All | n/a | 32 (25-40) | n/a | | (World Health Organization, 2023) [e_mort_100k] |
| | | 2019 | All | All | n/a | 25 (19 -31) | n/a | | (World Health Organization, 2023) [e_mort_100k] |
| | | 2022 | All | All | 90 (51-142) | n/a | 6.2 (5.3-7.1) | | (World Health Organization, 2023) [e_mort_100k] |
| | | | HIV+ | All | 52 (17-107) | n/a | n/a | | (World Health Organization, 2023) [e_mort_tbhiv_100k] |
| TB notifications | Case notification rate, which is the total of new and relapse cases and | 2000 | All | All | 321 (257-385) | 177 (106.2-247) | n/a | Note values for access and diag differ across TB states. | (World Health Organization, 2023) [c_newinc_100k] |

| | | | | | | | | | |
|---|---|---|---|---|---|---|---|---|---|
| | cases with unknown previous TB treatment history per 100 000 population [tinit*relinit*(S+C+R+T+I+Dn+Ds+Dc) / population] | 2007 | Prison | All | n/a | n/a | 804 (643-965) | Notification rates for IND are adjusted by a reporting factor accounting for the mix in IND's public private TB diagnosis (Clark, Weerasuriya, et al., 2023) | (Liu et al., 2024) |
| | | 2010 | All | All | n/a | n/a | 39 (31.2-46.8) | | (World Health Organization, 2023) [c_newinc_100k] |
| | | 2019 | Prison | All | n/a | n/a | 1303 (1042-1564) | | (Liu et al., 2024) |
| | | 2022 | All | All | 344 (275-413) | 182(145-218) | 42 (33.6-50.4) | | (World Health Organization, 2023) [c_newinc_100k] |
| | | | All | 0-14 | 97 (77-116) | 42(34-50) | 6.3 (5-7.6) | | (World Health Organization, 2023) [newrel_m014 + newrel_f014] / UN Population (0-14) |
| | | | All | 15+ | 464 (372-558) | 232 (186-239) | 50.4 (40.3-60.5) | | (World Health Organization, 2023) [newrel_m15plus + newrel_f15plus] / UN Population (15-99) |
| | | | HIV+ | All | 171 (137-206) | n/a | n/a | | (World Health Organization, 2023) [newrel_hivpos] / UN Population (0-99) |
| TB prevalence | Prevalence of bacteriologically confirmed pulmonary TB per 100 000 population aged 15 years or over [((Ds+Dc) / N)] | 2019 | All | All | 680 (535-821) | n/a | n/a | Survey cases on treatment: 12.0% in India, 4.3% in South Africa. Targets were adjusted to not include them. India's targets were also adjusted to not include extrapulmonary TB.<br><br>No data for Brazil | (World Health Organization, 2023) National survey reports |
| | | 2015 | All | All | n/a | 227 (152-381) | n/a | | |
| | | 2021 | All | All | n/a | 225 (157-292) | n/a | | |
| | Proportion of infectious TB prevalence that is asymptomatic | 2019 | All | 15+ | 57.7 (51.4-64.0) | n/a | n/a | | |
| | | 2021 | All | 15+ | n/a | 50.4 (36.1-79.7) | n/a | | |

| | | | | | | | | | |
|---|---|---|---|---|---|---|---|---|---|
| | [Ds / (Ds+Dc * 100)] | | | | | | | | |
| RR-TB | Estimated percentage of TB notifications with rifampicin resistant TB | 2022 | All | All | 3.9 (3.12-4.68) | 4 (3.2 - 4.8) | 2.7 (0.7-7.7) | Confidence intervals were calculated as +/- 20% of value for ZAF and IND. Ranges for BRA were much larger so lower and upper bound estimates for new and previously treated were weighted by the proportion of cases that were new and previously treated. | (World Health Organization, 2023) |
| **HIV targets** | | | | | | | | | |
| HIV prevalence | % | 2000 | All | 0-14 | 0.9 (0.5-1.2) | n/a | n/a | | (Joint United Nations Programme on HIV/AIDS, 2021) |
| | | | All | 15+ | 10.1 (6.1-13.8) | n/a | n/a | | |
| | | 2022 | All | 0-14 | 1.4 (0.8-3.2) | n/a | n/a | | (Joint United Nations Programme on HIV/AIDS, 2023) |
| | | | All | 15+ | 16.1 (11.3-20.4) | n/a | n/a | | |
| Percentage of PLHIV who are on ART | % | 2010 | All | 0-14 | 24 (14-34) | n/a | n/a | | (Joint United Nations Programme on HIV/AIDS, 2021) |
| | | | All | 15+ | 23 (15-31) | n/a | n/a | | |
| | | 2022 | All | 0-14 | 54 (34-100) | | | | (Joint United Nations Programme on HIV/AIDS, 2023) |
| | | | All | 15+ | 75 (53-98) | n/a | n/a | | |
| HIV mortality rate | per 100,000 | 2000 | All | All | 289 (162-445) | n/a | n/a | | (Joint United Nations Programme on HIV/AIDS, 2021) |

| | | | | | | | | | |
|---|---|---|---|---|---|---|---|---|---|
| | | 2022 | All | All | 72 (50-144) | n/a | n/a | | (Joint United Nations Programme on HIV/AIDS, 2023) |
| **Nutrition targets** | | | | | | | | | |
| PAF | Prevalence of undernutrition and relative risk of TB due to undernutrition | 2022 | All | All | n/a | 45.2 (17.0-71.0) | n/a | | (Bhargava et al., 2022) |
| **Incarceration targets** | | | | | | | | | |
| Prison entry rate | Prison entry rates per 100k among population aged 15+ (years) | 2016 | All | 15+ | n/a | n/a | 338 (210-468) | | (Liu et al., 2024) |
| | | 2019 | All | 15+ | n/a | n/a | 442 (275-612) | | |
| | | 2022 | All | 15+ | n/a | n/a | 347 (216-480) | | |
| Prison prevalence | Incarceration prevalence per 100k among population aged 15+ (years) | 2000 | All | 15+ | n/a | n/a | 189 (151-227) | | (Liu et al., 2024) |
| | | 2019 | All | 15+ | n/a | n/a | 452 (362-542) | | |
| | | 2022 | All | 15+ | n/a | n/a | 488 (390-586) | | |
