## Supplementary materials 2 (S2) for "The potential impact, cost and cost-effectiveness of tuberculosis interventions - a modelling exercise"

1 - TB Modelling Group, TB Centre, LSHTM, London, UK; 2 - Department of Infectious Disease Epidemiology, LSHTM, London, UK; 3 - Instituto de Medicina Tropical Alexander von Humboldt, Universidad Peruana Cayetano Heredia, Lima, Peru; 4 - Global Health Economics Centre, LSHTM, London, UK; 5 - Department of Epidemiology, Biostatistics, and Occupational Health, School of Population and Global Health, McGill University, Montreal, QC, Canada; 6 – KNCV Tuberculosis Foundation, The Hague, Netherlands; 7 - SEICHE Center for Health and Justice, Yale University School of Medicine, New Haven, CT, USA; 8 - Justice Collaboratory, Yale Law School, New Haven, CT, USA; 9 - Health Economics and Epidemiology Research Office, Wits Health Consortium, Johannesburg, South Africa; 10 - French Institute for Research in Sustainable Development (IRD), Montpellier, France; 11 - CRDF Global, Arlington, VA, USA

**Correspondence:** Rein M.G.J Houben, Department of Infectious Disease Epidemiology, London School of Hygiene and Tropical Medicine, Keppel St, London, WC1E 7HT United Kingdom

**GitHub:** https://github.com/lshtm-tbmg/PACE-TB

### INTERVENTIONS

This document describes the TB interventions modelled, first describing the principles and then each intervention in turn, followed by the scale-up scenarios.

### 1. PRINCIPLES

When deciding what to model, we are looking for the following characteristics for each intervention category:

- *One intervention per category:* model a single variation of the same intervention (e.g., one algorithm for community screening)
    - Rationale: With three countries, multiple interventions with multiple outcomes, the number of results will be quite large already. Also, the aim of this exercise is to compare across intervention categories, not within.
- *Similar level of novelty:* for each intervention category that is at the forefront of current development, but ideally has an empirical estimate of potential epidemiological impact.
    - Rationale: We are looking to have a fair comparison across the interventions. If one intervention is more aspirational rather than supported by empirical evidence of epidemiological impact, this introduces an imbalance.
- *Same interventions in each country: the* format of the intervention and empirical impact are applied equally across countries.
    - Rationale: the reason for differences in outcomes (epi, CEA, budget) is driven by differences in the epidemiological baseline situation between Brazil, India and South Africa. An additional reason for between-country differences is baseline levels of overlapping interventions, such as treatment for DS-TB and DR-TB. This will be explored as well.
- *Same roll-out and scale up:* for all interventions, we will start scale-up in 2025, and reach 80% coverage of the target population by 2030 (i.e., a 5-year scale-up period).
    - Rationale: Alignment in roll-out and scale-up means that comparisons across the outcomes are more likely to be comparable.

### 2. INTERVENTIONS

We considered the following list of intervention categories, across four domains:

1. **Prevention**: TB vaccination, TB Preventative Treatment
2. **Screening and diagnosis**: Community screening, enhanced diagnostic in clinics, drug susceptibility testing for all
3. **Treatment**: Shortened DS-TB regimen, shortened DR-TB regimen
4. **Structural drivers**: Mass screening in prisons in Brazil, nutritional support for HHC of individuals starting TB treatment in India.

Note: We will also define a ‘business-as-usual’ (BAU) scenario for each country to provide a baseline to compare against.

### 2.1 BUSINESS-AS-USUAL:

The following tables present the BAU scenario for each country. These scenarios share some assumptions to enable clearer comparisons between BAU and intervention scenarios, and facilitate costing:

- All TB treatment (DS- and DR-) is at home with only monthly visits to the clinic: no inpatient stays, no DOTS.
- Adverse events caused by TB treatment are not taken into account (for costs or utility).
- BCG vaccination occurs at current rates and remains unchanged in all scenarios, and there are no additional vaccinations against TB other than in the Vaccination intervention.
- It is assumed that there is 0% TPT for child contacts at present, only TPT for people living with HIV in South Africa (in reality, there is also some TPT for child contacts of DS-TB).
- It is assumed that there is 0% nutrition support provided for people with TB and low BMI or their families.
- It is assumed that there is 0% routine community screening.
- Costs do not vary over time.
- Comorbidities (other than HIV in South Africa and malnutrition in India) are not considered in the model.
- Wider or longer-term health service expenditure for the subjects is not considered.

#### 2.1.1 BUSINESS-AS-USUAL: BRAZIL

The following table describes the intervention-specific changes and costs for BAU in Brazil.

| Aspect | Details |
|---|---|
| **Intervention summary** | |
| Intervention number, abbreviation, name | 1a. BAU (BRA) - Business-as-usual in Brazil |
| Summary description | Current diagnosis (clinical examination, some CXR and Xpert) and standard TB treatments in Brazil |
| Population affected | All (no changes for any group) |
| **Changes to the model** | |
| What is the effectiveness of the intervention? | N/A - No intervention |
| Which compartments are affected? | N/A - No intervention |
| Which transitions are affected? | N/A - No intervention |
| Which parameters are changed? From what value to what value? | N/A - No intervention |
| Any further details of changes | N/A - No intervention |
| **Costs** | |

| | | | | |
|---|---|---|---|---|
| What is being costed, and where are the costs applied? | **Activity** | **Resource use** | **Population** | **Model output** |
| | Standard TB assessment | **1x** appointment at a clinic<br>**35%** of patients x CXR<br>**50%** of patients x GeneXpert Ultra | Those entering the diagnostic pathway | n_assess_y[n] |
| | Standard DS-TB treatment | **7x** monthly appointments at a clinic<br>**6 months** of H+R+Z+E | Those entering treatment for DS-TB | n_treat_DS_y[n] |
| | Standard DR-TB treatment | **19x** monthly appointments at a clinic<br>**18 months** of R+Lfx+Z+E | Those entering treatment for DR-TB | n_treat_DR_y[n] |
| Do costs vary by age? | No | | | |
| **Implementation / Scale-up** | | | | |
| Duration of scale-up | N/A - No intervention | | | |
| What happens after implementation? | N/A - No intervention | | | |
| How is this implemented in the model? | N/A - No intervention | | | |
| How is this implemented in the costings? | N/A - No intervention | | | |
| **Assumptions** | | | | |
| Any other assumptions to note? | It is assumed that baseline diagnosis and treatment in prison are the same as in the community<br>The proportion of patients who received a CXR and GeneXpert were estimated using the Notifiable Diseases Information System (SINAN), a health information system that consolidates data and monitors | | | |

| | |
|---|---|
| | TB care and prevention across Brazil (Governo Brasileiro, n.d.).<br>- CXR: The precise number of patients receiving CXR is not known. The proportion of all TB patients that were clinically confirmed (2022) was 31% (28,326/91,808). 35% was chosen as a slightly higher value to reflect the fact that some other individuals will have had inconclusive CXR<br>- Xpert: % of patients that received Xpert (year 2022) over all cases: 50% (46,252/91,808) |

#### 2.1.2 BUSINESS-AS-USUAL: INDIA

The following table describes the intervention-specific changes and costs for BAU in India.

| **Aspect** | **Details** |
|---|---|
| **Intervention summary** | |
| Intervention number, abbreviation, name | 1b. BAU (IND) - Business-as-usual in India |
| Summary description | Current diagnosis (clinical examination, some CXR and Xpert) and standard TB treatments in India |
| Population affected | ALL (no changes for any group) |
| **Changes to the model** | |
| What is the effectiveness of the intervention? | N/A - No intervention |
| Which compartments are affected? | N/A - No intervention |
| Which transitions are affected? | N/A - No intervention |
| Which parameters are changed? From what value to what value? | N/A - No intervention |
| Any further details of changes | N/A - No intervention |
| **Costs** | |

<table>
<tr><td rowspan="4">What is being costed, and where are the costs applied?</td><td>Activity</td><td>Resource use</td><td>Population</td><td>Model output</td></tr>
<tr><td>Standard TB assessment</td><td>1x appointment at a clinic<br>10% of patients x CXR<br>21% of patients x GeneXpert Ultra</td><td>Those entering the diagnostic pathway</td><td>n_assess_y[n]</td></tr>
<tr><td>Standard DS-TB treatment</td><td>7x monthly appointments at a clinic<br>6 months of H+R+Z+E</td><td>Those entering treatment for DS-TB</td><td>n_treat_DS_y[n]</td></tr>
<tr><td>Standard DR-TB treatment</td><td>10x monthly appointments at a clinic<br>9 months of Lfx+Cfz+Z+E+Hh+Eto+Bdq</td><td>Those entering treatment for DR-TB</td><td>n_treat_DR_y[n]</td></tr>
<tr><td>Do costs vary by age?</td><td colspan="4">No</td></tr>
<tr><td colspan="5">Implementation / Scale-up</td></tr>
<tr><td>Duration of scale-up</td><td colspan="4">N/A - No intervention</td></tr>
<tr><td>What happens after implementation?</td><td colspan="4">N/A - No intervention</td></tr>
<tr><td>How is this implemented in the model?</td><td colspan="4">N/A - No intervention</td></tr>
<tr><td>How is this implemented in the costings?</td><td colspan="4">N/A - No intervention</td></tr>
<tr><td colspan="5">Assumptions</td></tr>
<tr><td>Any other assumptions to note?</td><td colspan="4">The proportion of individuals receiving CXR was considered as the lower quartile of CXR tests used among the number of individuals diagnosed in 2022 (Central Tuberculosis Division, 2023)<br>The proportion of individuals receiving Xpert was considered as the % of NAATs among those tested in</td></tr>
</table>

| | |
|---|---|
| | an examination for presumptive TB in 2023 (Central Tuberculosis Division, 2024). |

#### 2.1.3 BUSINESS-AS-USUAL: SOUTH AFRICA

The following table describes the intervention-specific changes and costs for BAU in South Africa.

| Aspect | Details |
|---|---|
| **Intervention summary** | |
| Intervention number, abbreviation, name | 1c. BAU (ZAF) - Business-as-usual in South Africa |
| Summary description | Current diagnosis (clinical examination, some CXR and Xpert) and standard TB treatments in South Africa |
| Population affected | All (no changes for any group) |
| **Changes to the model** | |
| What is the effectiveness of the intervention? | N/A - No intervention |
| Which compartments are affected? | N/A - No intervention |
| Which transitions are affected? | N/A - No intervention |
| Which parameters are changed? From what value to what value? | N/A - No intervention |
| Any further details of changes | N/A - No intervention |
| **Costs** | |

<table>
<tr><td rowspan="4">What is being costed, and where are the costs applied?</td><td>Activity</td><td>Resource use</td><td>Population</td><td>Model output</td></tr>
<tr><td>Standard TB assessment</td><td>1x appointment at a clinic<br>3.12% of patients x CXR<br>61% of patients x GeneXpert Ultra</td><td>Those entering the diagnostic pathway</td><td>n_assess_y[n]</td></tr>
<tr><td>Standard DS-TB treatment</td><td>7x monthly appointments at a clinic<br>6 months of H+R+Z+E</td><td>Those entering treatment for DS-TB</td><td>n_treat_DS_y[n]</td></tr>
<tr><td>Standard DR-TB treatment</td><td>10x monthly appointments at a clinic<br>9 months of Lfx+Lzd+Cfz+Z+E+Hh+Bdq</td><td>Those entering treatment for DR-TB</td><td>n_treat_DR_y[n]</td></tr>
<tr><td>Do costs vary by age?</td><td colspan="4">No</td></tr>
<tr><td colspan="5">Implementation / Scale-up</td></tr>
<tr><td>Duration of scale-up</td><td colspan="4">N/A - No intervention</td></tr>
<tr><td>What happens after implementation?</td><td colspan="4">N/A - No intervention</td></tr>
<tr><td>How is this implemented in the model?</td><td colspan="4">N/A - No intervention</td></tr>
<tr><td>How is this implemented in the costings?</td><td colspan="4">N/A - No intervention</td></tr>
<tr><td colspan="5">Assumptions</td></tr>
<tr><td>Any other assumptions to note?</td><td colspan="4">It is assumed that people living with HIV receive TPT for DS-TB in all scenarios (and so this is not modelled). TPT for DR-TB in PLHIV is not investigated in this model.<br>Proportion of individuals receiving Xpert was calculated from WHO data (World Health Organization,</td></tr>
</table>

| | |
|---|---|
| | 2024a), which suggested that 61% of the diagnoses were bacteriologically confirmed. Data from South Africa suggested that more than 90% of the tests conducted in people suspected of TB were Xpert (National Department of Health, Republic of South Africa, 2023). Therefore, it was assumed that everyone with a bacteriological diagnosis in South Africa would have received an Xpert test. Proportion of individuals receiving a CXR was calculated as the number of CXRs performed divided by the number of Xpert tests performed (National Department of Health, Republic of South Africa, 2023) |

### 2.2 VACCINATION:

The following table describes the specific changes and costs for TB vaccination across all three settings.

| Aspect | Details |
|---|---|
| **Intervention summary** | |
| Intervention number, abbreviation, name | 2. VAX - TB vaccination |
| Summary description | Annually vaccinate current 15-year-olds with 2 doses of a new M72-like TB vaccine.<br>Catch-up campaign for 16+ year-olds over five years, repeated after 10 years |
| Population affected | All 15-year-olds (assumed as 20% of the 15-19 age group)<br>All 16+ year-olds (assumed as 80% of the 15-19 age group, and all of the 20+ age groups) |
| **Changes to the model** | |
| What is the effectiveness of the intervention? | 50% reduction of progression to infectious disease (Tait et al., 2019) |
| Which compartments are affected? | Infected (I_s_N, I_r_N, I_s_P, I_r_P), non-infectious TB (Dn_s_N, Dn_r_N, Dn_s_P, Dn_r_P), and recently treated (Tr_s_P, Tr_r_P) compartments |
| Which transitions are affected? | Progression to infectious TB |
| Which parameters are changed? From what value to what value? | *infsub, nonsub* and *relsub* were reduced by 50% in individuals who received vaccination |
| Any further details of changes | Loss of vaccine efficacy was assumed to occur eventually after 10 years (rate = 0.1). Individuals who were vaccinated and progressed to infectious TB anyway were assumed to immediately lose that protection from vaccines (rate = 10). |
| **Costs** | |

<table>
<tr><td rowspan="6">What is being costed, and where are the costs applied?</td><td>Activity</td><td>Resource use</td><td>Population</td><td>Model output</td></tr>
<tr><td>Standard TB assessment</td><td>As BAU (varies by country)</td><td>Those entering the diagnostic pathway</td><td>n_assess_y[n]</td></tr>
<tr><td>Standard DS-TB treatment</td><td>As BAU (6 months of H+R+Z+E)</td><td>Those entering treatment for DS-TB</td><td>n_treat_DS_y[n]</td></tr>
<tr><td>Standard DR-TB treatment</td><td>As BAU (varies by country)</td><td>Those entering treatment for DR-TB</td><td>n_treat_DR_y[n]</td></tr>
<tr><td>Routine vaccination</td><td>Annually:<br>2x vaccine dose (including 5% wastage)<br>2x vaccination (including supply costs)</td><td>15-year-olds</td><td>n_vaccinated_y[n] (also includes below)</td></tr>
<tr><td>Catch-up vaccination</td><td>At scale-up and repeated after 10 years:<br>2x vaccine dose (including 5% wastage)<br>2x vaccination (including supply costs)</td><td>16+ year-olds</td><td>n_vaccinated_y[n] (also includes above)</td></tr>
<tr><td>Do costs vary by age?</td><td colspan="4">No</td></tr>
<tr><td colspan="5">Implementation / Scale-up</td></tr>
<tr><td>Duration of scale-up</td><td colspan="4">5 years</td></tr>
<tr><td>What happens after implementation?</td><td colspan="4">2030 - 2038: 80% of 15-year-olds vaccinated annually.<br>2039: one-year (re)vaccination campaign for all 15+ year olds.<br>2040 - 2050: 80% of 15-year-olds vaccinated annually.</td></tr>
<tr><td>How is this implemented in the model?</td><td colspan="4">Over each of the first 5 years, 16% of those of ages eligible for either initial or catch-up vaccination (15+ years) receive vaccination - and so by the end of 2029 the equivalent of 80% of 15+ year olds have been</td></tr>
</table>

| | |
|---|---|
| | vaccinated (noting that this will not be exactly 80% as the number of people 15+ will change with demographics as some will die or turn 15 during this period). This means that approximately the same number of vaccinations will be delivered in each of the first 5 years, not a gradual increase as in other interventions.<br>From 2030 - 2038, 80% of 15-year-olds are vaccinated each year - this is assumed as 16% of the 15-19 year-old age group.<br>In 2039, 80% of all 15+ year-olds are vaccinated<br>From 2040 - 2049, again, 16% of 15-19 year-olds are vaccinated |
| How is this implemented in the costings? | The model output n_vaccinated_y[n] counts the exact number vaccinated each year, regardless of age and incorporates scale-up, so the cost of vaccination is just multiplied by the number vaccinated. |
| **Assumptions** | |
| Any other assumptions to note? | Costs of introducing the vaccination programme in the first year ($2.40 per person, estimated by GAVI) are currently excluded from the analysis, to keep consistent with other interventions for which we do not have introduction costs. |

### 2.3 TB PREVENTIVE TREATMENT:

The following table describes the specific changes and costs for TB preventive treatment across all three settings.

| Aspect | Details |
|---|---|
| **Intervention summary** | |
| Intervention number, abbreviation, name | 3. TPT - TB preventive treatment |
| Summary description | Screen household members of individuals with asymptomatic or symptomatic TB diagnosed with TB (considered as bacteriologically diagnosed), and treat if diagnosed with TB. Give TPT to all children (<15 years old) without TB disease in households with an adult with TB (DS-TPT: 3HP, if index patient has DS-TB; DR-TPT: Lfx, if index patient has DR-TB). |
| Population affected | People newly diagnosed with and starting treatment for TB (both DS-TB and DR-TB). All household contacts of newly diagnosed asymptomatic or symptomatic TB on TB treatment. Household members of those newly diagnosed who are a) children <15 years old without TB disease, and b) children and adults with infectious TB. |
| **Changes to the model** | |
| What is the effectiveness of the intervention? | Reduction in progression among recently exposed individuals (DS-TPT: 64%; DR-TPT: 62%) (World Health Organization, 2024b) |
| Which compartments are affected? | Eligible for TPT: Children HHCs in susceptible (*S*), cleared (*C*), infection (*I*) and treated more than 1 year ago (*Tc*) compartments. Eligible for TB treatment: Adult or children HHCs with bacteriologically confirmed TB (*Ds* and *Dc*). |
| Which transitions are affected? | TB progression and the rate at which the household contacts start treatment are affected |
| Which parameters are changed? From what value to what value? | For TPT in children with no TB: TPT reduces the chances of progressing to TB by multiplying 0.36 in DS-TB TPT and 0.4 in DR-TB TPT to the following progression parameters: *infnon*, *infsub*, *nonsub*, and *subclin.* Children receiving TPT can not receive TB treatment. |

| | | | | |
|---|---|---|---|---|
| | For TB treatment in adults and children with infectious TB: We assumed a proportion of 3.4% with prevalent TB (Fox et al., 2013), which bypasses the usual treatment cascade and starts treatment through the following equation:<br>screening_coverage * HH_size_children/adults * proportion_prevalent_TB<br>N.B. The HHC starts DS-TB or DR-TB treatment based on their contact's treatment regimen. | | | |
| Any further details of changes | We used the following equations to represent the movement from no intervention to TPT:<br><br>- For *I*:<br>screening_coverage * HH_size_children * (1 - proportion_prevalent_TB) * proportion_completion_TPT * proportion_IGRApositive<br><br>- For *S*, *C*, *R* and *Tc*:<br>screening_coverage * HH_size_children * (1 - proportion_prevalent_TB) * proportion_completion_TPT * (1 - proportion_IGRApositive)<br><br>N.B. The *proportion_IGRApositive* was 35% (Martinez et al., 2020). | | | |
| **Costs** | | | | |
| What is being costed, and where are the costs applied? | **Activity** | **Resource use** | **Population** | **Model output** |
| | Standard TB assessment | As BAU (varies by country) | Those entering the diagnostic pathway | n_assess_y[n] |
| | Standard DS-TB treatment | As BAU (6 months of H+R+Z+E) | Those entering treatment for DS-TB | n_treat_DS_y[n] |
| | Standard DR-TB treatment | As BAU (varies by country) | Those entering treatment for DR-TB | n_treat_DR_y[n] |
| | Household contact assessment | **1x** appointment at a clinic<br>**1x** GeneXpert MTB/RIF Ultra | All household contacts of people newly diagnosed with | n_HHC_DS_y[n] +<br>n_HHC_DR_y[n] |

| | | | | |
|---|---|---|---|---|
| | | | DS-TB or DR-TB | |
| | TPT for DS-TB | **4x** monthly appointments at a clinic<br>**3 months** DS-TPT drugs (3HP) | Household contacts receiving TPT for DS-TB | n_HHC_DSTPT_y[n] |
| | TPT for DR-TB | **7x** monthly appointments at a clinic<br>**6 months** DR-TPT drugs (Lfx) | Household contacts receiving TPT for DR-TB | n_HHC_DRTPT_y[n] |
| Do costs vary by age? | No. TPT costs are based on a 25kg child (only children receive TPT). | | | |
| **Implementation / Scale-up** | | | | |
| Duration of scale-up | 5 years | | | |
| What happens after implementation? | Continues at 80% for the remaining 20 years | | | |
| How is this implemented in the model? | Contacts being assessed for TB and contacts receiving TPT: Straight line continuous increase from 0% at the start of 2025 to 80% at the end of 2029, then maintained at 80% (noting caveats under ‘Population affected’ above) | | | |
| How is this implemented in the costings? | Cost of assessment applied to 16% of total eligible populations (n_HHC_DS/DR_y) at the end of Y1, 32% at the end of Y2, 48% at the end of Y3, 64% at the end of Y4, and 80% in Y5 and in all remaining years.<br>n_HHC_DS/DRTPT_y outputs incorporate scale up (i.e. these outputs report the exact number of people receiving TPT each year), so the cost of TPT is just directly applied per person without further adjustment. | | | |
| **Assumptions** | | | | |
| Any other assumptions to note? | Adverse events caused by taking TPT are not included in the results, either in terms of loss of | | | |

| | |
|---|---|
| | utility/increased disability or in terms of the cost of additional medical care. |

### 2.4 NUTRITION:

The following table describes the specific changes and costs for nutrition in India.

| **Aspect** | **Details** |
|---|---|
| **Intervention summary** | |
| Intervention number, abbreviation, name | 4. NTN - Nutrition |
| Summary description | Provide nutritional support for 6 months to all those diagnosed with TB and their household members. |
| Population affected | All individuals starting TB treatment and their households. |
| **Changes to the model** | |
| What is the effectiveness of the intervention? | For individuals with TB: 33% reduced risk of on-treatment mortality (McQuaid et al., 2025).<br>For household contacts: 39% reduced progression to disease (Bhargava et al., 2023) |
| Which compartments are affected? | For the reduced risk of on-treatment mortality: DS and DR treatment for symptomatic TB (*Dc*).<br>For the reduced progression to disease: Infected (*I*) compartments. |
| Which transitions are affected? | Reduced risk of treatment mortality: on-treatment mortality parameters (*imort_stDc* and *imort_rtDc*)<br>Reduced progression to disease: progression from infected (*I*) to asymptomatic TB (*Ds*) (*nonsub*) |
| Which parameters are changed? From what value to what value? | Intervention-specific parameter T_mort = 0.67 represents the reduced risk of on-treatment mortality<br>The reduced risk among household contacts is implemented using the following intervention-specific parameters (McQuaid et al., 2025):<br>- Number of HHC = 3.4 (household size in IND - index case)<br>- Proportion of HHC who would develop TB = 3419/100,000<br>- Relative risk of progression = 0.39 |

| | | | | |
|---|---|---|---|---|
| Any further details of changes | The reduction in on-treatment mortality is implemented as a weighted average of the BAU and intervention risks (weighted by the time-varying coverage of the intervention)<br><br>The reduction in progression is implemented using the “trigger” functionality. This uses the number of individuals starting treatment to reduce the progression rate for a proportion of the population (based on the parameters described above). | | | |
| **Costs** | | | | |
| What is being costed, and where are the costs applied? | **Activity** | **Resource use** | **Population** | **Model output** |
| | Standard TB assessment | As BAU (India):<br>**1x** appointment at a clinic<br>**10%** of patients x CXR<br>**21%** of patients x Xpert Ultra | Those entering the diagnostic pathway | n_assess_y[n] |
| | Standard DS-TB treatment | As BAU (India): **7x** monthly appointments at a clinic<br>**6 months** of H+R+Z+E | Those entering treatment for DS-TB | n_treat_DS_y[n] |
| | Standard DR-TB treatment | As BAU (India): **10x** monthly appointments at a clinic<br>**9 months** of Lfx+Cfz+Z+E+Hh+Eto+Bdq | Those entering treatment for DR-TB | n_treat_DR_y[n] |
| | Nutritional supplementation for TB patients | **6x** monthly food baskets, including delivery<br>**0x** additional appointments | People diagnosed with TB receiving nutritional support | n_nutrition_index_y[n] |
| | Nutritional supplementation for household contacts | **6x** monthly food baskets, including delivery [N.B. different from baskets for TB patients]<br>**0x** appointments | Household contacts of people diagnosed with TB receiving nutritional support | n_nutrition_HHC_y[n] |

| | |
|---|---|
| Do costs vary by age? | No |
| **Implementation / Scale-up** | |
| Duration of scale-up | 5 years |
| What happens after implementation? | Continues at 80% for the remaining years |
| How is this implemented in the model? | Straight line continuous increase from 0% at the start of 2025 to 80% at the end of 2029, then maintained at 80% (for both people with TB and their household contacts) |
| How is this implemented in the costings? | Model outputs (n_nutrition_index_y and n_nutrition_HHC_y) count the actual number of people receiving the intervention in the model and so incorporate the scale-up, so the unit costs are applied to the number of people in each of the two categories. |
| **Assumptions** | |
| Any other assumptions to note? | Cost of delivery includes transportation, and also salaries for community health workers who check that the nutrition has reached the intended recipients. |

### 2.5 COMMUNITY-WIDE SCREENING:

The following table describes the specific changes and costs for community-wide screening across all three settings.

| Aspect | Details |
|---|---|
| **Intervention summary** | |
| Intervention number, abbreviation, name | 5. SCR - Community screening |
| Summary description | Annually for 5 years, screen the adult population (≥15 years old) for TB using mobile CXR for all and a confirmatory Xpert MTB/RIF Ultra for those with abnormal findings on the CXR, then start TB treatment on all those diagnosed. |
| Population affected | All, including incarcerated individuals in Brazil. |
| **Changes to the model** | |
| What is the effectiveness of the intervention? | It is expected to mirror the impact observed in the ACT3 trial (Marks et al., 2019) |
| Which compartments are affected? | Adults in all states except those on treatment. |
| Which transitions are affected? | The diagnostic pathway was added as a separate screening process from the usual one. |
| Which parameters are changed? From what value to what value? | Access to community screening was defined as *iaccess*, and scaled up to 80% of the adult population being screened.<br>Test positivity (*itestpos*) was modified to represent CXR screening with confirmatory Xpert Ultra (Schwalb et al., 2025)**:** |

<table>
<tr><td></td><td colspan="4">- S, C, and I = 0.0005 (CXR = 0.085, Xpert Ultra = 0.006)<br>- R and Tc = 0.0201 (CXR = 0.503, Xpert Ultra = 0.040)<br>- Dn = 0.0298 (CXR = 0.677, Xpert Ultra = 0.044)<br>- Ds = 0.5247 (CXR = 0.677, Xpert Ultra = 0.775)<br>- Dc = 0.7725 (CXR = 0.850, Xpert Ultra = 0.909)<br><br>N.B.: This test positivity is assumed to be the same between PLHIV and HIV negative individuals.<br>- Xpert sensitivity and specificity showed confidence intervals that overlapped (Zifodya et al., 2021)<br>- CXR does not seem to differ by HIV status (Fehr et al., 2021)<br><br>Given that Xpert can also be used to test for drug-susceptibility and everybody that tests positive for CXR is tested with Xpert, the probability of receiving a DST (idst_n and idst_p) was set as 1.<br><br>Mortality on treatment (imort_st and imort_rt) was null to represent the improvement in outcome due to early diagnosis, slightly increasing as a result of the success and failure.<br><br>We assumed that individuals would be more likely to accept and receive treatment (iaccept) after being diagnosed with TB with both CXR and Xpert, thus the value was set to 1.</td></tr>
<tr><td>Any further details of changes</td><td colspan="4">None</td></tr>
<tr><td colspan="5">Costs</td></tr>
<tr><td rowspan="3">What is being costed, and where are the costs applied?</td><td>Activity</td><td>Resource use</td><td>Population</td><td>Model output</td></tr>
<tr><td>Standard TB assessment</td><td>As BAU (varies by country)</td><td>Those entering the diagnostic pathway</td><td>n_assess_y[n]</td></tr>
<tr><td>Standard DS-TB treatment</td><td>As BAU (6 months of H+R+Z+E)</td><td>Those entering treatment for DS-TB</td><td>n_treat_DS_y[n]</td></tr>
</table>

| | | | | |
|---|---|---|---|---|
| | Standard DR-TB treatment | As BAU (varies by country) | Those entering treatment for DR-TB | n_treat_DR_y[n] |
| | Community screening - CXR | **1x** CXR in a mobile clinic | People receiving screening | n_screen_y[n] |
| | Community screening - Xpert | **1x** GeneXpert MTB/RIF Ultra | Those with abnormal findings on CXR | n_screen_CXRpositive_y[n] |
| Do costs vary by age? | No | | | |
| **Implementation / Scale-up** | | | | |
| Duration of scale-up | 5 years | | | |
| What happens after implementation? | Remains at 80% for 5 years, then 0% for the remaining 15 years | | | |
| How is this implemented in the model? | Straight line continuous increase from 0% at the start of 2025 to 80% at the end of 2029, then maintained at 80% until the end of 2034 | | | |
| How is this implemented in the costings? | Model output (n_Test_count_y) counts the exact number of people screened each year in the model, and n_screen_CXRpositive_y counts the exact number of people getting follow-up Xpert, so unit costs are just applied directly to these counts. | | | |
| **Assumptions** | | | | |
| Any other assumptions to note? | The cost analysis for this intervention is being run twice for two separate scenarios:<br>‘high-cost community screening’: based on costs consistent with data from ZAF<br>‘low-cost community screening: based on costs consistent with data from BRA/IND | | | |

### 2.6 IMPROVED DIAGNOSIS:

The following table describes the specific changes and costs for improved diagnosis across all three settings.

| Aspect | Details |
|---|---|
| **Intervention summary** | |
| Intervention number, abbreviation, name | 6. DGN - Improved diagnosis |
| Summary description | Everyone assessed for TB will receive GeneXpert MTB/RIF Ultra instead of the previous standard care |
| Population affected | People entering the diagnostic pathway |
| **Changes to the model** | |
| What is the effectiveness of the intervention? | Increased sensitivity for bacteriologically confirmed TB, increased specificity for non-disease states, and increased DST coverage (Zifodya et al., 2021) |
| Which compartments are affected? | All individuals in states not on treatment |
| Which transitions are affected? | Diagnostic pathway in the clinic (in line with the usual care pathway) |
| Which parameters are changed? From what value to what value? | Test positivity (*testpos*) is changed to reflect 80% receiving Xpert Ultra and 20% utilising the previous diagnostic standard practice. New *testpos* values varied by TB state:<br>- *S*, *C*, and *I* = 0.0140<br>- *R* = 0.047<br>- *Tc* = 0.1345<br>- *Dn* = 0.08<br>- *Ds* = 0.525<br>- *Dc* = 0.8189 |

| | | | | |
|---|---|---|---|---|
| | Previous values of *testpos* varied by country and TB state.<br>- *S*, *C* and *I* (BRA // IND // ZAF) = 0.0208 // 0.0294 // 0.0251<br>- *R* (BRA // IND // ZAF) = 0.0635 // 0.0821 // 0.067<br>- *Tc* (BRA // IND // ZAF) = 0.1983 // 0.2778 // 0.2366<br>- *Dn* (BRA // IND // ZAF) = 0.0909 // 0.1307 // 0.0882<br>- *Ds* (BRA // IND // ZAF) = 0.4420 // 0.3549 // 0.4481<br>- *Dc* (BRA // IND // ZAF) = 0.7901 // 7599 // 0.7922<br><br>The probability of getting a DST (*dst_n* and *dst_p*) changed to 0.8, as the proportion receiving Xpert Ultra also increased. The previous value of *dst_n* and *dst_p* varied by country and treatment history:<br>- *dst_n* (BRA // IND // ZAF) = 0.318 // 0.06 // 0.12<br>- *dst_p* (BRA // IND // ZAF) = 0.341 // 0.26 // 0.28 | | | |
| Any further details of changes | Test positivity values for *Dn* were changed to 0.08 for all countries to ensure prevalence did not increase in ZAF due to their large proportion of individuals in *Dn* (the previously calculated value was 0.053). | | | |
| **Costs** | | | | |
| What is being costed, and where are the costs applied? | **Activity** | **Resource use** | **Population** | **Model output** |
| | Improved TB assessment | **1x** appointment at a clinic<br>**1x** GeneXpert MTB/RIF Ultra<br>**0x** CXR | Those entering the diagnostic pathway | n_assess_y[n]<br>[subject to scale-up] |
| | Standard TB assessment | As BAU (see tables 2.1.1-2.1.3):<br>**1x** appointment at a clinic<br>**%** CXR - varies by country<br>**%** Xpert Ultra - varies by country | Those entering the diagnostic pathway | n_assess_y[n]<br>[subject to scale-down] |
| | Standard DS-TB treatment | As BAU (6 months of H+R+Z+E) | Those entering treatment for DS- | n_treat_DS_y[n] |

| | | | | |
|---|---|---|---|---|
| | | | TB | |
| | Standard DR-TB treatment | As BAU (varies by country) | Those entering treatment for DR-TB | n_treat_DR_y[n] |
| Do costs vary by age? | No | | | |
| **Implementation / Scale-up** | | | | |
| Duration of scale-up | 5 years | | | |
| What happens after implementation? | Remains at 80% for the remaining 20 years | | | |
| How is this implemented in the model? | Straight line continuous increase from 0% at the start of 2025 to 80% at the end of 2029, then maintained at 80% for the remaining 20 years. Conversely, those not receiving the improved intervention continue to receive the current standard assessment (100% at start, declining to 20% by 5 years) | | | |
| How is this implemented in the costings? | Cost of improved assessment applied to 16% of total eligible population at the end of Y1, 32% at the end of Y2, 48% at the end of Y3, 64% at the end of Y4, and 80% at the end of Y5 onwards, with the cost of the current standard assessment applied to the rest of the eligible population in each year. | | | |
| **Assumptions** | | | | |
| Any other assumptions to note? | None. | | | |

### 2.7 DRUG SUSCEPTIBILITY TESTING:

The following table describes the specific changes and costs for drug susceptibility testing across all three settings.

| **Aspect** | **Details** |
|---|---|
| **Intervention summary** | |
| Intervention number, abbreviation, name | 7. DST - Drug susceptibility testing |
| Summary description | Everyone diagnosed with TB will be given a GeneXpert MTB/RIF Ultra test to test for rifampicin-resistance, except those who have already had an Xpert test through BAU diagnosis |
| Population affected | Those newly diagnosed with TB |
| **Changes to the model** | |
| What is the effectiveness of the intervention? | Increased initiation of drug-resistant treatment (Zifodya et al., 2021) |
| Which compartments are affected? | All individuals in states not on treatment |
| Which transitions are affected? | Usual diagnostic pathway |
| Which parameters are changed? From what value to what value? | The probability of getting a DST (*dst_n* and *dst_p*) was increased to 0.8 from 2025 (with a five-year scale-up) to 2030. The previous values of *dst_n* and *dst_p* varied by country and treatment history:<br>- *dst_n* (BRA // IND // ZAF) = 0.318 // 0.06 // 0.12<br>- *dst_p* (BRA // IND // ZAF) = 0.341 // 0.26 // 0.28 |
| Any further details of changes | Mortality on DR-TB treatment decreased over time (2025 - 2030) to ensure that a higher number of people accessing DR treatment does not lead to an increased number of deaths. The new values varied by country and year: |

| | | | | |
|---|---|---|---|---|
| | - BRA: *mort_rt* (2026, 2027, 2028, 2029, 2030) = 0.192, 0.186, 0.181, 0.178, 0.175<br>- IND: *mort_rt* (2026, 2027, 2028, 2029, 2030) = 0.230, 0.200, 0.185, 0.170, 0.160<br>- ZAF: *mort_rt* (2026, 2027, 2028, 2029, 2030) = 0.253, 0.234, 0.220, 0.210, 0.200 | | | |
| **Costs** | | | | |
| What is being costed, and where are the costs applied? | **Activity** | **Resource use** | **Population** | **Model output** |
| | Standard TB assessment | As BAU:<br>**1x** appointment at a clinic<br>**%** CXR - varies by country<br>**%** Xpert Ultra -varies by country | Those entering the diagnostic pathway | n_assess_y[n] |
| | Drug susceptibility testing | **1x** Xpert MTB/RIF Ultra | Those diagnosed with TB and not already tested using Xpert | n_diag_noXpert_y[n] [subject to scale-up] |
| | Standard DS-TB treatment | As BAU (6 months of H+R+Z+E) | Those entering treatment for DS-TB | n_treat_DS_y[n] |
| | Standard DR-TB treatment | As BAU (varies by country) | Those entering treatment for DR-TB | n_treat_DR_y[n] |
| Do costs vary by age? | No | | | |
| **Implementation / Scale-up** | | | | |
| Duration of scale-up | 5 years | | | |
| What happens after | Remains at 80% for the remaining 20 years | | | |

| | |
|---|---|
| implementation? | |
| How is this implemented in the model? | Straight line continuous increase from 0% at the start of 2025 to 80% at the end of 2029, then maintained at 80% for the remaining 20 years. The number diagnosed with TB and assumed not to have already been tested with Xpert is calculated outside the model. |
| How is this implemented in the costings? | Cost of additional DST applied to 16% of those diagnosed with TB and estimated not to have had an Xpert during BAU diagnostic process at the end of Y1, 32% at the end of Y2, 48% at the end of Y3, 64% at the end of Y4, and 80% at the end of Y5 onwards. |
| **Assumptions** | |
| Any other assumptions to note? | Note that *n_diag_noXpert_y* is estimated through a simplistic approach, using the proportion of patients evaluated with GeneXpert under the standard TB assessment (BRA = 50%, IND = 21%, ZAF = 61%). |

### 2.8 SCREENING IN PRISONS:

The following table describes the specific changes and costs for screening in prisons in Brazil.

| Aspect | Details |
|---|---|
| **Intervention summary** | |
| Intervention number, abbreviation, name | 8. PRI - Screening in prisons |
| Summary description | Annually screen all adults in prison for TB using GeneXpert MTB/RIF Ultra, then start TB treatment for all those diagnosed. |
| Population affected | Incarcerated individuals in Brazil. |
| **Changes to the model** | |
| What is the effectiveness of the intervention? | Early diagnosis of bacteriologically confirmed TB, increased DST coverage (Pivetta de Araujo et al., 2024). |
| Which compartments are affected? | Adults in all states, except those on treatment, on the prison risk dimension. |
| Which transitions are affected? | The diagnostic pathway, which was added as a separate screening process from the usual diagnostic pathway |
| Which parameters are changed? From what value to what value? | Access to screening was defined as *iaccess*, and scaled up to 80% of the prison population.<br><br>Test positivity (*itestpos*) values were modified to show the probability of testing positive with GeneXpert Ultra. These values varied by TB state:<br>- *S*, *C,* and *I* = 0.006 (Kendall et al., 2021)<br>- *Dn* = 0.044 (Zifodya et al., 2021)<br>- *R and Tc* = 0.040 (World Health Organization, 2023) |

| | | | | |
|---|---|---|---|---|
| | - *Ds* = 0.775 (Zifodya et al., 2021)<br>- *Dc* = 0.909 (Zifodya et al., 2021)<br><br>Given that Xpert can also be used to test for drug-susceptibility, the probability of receiving a DST (*idst_n* and *idst_p*) was set as 1.<br><br>Mortality on treatment (*imort_st* and *imort_rt*) was null to represent the improvement in outcome due to early diagnosis, slightly increasing as a result of the success and failure. | | | |
| Any further details of changes | None | | | |
| **Costs** | | | | |
| What is being costed, and where are the costs applied? | **Activity** | **Resource use** | **Population** | **Model output** |
| | Standard TB assessment | As BAU (Brazil):<br>**1x** appointment at a clinic<br>**35%** of patients x CXR<br>**50%** of patients x GeneXpert Ultra | Those entering the diagnostic pathway | n_assess_y[n] |
| | Standard DS-TB treatment | As BAU (Brazil):<br>**7x** monthly appointments at a clinic<br>**6 months** of H+R+Z+E | Those entering treatment for DS-TB | n_treat_DS_y[n] |
| | Standard DR-TB treatment | As BAU (Brazil):<br>**19x** monthly appointments at a clinic<br>**18 months** of R+Lfx+Z+E | Those entering treatment for DR-TB | n_treat_DR_y[n] |
| | Prison screening | **1x** appointment in prison<br>**1x** GeneXpert MTB/RIF Ultra | People screened in prison | n_screen_prison_y[n] |
| Do costs vary by age? | No. This intervention only applies to adults | | | |

| **Implementation / Scale-up** | |
|---|---|
| Duration of scale-up | 5 years |
| What happens after implementation? | Remains at 80% for the remaining 20 years |
| How is this implemented in the model? | Straight line continuous increase from 0% at the start of 2025 to 80% at the end of 2029, and then maintained for 20 years. |
| How is this implemented in the costings? | The model output n_screen_prison_y[n] counts the exact number of people screened in prison each year, and incorporates scale-up, so the cost of screening is just multiplied by the number screened. |
| **Assumptions** | |
| Any other assumptions to note? | Note that ‘n_prison_y’ counts the number of people in prison at any one point - i.e. this intervention is equivalent to having a screening campaign on a particular day once a year and screening (80% of) all the people in prison on that day. It does not take into account the constant entry and exit of individuals from prison throughout the year, and so it is not the case that (80% of) all people who go into prison will be screened, as some who are in prison for less than a year may miss screening. |

### 2.9 SHORTER DS-TB TREATMENT:

The following table describes the specific changes and costs for shorter DS-TB treatment across all three settings.

| Aspect | Details |
|---|---|
| **Intervention summary** | |
| Intervention number, abbreviation, name | 9. SDS - Shorter DS-TB treatment |
| Summary description | The current standard DS-TB treatment (6 months of H+R+Z+E) replaced with 4 months of H+P+Z+M. |
| Population affected | All individuals receiving treatment for DS-TB. |
| **Changes to the model** | |
| What is the effectiveness of the intervention? | We assumed equal effectiveness of the current DS-TB treatment, given observed non-inferiority in a treatment-shortening trial (Dorman et al., 2021) |
| Which compartments are affected? | None |
| Which transitions are affected? | None |
| Which parameters are changed? From what value to what value? | None |
| Any further details of changes | None |
| **Costs** | |

<table>
<tr><td rowspan="5">What is being costed, and where are the costs applied?</td><td>Activity</td><td>Resource use</td><td>Population</td><td>Model output</td></tr>
<tr><td>Standard TB assessment</td><td>As BAU (varies by country)</td><td>Those entering the diagnostic pathway</td><td>n_assess_y[n]</td></tr>
<tr><td>Short DS-TB treatment</td><td>5x monthly appointments at a clinic<br>4 months of H+P+Z+M</td><td>Those entering treatment for DS-TB</td><td>n_treat_DS_y[n]<br>[subject to scale-up]</td></tr>
<tr><td>Standard DS-TB treatment</td><td>As BAU (same in all countries):<br>7x monthly appointments at a clinic<br>6 months of H+R+Z+E</td><td>Those entering treatment for DS-TB</td><td>n_treat_DS_y[n]<br>[subject to scale-down]</td></tr>
<tr><td>Standard DR-TB treatment</td><td>As BAU (varies by country)</td><td>Those entering treatment for DR-TB</td><td>n_treat_DR_y[n]</td></tr>
<tr><td>Do costs vary by age?</td><td colspan="4">No</td></tr>
<tr><td colspan="5">Implementation / Scale-up</td></tr>
<tr><td>Duration of scale-up</td><td colspan="4">5 years</td></tr>
<tr><td>What happens after implementation?</td><td colspan="4">Remains at 80% for the remaining 20 years</td></tr>
<tr><td>How is this implemented in the model?</td><td colspan="4">Not directly applied in the model as treatment effectiveness is assumed to be the same for both options, and treatment time is not varied for the purpose of the model to avoid paradoxical increase in reinfection, so no distinction within the model between those treated with standard treatment or short treatment.</td></tr>
<tr><td>How is this implemented in the costings?</td><td colspan="4">Cost of short DS-TB treatment applied to 16% of those treated for DS-TB at the end of Y1, 32% at the end of Y2, 48% at the end of Y3, 64% at the end of Y4, and 80% at the end of Y5 onwards, with the cost of current DS-TB treatment applied to the rest of the eligible population in each year</td></tr>
</table>

| Assumptions | |
|---|---|
| Any other assumptions to note? | None. |

### 2.10 SHORTER DR-TB TREATMENT:

The following table describes the specific changes and costs for shorter DR-TB treatment across all three settings.

| Aspect | Details |
|---|---|
| **Intervention summary** | |
| Intervention number, abbreviation, name | 10. SDR - Shorter DR-TB treatment |
| Summary description | The current standard DR-TB treatment (9 months of Lfx+Lzd+Cfz+Z+E+Hh+Bdq in ZAF; 9 months of Lfx+Cfz+Z+E+Hh+Eto+Bdq in IND; 18 months of R+Lfx+Z+E in BRA) replaced with 6 months of BPaLM in all countries. |
| Population affected | All individuals receiving treatment for DR-TB. |
| **Changes to the model** | |
| What is the effectiveness of the intervention? | DR-TB treatment success of 89% (Nyang'wa et al., 2022) |
| Which compartments are affected? | Individuals starting DR-TB treatment |
| Which transitions are affected? | DR-TB treatment outcomes |
| Which parameters are changed? From what value to what value? | Reducing the duration of DR treatment: Increase the value of DR-TB treatment completion rate to reflect the shorter duration of the regimen from 9-18 months to 6 months.<br><br>Increase success in DR-TB (*isucc_rt*) treatment to 0.89 when previous success ranged between 0.52 and 0.55. The difference in increase is assumed to come half from the proportion of mortality on treatment and half from the proportion with unsuccessful treatment. Therefore, mortality on DR-TB (*mort_rt*) treatment also changes. To calculate new *mort_rt* values, we used the following equations: |

| | | | | |
|---|---|---|---|---|
| | - Difference between new and BAU success in DR-TB treatment:<br>new_succ_rt - BAU_succ_rt = difference_succ_rt<br><br>- Calculate half of the difference:<br>difference_succ_rt/2 = half_difference<br><br>- Half of the difference comes from mortality, so the mortality decrease are calculated:<br>BAU_mort_rt - half_difference = new_mort_rt<br><br>The change in *mort_rt* differs by country:<br>- BRA new *mort_rt* = 0.03<br>- BAU_succ_rt = 0.55<br>- BAU_mort_rt = 0.17<br>- IND new *mort_rt* = 0.075<br>- BAU_succ_rt = 0.52<br>- BAU_mort_rt = 0.26<br>- ZAF new *mort_rt* = 0.095<br>- BAU_succ_rt = 0.52<br>- BAU_mort_rt = 0.28 | | | |
| Any further details of changes | None | | | |
| **Costs** | | | | |
| What is being costed, and where are the costs applied? | **Activity** | **Resource use** | **Population** | **Model output** |
| | Standard TB assessment | As BAU (varies by country) | Those entering the diagnostic pathway | n_assess_y[n] |
| | Standard DS-TB treatment | As BAU (6 months of H+R+Z+E) | Those entering treatment for DS-TB | n_treat_DS_y[n] |

| | | | | |
|---|---|---|---|---|
| | Short DR-TB treatment | **7x** monthly appointments at a clinic<br>**6 months** of B+Pa+L+M | Those entering treatment for DR-TB receiving short regimen | n_treat_DR_short_y[n] |
| | Standard DR-TB treatment | As BAU (varies by country) | Those entering treatment for DR-TB receiving BAU regimen | n_treat_DR_y[n] - n_treat_DR_short_y[n] |
| Do costs vary by age? | No | | | |
| **Implementation / Scale-up** | | | | |
| Duration of scale-up from 0% to 80% | 5 years | | | |
| What happens after implementation? | Remains at 80% for the remaining 20 years | | | |
| How is this implemented in the model? | Straight line continuous increase from 0% at the start of 2025 to 80% at the end of 2029, and then maintained for 20 years. Conversely, those not receiving the shortened treatment continue to receive the current DR-TB treatment (100% at start, declining to 20% by 5 years). | | | |
| How is this implemented in the costings? | Model outputs (number receiving any DR-TB treatment and number receiving shortened DR-TB treatment) count the exact number treated in the model each year, so unit costs are just applied directly per person to these outputs. | | | |
| **Assumptions** | | | | |
| Any other assumptions to note? | Adverse events caused by standard or shortened DR-TB treatment are not included in the results, either in terms of loss of utility/increased disability or in terms of the cost of additional medical care. | | | |

### 3. SCALE-UP

The figure shows the shape of coverage for each intervention. Notably, for TB vaccines, we implemented the consensus schedule (World Health Organization, 2022), with 2 mass-campaigns for all adults and continued vaccination of 15-19 year olds. Mass community screening would not be maintained for 25 years, as historical and contemporary data suggest the impact will reduce over time (Esmail et al., 2025; Marks et al., 2019). As a mid-point, we assumed a year year scale up to 80%, followed by five annual campaigns, after which screening would stop.

**Figure S1. Coverage scale-up for interventions.**

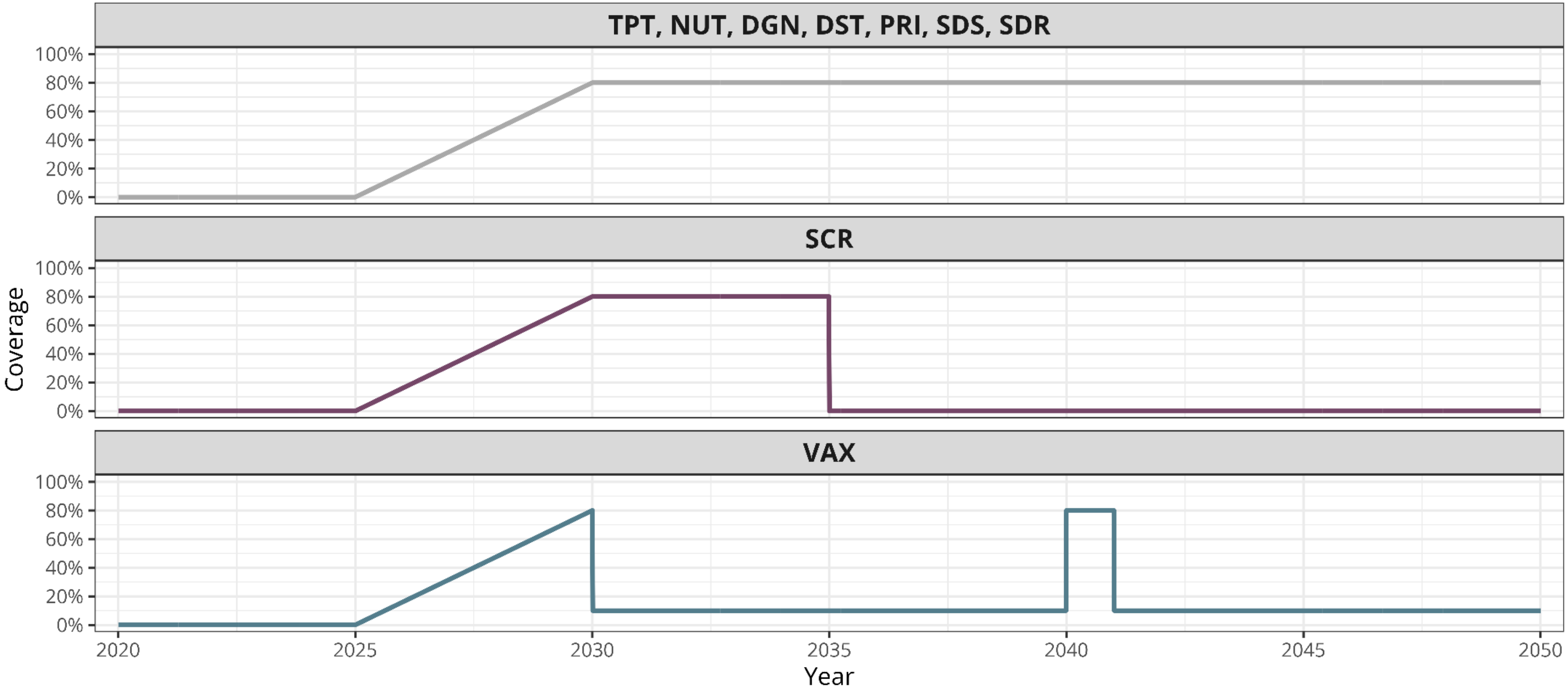
