## Supplementary materials 2 (S2) for "The potential impact, cost and cost-effectiveness of tuberculosis interventions - a modelling exercise"

### SUPPLEMENTARY MATERIAL - S3 - HEALTH ECONOMIC MODEL:
### The potential impact, cost and cost-effectiveness of tuberculosis interventions - a modelling exercise

Katherine C. Horton [1 2], Alvaro Schwalb [1 2 3], Martin J. Harker [1 2 4], Lara Goscé [1 2], Elena Venero-Garcia [1 2], Lily O'Brien [1 2], Arkaprabha Gun [1 2 5], Tom Sumner [1 2], C. Finn McQuaid [1 2], Rebecca A. Clark [1 2], Tomos O. Prys-Jones [1 2], Roel Bakker [1 2 6], Yiran E Liu [7 8], Mmamapudi Kubjane [9], Christian Lienhardt [10 11], Richard G. White [1 2], Rein M.G.J. Houben [1 2]

1 - TB Modelling Group, TB Centre, LSHTM, London, UK; 2 - Department of Infectious Disease Epidemiology, LSHTM, London, UK; 3 - Instituto de Medicina Tropical Alexander von Humboldt, Universidad Peruana Cayetano Heredia, Lima, Peru; 4 - Global Health Economics Centre, LSHTM, London, UK; 5 - Department of Epidemiology, Biostatistics, and Occupational Health, School of Population and Global Health, McGill University, Montreal, QC, Canada; 6 – KNCV Tuberculosis Foundation, The Hague, Netherlands; 7 - SEICHE Center for Health and Justice, Yale University School of Medicine, New Haven, CT, USA; 8 - Justice Collaboratory, Yale Law School, New Haven, CT, USA; 9 - Health Economics and Epidemiology Research Office, Wits Health Consortium, Johannesburg, South Africa; 10 - French Institute for Research in Sustainable Development (IRD), Montpellier, France; 11 - CRDF Global, Arlington, VA, USA

**Correspondence:** Rein M.G.J Houben, Department of Infectious Disease Epidemiology, London School of Hygiene and Tropical Medicine, Keppel St, London, WC1E 7HT United Kingdom

**GitHub:** https://github.com/lshtm-tbmg/PACE-TB

### HEALTH ECONOMIC MODEL

This document describes the health economic model used, first describing the analyses and key principles, resource use and cost (including data sources, followed by the calculation of key health effects.

#### 1. ECONOMIC ANALYSES

A cost–utility analysis was conducted for each of the three countries to compare nationwide implementation of each of the modelled interventions with business as usual (BAU) in that country. The epidemiological outputs generated (listed in detail in *Supplementary Material S2*) were combined with the costs associated with each person assessed for TB, treated for TB or subject to the intervention to generate a total cost of implementing each intervention. Disability-adjusted life years (DALYs) were calculated for each intervention based on the number of years of life lost (YLL) and the number of years lived with disability (YLD) due to TB under each intervention. These were combined to calculate the incremental cost-effectiveness ratio (ICER) for each intervention compared to BAU and to display these results on a cost-effectiveness plane, compared with a proposed cost-effectiveness threshold range for each country.

Each intervention was compared to BAU, rather than an incremental analysis comparing each intervention in turn to the next most effective intervention, because these interventions are not exclusive alternatives, and we are not interested only in the single intervention which would be cost-effective compared to all other interventions. National policy-makers may consider implementing more than one of the interventions concurrently, and will be taking into account other considerations such as total budget impact and local feasibility and acceptability in addition to the cost–utility analysis results. Therefore it would not be helpful in this context to exclude an intervention solely due to being dominated by another intervention (that is, being more costly and less effective), but instead we have supplied full costs and DALY results for all interventions, which can be used to calculate incremental analysis comparing any set of interventions chosen as most relevant in a particular context. Full results can be found in Section 3 of *Supplementary Material S4*.

#### 2. ECONOMIC PRINCIPLES

##### 2.1 PERSPECTIVE

The cost–utility analyses were conducted from the perspective of the health system in each country. This enabled ICERs to be calculated with reference to a cost-effectiveness threshold range in each country, thus allowing the cost-effectiveness of each of the TB control measures investigated in this study to be judged in comparison with other areas of healthcare in each country. This is therefore the most useful perspective for health decision-makers and health funders in each country.

#### 2.2 TIME HORIZON

The analyses consider the effects of the TB interventions being implemented for 25 years from the start of 2025 to the end of 2049 in line with the time period being modelled. We assume a five-year implementation period (2025 - 2029) during which the interventions are steadily rolled out across each country, scaling up to reach an implementation rate of 80% by 2030, which is then maintained for the remaining 20 years. Some interventions, such as community-wide screening and vaccination, have different intervention trajectories post-scale-up.

The expected future health effects of TB episodes and deaths due to TB which occur during the study period are included in the analyses, even if part of these effects would fall after 2049 – for example, if an individual dies from TB in 2045, then the years of life lost included in the calculations of DALYs are based on their age at death and their standard life expectancy had they not had TB, and are not capped at 5 years. The total budget costs of each intervention include all costs of the intervention, as well as TB diagnosis and TB treatment required over the whole 25-year period. During the implementation phase, these costs reflect the gradual scale-up of the interventions. We note that some interventions may be more challenging than others to roll out nationally to 80% coverage within a 5-year timeframe, and that some interventions (notably vaccination) are not currently ready for large scale implementation, however the aim of this analysis is to compare the potential future effects of all of the interventions studied on an equal basis.

#### 2.3 DISCOUNTING

Costs and health benefits in future years are discounted at 5% annually for each country. In South Africa, a 5% discount rate is specified in the Essential Drugs Programme reference case (National Department of Health, 2023). India and Brazil have no officially recommended discount rates (Sharma et al., 2021), and expert opinion varies on the optimal approach (Severens & Milne, 2004). We note the guidance of Haacker et al. that discount rates for lower-middle-income countries should be at least 5% (Haacker et al., 2020), and have chosen to use the same rate in each of the countries to aid in the comparability of the results. Discounting is calculated using the following formula, where *r* is the discount rate and *n* is the number of years into the future:

$$Present\ value = \frac{1}{(1+r)^n}$$

#### 2.4 CURRENCIES AND COST YEAR

Costs and economic results in the paper are reported in US dollars (US$) for the convenience of comparison between countries and for the international audience. These results are repeated below in both US$ and local currencies for each country. Costs are reported for 2023, as that was the latest year for which cost data were available during the study.

#### 2.5 COMPOSITE MEASURES OF TOTAL HEALTH

To assess the overall impact of health interventions on the total health of a population, measures of health which combine both morbidity and mortality into a single valuation are preferred. The most well-known and widely accepted of these are the disability-adjusted life year (DALY) and the quality-adjusted life year (QALY). In this study, we report outcomes in DALYs, to be consistent with the majority of existing tuberculosis studies in the target countries, to aid in comparison.

#### 2.6 COST-EFFECTIVENESS THRESHOLDS

We have chosen to use cost-effectiveness threshold estimates (US$ per DALYs averted) calculated by Ochalek et al. for the International Decision Support Initiative for all of the countries (Ochalek et al., 2018). We have selected the highest and lowest of the four estimates Ochalek et al. calculated for each country to produce a range of potential values for the threshold, to illustrate the uncertainty in these values, which are not officially endorsed as policy thresholds in these countries. We inflated these from 2018 to 2023 values by applying the percentage of per capita GDP estimated by Ochalek et al. in 2015 to the 2023 GDP of each country (Ochalek et al., 2018; World Bank, 2025a).

**Table E1. Cost-effectiveness threshold ranges for each country**

| Country | High or low threshold | Value (2015 US$)* | % 2015 GDP | Value (2023 US$) |
|---|---|---|---|---|
| Brazil | Bottom of threshold range | 6048.00 | 68% | 6967.55 |
| Brazil | Top of threshold range | 9318.00 | 104% | 10734.72 |
| India | Bottom of threshold range | 264.00 | 17% | 413.47 |
| India | Top of threshold range | 363.00 | 23% | 568.52 |
| South Africa | Bottom of threshold range | 2480.00 | 41% | 2443.59 |

| | | | | |
|---|---|---|---|---|
| South Africa | Top of threshold range | 3334.00 | 55% | 3285.06 |

*Source: (Ochalek et al., 2018)

### 3. RESOURCE USE AND COSTS

We calculated the cost of implementing each intervention across the whole population by assessing the additional resources required for the intervention, as well as the effect of the intervention on the quantity of TB diagnoses and TB treatment provided. We multiply each resource used by the unit cost of that resource in each country and the number of people affected per year to calculate the total cost of each intervention over 25 years.

#### 3.1 RESOURCE USE

The definition of each intervention was described in detail in the Intervention Supplementary Material, and this was translated into a list of resources required per person receiving the intervention. These were principally informed by existing published research studies most closely resembling the interventions being modelled, with details checked by experts and agreed by the study team. We identified 17 separate activities which are required to deliver the 10 interventions, shown in Table E2 below. These activities were further divided into 17 resource use components (see Table E3 below). These components were chosen based on the availability of existing cost data. Each component represents a collection of resources required as part of a TB intervention. For example, 'DS-TB standard treatment drugs' includes the cost of each of the four drugs included in a standard six-month treatment regimen for DS-TB. 'CXR at mobile van' includes all the resources required to run a mobile screening van, including the vehicle, CXR machine and staffing, divided proportionately among the expected number of users. Resource use includes equipment (such as CXR and GeneXpert machines), drugs, consumable supplies, and staffing. Unrelated health costs, that is, any impacts on future demand for healthcare unrelated to TB, are not included in this analysis, nor are potential impacts on other areas of public spending, such as education or welfare.

**Table E2. Activities making up each intervention.**

| Code | Intervention | **Ac tiv** | Va cci | H H | TP T | T P | N utr | N utr | Sc re | Sc re | Cli ni | Cli ni | Dr ug | D S- | D S- | D R- | D R- | Pri so |
|---|---|---|---|---|---|---|---|---|---|---|---|---|---|---|---|---|---|---|
| BAU | Business as usual | | | | | | | | | | ■ | | | ■ | | ■ | | |
| VAX | Vaccination | | ■ | | | | | | | | ■ | | | ■ | | ■ | | |
| TPT | TB preventive treatment | | | ■ | ■ | ■ | | | | | ■ | | | ■ | | ■ | | |
| NTN | Nutrition | | | | | | ■ | ■ | | | ■ | | | ■ | | ■ | | |
| SCR | Community screening | | | | | | | | ■ | ■ | ■ | | | ■ | | ■ | | |
| DGN | Improved diagnosis | | | | | | | | | | ■ | ■ | | ■ | | ■ | | |
| DST | Drug susceptibility testing | | | | | | | | | | ■ | | ■ | ■ | | ■ | | |
| PRI | Screening in prisons | | | | | | | | | | ■ | | | ■ | | ■ | | ■ |
| SDS | Shorter DS-TB treatment | | | | | | | | | | ■ | | | ■ | ■ | ■ | | |

| | |
|---|---|
| SDR | Shorter DR-TB treatment |

**Table E3. Resource use components required, per person, for each activity.**

| Component | Vaccination | HHCM assessment | TPT for DS-TB | TPT for DR-TB | Nutrition for TB patients | Nutrition for household contacts | Screening CXR (high estimate) | Screening CXR (low estimate) | Screening Xpert | Clinic diagnosis – standard [BRA] | Clinic diagnosis – standard [IND] | Clinic diagnosis – standard [ZAF] | Clinic diagnosis - improved | DST | DS-TB treatment - standard | DS-TB treatment - shortened | DR-TB treatment – standard [BRA] | DR-TB treatment – standard [IND] | DR-TB treatment – standard [ZAF] | DR-TB treatment - shortened | Prison screening |
|---|---|---|---|---|---|---|---|---|---|---|---|---|---|---|---|---|---|---|---|---|---|
| Vaccine | 2 | 0 | 0 | 0 | 0 | 0 | 0 | 0 | 0 | 0 | 0 | 0 | 0 | 0 | 0 | 0 | 0 | 0 | 0 | 0 | 0 |
| Carry out vaccination | 2 | 0 | 0 | 0 | 0 | 0 | 0 | 0 | 0 | 0 | 0 | 0 | 0 | 0 | 0 | 0 | 0 | 0 | 0 | 0 | 0 |
| Appointment at clinic | 0 | 1 | 4 | 7 | 0 | 0 | 0 | 0 | 0 | 1 | 1 | 1 | 1 | 0 | 7 | 5 | 19 | 10 | 10 | 7 | 0 |
| Appointment in prison | 0 | 0 | 0 | 0 | 0 | 0 | 0 | 0 | 0 | 0 | 0 | 0 | 0 | 0 | 0 | 0 | 0 | 0 | 0 | 0 | 1 |
| CXR at clinic/hospital | 0 | 0 | 0 | 0 | 0 | 0 | 0 | 0 | 0 | 0.35 | 0.10 | 0.03 | 0 | 0 | 0 | 0 | 0 | 0 | 0 | 0 | 0 |
| CXR at mobile van - high estimate | 0 | 0 | 0 | 0 | 0 | 0 | 1 | 0 | 0 | 0 | 0 | 0 | 0 | 0 | 0 | 0 | 0 | 0 | 0 | 0 | 0 |
| CXR at mobile van - low estimate | 0 | 0 | 0 | 0 | 0 | 0 | 0 | 1 | 0 | 0 | 0 | 0 | 0 | 0 | 0 | 0 | 0 | 0 | 0 | 0 | 0 |
| Xpert Ultra sample collection/processing in clinic | 0 | 1 | 0 | 0 | 0 | 0 | 0 | 0 | 1 | 0.50 | 0.21 | 0.61 | 1 | 1 | 0 | 0 | 0 | 0 | 0 | 0 | 0 |
| Xpert Ultra sample collection/processing in prison | 0 | 0 | 0 | 0 | 0 | 0 | 0 | 0 | 0 | 0 | 0 | 0 | 0 | 0 | 0 | 0 | 0 | 0 | 0 | 0 | 1 |
| DS-TB standard treatment drugs | 0 | 0 | 0 | 0 | 0 | 0 | 0 | 0 | 0 | 0 | 0 | 0 | 0 | 0 | 1 | 0 | 0 | 0 | 0 | 0 | 0 |
| DS-TB shortened treatment drugs | 0 | 0 | 0 | 0 | 0 | 0 | 0 | 0 | 0 | 0 | 0 | 0 | 0 | 0 | 0 | 1 | 0 | 0 | 0 | 0 | 0 |
| DR-TB standard treatment drugs | 0 | 0 | 0 | 0 | 0 | 0 | 0 | 0 | 0 | 0 | 0 | 0 | 0 | 0 | 0 | 0 | 1 | 1 | 1 | 0 | 0 |
| DR-TB shortened treatment drugs | 0 | 0 | 0 | 0 | 0 | 0 | 0 | 0 | 0 | 0 | 0 | 0 | 0 | 0 | 0 | 0 | 0 | 0 | 0 | 1 | 0 |
| TPT for DS-TB drugs | 0 | 0 | 1 | 0 | 0 | 0 | 0 | 0 | 0 | 0 | 0 | 0 | 0 | 0 | 0 | 0 | 0 | 0 | 0 | 0 | 0 |
| TPT for DR-TB drugs | 0 | 0 | 0 | 1 | 0 | 0 | 0 | 0 | 0 | 0 | 0 | 0 | 0 | 0 | 0 | 0 | 0 | 0 | 0 | 0 | 0 |
| Nutritional support for TB patients | 0 | 0 | 0 | 0 | 1 | 0 | 0 | 0 | 0 | 0 | 0 | 0 | 0 | 0 | 0 | 0 | 0 | 0 | 0 | 0 | 0 |
| Nutritional support for household contacts | 0 | 0 | 0 | 0 | 0 | 1 | 0 | 0 | 0 | 0 | 0 | 0 | 0 | 0 | 0 | 0 | 0 | 0 | 0 | 0 | 0 |

#### 3.2 COSTS

##### 3.2.1 COSTS PER COMPONENT

Costs were obtained for each of the resource use components for each country. As for resource use, where possible, costs were taken from the existing published research studies on which each of the interventions was based, where these were applicable to one or more of the target countries. Additional data were taken from other publicly available sources, preferring official national sources where available. Drug prices were taken from the Global Drug Facility international TB drugs price list (Global Drug Facility, 2025). World Health Organisation country-specific costs for inpatient stays and outpatient consultations were used. Costs quoted in previous years or in different currencies were inflated to the common cost year and converted to US dollars using World Bank exchange rates (World Bank, 2025b). Vaccination costs were sourced from costs for the roll-out of the human papillomavirus vaccine among adolescents and expert opinion (Clark 2023).

For the cost of community screening, using chest X-ray in a mobile clinic, the costs obtained varied notably between countries, being considerably higher in South Africa than in Brazil or India. As these interventions are provided to very large populations this introduces large uncertainty into the true potential cost of implementing this intervention at a national scale. We therefore decided to model two alternative scenarios as separate interventions: 'high-cost community screening' using the cost of screening from South Africa in all countries; and 'low-cost community screening' using the Brazilian cost of screening in Brazil and South Africa, and the Indian cost in India. Results for both these alternatives are presented alongside results for BAU and the eight other interventions.

**Table E4. Costs by resource use component in each country (2023 US$)**

| Component | Brazil | | India | | South Africa | |
|---|---|---|---|---|---|---|
| | Cost | Source | Cost | Source | Cost | Source |
| Vaccine | 2.625 | (Clark et al., 2023) | 2.625 | (Clark et al., 2023) | 2.625 | (Clark et al., 2023) |
| Carry out vaccination | 2.610 | (Clark et al., 2023) | 2.610 | (Clark et al., 2023) | 2.610 | (Clark et al., 2023) |

| | | | | | | |
|---|---|---|---|---|---|---|
| Appointment at clinic | 4.498 | (WHO: Economic Evaluation and Analysis, 2021) | 7.795 | (WHO: Economic Evaluation and Analysis, 2021) | 19.949 | (WHO: Economic Evaluation and Analysis, 2021) |
| Appointment in prison | 2.257 | (da Silva Santos et al., 2021) | N/A | | N/A | |
| CXR at clinic or hospital | 6.744 | (da Silva Santos et al., 2021) | 4.002 | (Chatterjee et al., 2021) | 18.292 | (Kubjane et al., 2024) |
| CXR at mobile van - high estimate | 41.396 | Using South Africa cost | 41.396 | Using South Africa cost | 41.396 | (Kubjane et al., 2024) |
| CXR at mobile van - low estimate | 5.360 | (da Silva Santos et al., 2021) | 2.425 | (Datta et al., 2019) | 5.360 | Using Brazil cost |
| GeneXpert Ultra sample collection & processing in clinic | 19.360 | (da Silva Santos et al., 2021; Global Drug Facility, 2025) | 24.365 | (Chatterjee et al., 2021; Global Drug Facility, 2025) | 16.932 | (Kubjane et al., 2024) |
| GeneXpert Ultra sample collection & processing in prison | 19.260 | (da Silva Santos et al., 2021; Global Drug Facility, 2025) | n/a | | n/a | |
| DS-TB standard treatment drugs | 87.831 | (Global Drug Facility, 2025) | 87.831 | (Global Drug Facility, 2025) | 87.831 | (Global Drug Facility, 2025) |
| DS-TB shortened treatment drugs | 176.389 | (Global Drug Facility, 2025) | 176.389 | (Global Drug Facility, 2025) | 176.389 | (Global Drug Facility, 2025) |
| DR-TB standard treatment drugs | 3514.819 | (Global Drug Facility, 2025) | 372.588 | (Global Drug Facility, 2025) | 343.539 | (Global Drug Facility, 2025) |

| | | | | | | |
|---|---|---|---|---|---|---|
| DR-TB shortened treatment drugs | 362.898 | (Global Drug Facility, 2025) | 362.898 | (Global Drug Facility, 2025) | 362.898 | (Global Drug Facility, 2025) |
| TPT for DS-TB drugs | 4.247 | (Global Drug Facility, 2025) | 4.247 | (Global Drug Facility, 2025) | 4.247 | (Global Drug Facility, 2025) |
| TPT for DR-TB drugs | 9.648 | (Global Drug Facility, 2025) | 9.648 | (Global Drug Facility, 2025) | 9.648 | (Global Drug Facility, 2025) |
| Nutritional support for TB patients | n/a | | 16.527 | (Bhargava et al., 2023) | n/a | |
| Nutritional support for household contact | n/a | | 5.968 | (Bhargava et al., 2023) | n/a | |

##### 3.2.2 COSTS PER ACTIVITY

Applying the component costs in Table E4 to the resource use in Table E3 gives the following costs for each activity:

**Table E5. Costs by activity in each country (2023 US$)**

| Activity | Parameter name | Activity cost | | |
|---|---|---|---|---|
| | | **Brazil** | **India** | **South Africa** |
| Vaccination | c_a_vax | 10.470 | 10.470 | 10.470 |
| HHCM assessment | c_a_HHCM | 23.858 | 32.161 | 36.882 |
| TPT for DS-TB | c_a_TPT_DS | 22.241 | 35.428 | 84.044 |
| TPT for DR-TB | c_a_TPT_DR | 41.136 | 64.215 | 149.292 |

| | | | | |
|---|---|---|---|---|
| Nutrition for TB patients | c_a_nutrition_index | n/a | 16.527 | n/a |
| Nutrition for household contacts | c_a_nutrition_contact | n/a | 5.968 | n/a |
| Community screening CXR (high-cost estimate) | c_a_screen_CXR_hi | 41.396 | 41.396 | 41.396 |
| Community screening CXR (low-cost estimate) | c_a_screen_CXR_lo | 5.360 | 2.425 | 5.360 |
| Community screening Xpert Ultra | c_a_screen_Xpert | 19.360 | 24.365 | 16.932 |
| Clinic diagnosis - standard | c_a_diag_standard | 16.539 | 13.312 | 30.849 |
| Clinic diagnosis - improved | c_a_diag_improved | 23.858 | 32.161 | 36.882 |
| Drug sensitivity testing | c_a_DST | 19.360 | 24.365 | 16.932 |
| DS-TB treatment - standard | c_a_treat_DS_standard | 119.320 | 142.398 | 227.476 |
| DS-TB treatment - shortened | c_a_treat_DS_improved | 198.881 | 215.366 | 276.135 |
| DR-TB treatment - standard | c_a_treat_DR_standard | 3600.287 | 450.541 | 543.030 |
| DR-TB treatment - shortened | c_a_treat_DR_improved | 394.386 | 417.465 | 502.542 |
| Prison screening | c_a_prison | 21.517 | n/a | n/a |

To capture uncertainty in costs we generated a gamma distribution around each point estimate in Table E5 using a broad range of +/-50%, which was chosen to be conservative. Using these distributions we randomly sampled 500 sets of costs for each country, without correlating costs within each set. Each cost set was randomly matched to one of the epidemiological model parameter sets to provide cost multipliers for each activity, generating 500 unique runs for each intervention.

##### 3.2.3 COSTS PER INTERVENTION

The cost of each intervention was calculated annually, based on relevant model outputs for each activity (for example, the number of people receiving the intervention, or the number of people receiving treatment for DS-TB), multiplied by the activity costs above, then summed across the 25-year span of the intervention. Calculations were based on the following equations for each intervention, where y[n] represents the year in the model (from y1 to y25). For the improved diagnosis and shorter DS-TB treatment interventions, annual adjustments were made to calibrate the scale-up of the intervention from 0% to 80% of those eligible over the first 5 years, thereafter remaining at 80%. For the other interventions, scale-up of the interventions was embedded in the model outputs, included in these calculations (see *Supplementary Material S2 Interventions* for more information on the model outputs and the methodology used to scale up each of the interventions). Future costs were discounted as explained in section 2.3 above.

**Table E6. Cost equations by intervention**

| Code | Intervention | Cost equation |
|---|---|---|
| BAU | Business as usual | n_assess_y[n] * c_a_diag_standard + n_treat_DS_y[n] * c_a_treat_DS_standard + n_treat_DR_y[n] * c_a_treat_DR_standard |
| VAX | Vaccination | n_assess_y[n] * c_a_diag_standard + n_treat_DS_y[n] * c_a_treat_DS_standard + n_treat_DR_y[n] * c_a_treat_DR_standard + n_vaccinated_y[n] * c_a_vax |
| TPT | TB preventive treatment | n_assess_y[n] * c_a_diag_standard + n_treat_DS_y[n] * c_a_treat_DS_standard + n_treat_DR_y[n] * c_a_treat_DR_standard + (n_HHC_DS_y[n] + n_HHC_DR_y[n]) * c_a_HHCM + n_HHC_DSTPT_y[n] * c_a_TPT_DS + n_HHC_DRTPT_y[n] * c_a_TPT_DR |
| NTN | Nutrition | n_assess_y[n] * c_a_diag_standard + n_treat_DS_y[n] * c_a_treat_DS_standard + n_treat_DR_y[n] * c_a_treat_DR_standard + n_nutrition_index_y[n] * c_a_nutrition_index + n_nutrition_HHC_y[n] * c_a_nutrition_contact |
| SCR | Community screening | n_assess_y[n] * c_a_diag_standard + n_treat_DS_y[n] * c_a_treat_DS_standard + n_treat_DR_y[n] * c_a_treat_DR_standard + n_screen_y[n] * c_a_screen_CXR + n_screen_CXRpositive_y[n] * c_a_screen_Xpert |
| DGN | Improved diagnosis | n_assess_y[n] * ([scale-down %] * c_a_diag_standard + [scale-up %] * c_a_diag_improved) + n_treat_DS_y[n] * c_a_treat_DS_standard + n_treat_DR_y[n] * c_a_treat_DR_standard |
| DST | Drug susceptibility testing | n_assess_y[n] * c_a_diag_standard + n_treat_DS_y[n] * c_a_treat_DS_standard + n_treat_DR_y[n] * c_a_treat_DR_standard + n_diag_noXpert_y[n] * c_a_DST |

| | | |
|---|---|---|
| PRI | Screening in prisons | n_assess_y[n] * c_a_diag_standard + n_treat_DS_y[n] * c_a_treat_DS_standard + n_treat_DR_y[n] * c_a_treat_DR_standard + n_prison_y[n] * c_a_prison |
| SDS | Shorter DS-TB treatment | n_assess_y[n] * c_a_diag_standard + n_treat_DS_y[n] * ([scale-down %] * c_a_treat_DS_standard + [scale-up %] * c_a_treat_DS_improved) + n_treat_DR_y[n] * c_a_treat_DR_standard |
| SDR | Shorter DR-TB treatment | n_assess_y[n] * c_a_diag_standard + n_treat_DS_y[n] * c_a_treat_DS_standard + (n_treat_DR_y[n] - n_treat_DR_short_y[n]) * c_a_treat_DR_standard + n_treat_DR_short_y[n] * c_a_treat_DR_improved |

The total costs of each intervention include both the cost of implementing the particular intervention and the costs of TB diagnosis in clinics and TB treatment. The amount of TB diagnosis or treatment required may be decreased by the intervention (for example, for preventive interventions), or increased (for example, for diagnostic interventions), whether immediately or in subsequent years, and so these changes will also be captured in the total costs.

For community screening, the cost of the screening (c_a_screen_CXR) was varied between c_a_screen_CXR_hi and c_a_screen_CXR_lo in the high-cost or low-cost scenarios

### 4. HEALTH EFFECTS

#### 4.1 DALY CALCULATIONS

The time horizon was set at 2050. DALYs were calculated for each intervention in each country, according to the WHO and Global Burden of Disease definition as the sum of years of life lost (YLL) due to premature mortality in the population and years lived with disability (YLD) for people living with the health condition or its consequences (Institute for Health Metrics and Evaluation, 2024; World Health Organization, n.d.). YLL were further divided into years lost from TB mortality (deaths of people with symptomatic clinical TB disease) and post-TB mortality (the increased death rate expected in people who have previously had TB). YLD were further divided into disability for people with symptomatic TB and disability for people with post-TB conditions. DALYs were calculated by combining epidemiological outputs from the modelling in TBMod (such as the number of TB episodes and the number of deaths from TB), life table data, and disability weights, according to the equations below:

**YLL =** TB death DALYs + Post-TB death DALYs

**YLD =** TB disability DALYs + Post-TB disability DALYs

**TB death DALYs =** number of lives lost from TB × expected life years lost per death (discounted)

**DALY_mort_TB_y[n]_ag[n] =** n_TBdeath_y[n]_ag[n] * LY_healthy_y[n]_ag[n]

**Post-TB death DALYs =** number of people with post-TB × expected life years lost per person from premature deaths (discounted)

**DALY_mort_postTB_y[n]_ag[n] =** n_postTB_y[n]_ag[n] * LY_lost_postTB_y[n]_ag[n]

**TB disability DALYs =** number of people with clinical TB × duration of clinical TB × disability weight for TB × discount factor

**DALY_dis_TB_y[n]_ag[n] =** n_Dc_y[n]_ag[n] * duration_Dc_ag[n] * dw_TB * (1+disc)^-(n-1)

**Post-TB disability DALYs =** number of people with post-TB × duration of post-TB (discounted) × disability weight for post-TB

**DALY_dis_postTB_y[n]_ag[n] =** n_postTB_y[n]_ag[n] * LY_postTB_y[n]_ag[n] * dw_postTB

Where y[n] represents the year in the model (from y1 to y25), and ag[n] represents the age groups in the model (from ag1 [0-4 years] to ag16 [75-99 years]). These were calculated annually over each of the 25 years and for all age groups and were summed to give the total DALYs for each intervention. DALYs averted were calculated by subtracting the total DALYs for a particular intervention from the total DALYs for BAU.

#### 4.2 POST-TB SEQUELAE

A history of previous TB impacts on the future expected health of an individual due to a wide range of 'post-TB' sequelae, contributing a substantial proportion of the DALYs lost due to TB (Menzies et al., 2021; Quaife et al., 2020). This includes both a decrease in the quality of life of the individual (that is, a disability weight should be applied to years of life following TB disease) and an increase in the expected mortality rate above the standard age-based national mortality rate. We adopt the reductions in future morbidity and mortality reported in previous work (Menzies et al., 2021; Quaife et al., 2020).

The effects of post-TB are only applied to people who have experienced symptomatic TB in the model. This is because research on the impacts of post-TB sequelae is based on individuals who have been diagnosed and treated with TB, who will normally have had symptomatic TB. It is possible that there may also be long-term effects of asymptomatic or non-infectious TB, in which case the health burden of post-TB sequelae calculated here may conservatively underestimate the true total burden. The additional mortality impact was not modelled within TBMod, but has been calculated based on an increase in the age-based mortality rate for those individuals who have previously experienced TB.

#### 4.3 DISABILITY WEIGHTS

A disability weight for TB disease (dw_TB) of 0.333 for all countries was sourced from the Global Burden of Disease project (Institute for Health Metrics and Evaluation, 2024). This was applied to each individual in the 'symptomatic TB' state and in the 'TB treatment' states, for as long as they remained in those states. No disability weight was applied to people with asymptomatic TB or non-infectious TB. Total years of life with disability due to TB are therefore calculated from the mean duration of an episode of symptomatic TB in the model (duration_Dc_ag[n]) multiplied by the number of people experiencing symptomatic TB each year (n_Dc_y[n]_ag[n]) multiplied by the disability weight (dw_TB), discounted with respect to the relevant year (equation for 'TB disability DALYs' above). For people who have previously had TB, a disability weight of 0.036 (dw_postTB) was applied for the rest of their lifetimes. This is the median weight found by Menzies et al. in their study of the impact of post-TB sequelae (Menzies et al., 2021).

#### 4.4 LIFE YEARS

Life tables from the UN World Population Prospects were used to calculate for each country the standard (healthy) discounted life expectancy of an individual in each of the 16 age groups used in the modelling in each year from 2025 to 2049 (LY_healthy_y[n]_ag[n]) (United Nations, n.d.). This equates to the years of life lost when an individual in a given age group dies in a given year due to TB (equation 'TB death DALYs' above). A mortality rate modifier of 1.14x was adopted from Menzies et al. (Menzies et al., 2021) to recalculate discounted life expectancy for those who have previously experienced TB (LY_postTB_y[n]ag[n]). This equates to the years lived with disability after an individual in a given age group has symptomatic TB in a given year. The number of people in this group (n_postTB_y[n]_ag[n]) is calculated by subtracting the number of TB deaths from the number of people who experience symptomatic TB for the first time each year (equation 'Post-TB disability DALYs' above). By subtracting post-TB life expectancy from health life expectancy, we calculate the expected years of life lost due to post-TB sequelae (LY_lost_postTB_y[n]_ag[n]). This equates to the years of life lost when an individual in a given age group dies in a given year due to post-TB sequelae (equation 'Post-TB death DALYs' above).

**Table E7. Discounted healthy and post-TB life expectancy by year and age group for each country**

| Year | Age group | Healthy life expectancy | | | Post-TB life expectancy | | | Life years lost from post-TB | | |
|---|---|---|---|---|---|---|---|---|---|---|
| | | BRA | IND | ZAF | BRA | IND | ZAF | BRA | IND | ZAF |
| 2025 (year 1) | 0 - 4 years | 19.657 | 19.451 | 19.061 | 19.585 | 19.363 | 18.936 | 0.072 | 0.087 | 0.125 |
| | 5 - 9 years | 19.464 | 19.288 | 18.811 | 19.378 | 19.194 | 18.671 | 0.086 | 0.094 | 0.140 |
| | 10 - 14 years | 19.202 | 19.013 | 18.418 | 19.095 | 18.901 | 18.249 | 0.107 | 0.112 | 0.169 |
| | 15 - 19 years | 18.900 | 18.666 | 17.924 | 18.771 | 18.532 | 17.718 | 0.129 | 0.135 | 0.205 |
| | 20 - 24 years | 18.578 | 18.257 | 17.349 | 18.430 | 18.098 | 17.104 | 0.149 | 0.159 | 0.245 |
| | 25 - 29 years | 18.199 | 17.764 | 16.707 | 18.029 | 17.577 | 16.422 | 0.170 | 0.187 | 0.285 |
| | 30 - 34 years | 17.717 | 17.155 | 16.008 | 17.520 | 16.935 | 15.687 | 0.197 | 0.220 | 0.322 |
| | 35 - 39 years | 17.128 | 16.421 | 15.265 | 16.899 | 16.164 | 14.912 | 0.229 | 0.257 | 0.353 |
| | 40 - 44 years | 16.417 | 15.546 | 14.471 | 16.152 | 15.248 | 14.093 | 0.265 | 0.298 | 0.378 |
| | 45 - 49 years | 15.575 | 14.511 | 13.566 | 15.271 | 14.170 | 13.167 | 0.304 | 0.341 | 0.399 |
| | 50 - 54 years | 14.602 | 13.332 | 12.530 | 14.259 | 12.949 | 12.113 | 0.343 | 0.383 | 0.417 |
| | 55 - 59 years | 13.486 | 12.033 | 11.335 | 13.105 | 11.615 | 10.904 | 0.381 | 0.418 | 0.430 |
| | 60 - 64 years | 12.231 | 10.615 | 9.970 | 11.817 | 10.171 | 9.531 | 0.414 | 0.444 | 0.439 |
| | 65 - 69 years | 10.871 | 9.110 | 8.480 | 10.433 | 8.653 | 8.044 | 0.438 | 0.457 | 0.436 |
| | 70 - 74 years | 9.417 | 7.606 | 6.925 | 8.969 | 7.153 | 6.507 | 0.448 | 0.454 | 0.417 |
| | 75+ years | 7.284 | 5.573 | 4.708 | 6.846 | 5.153 | 4.343 | 0.438 | 0.420 | 0.365 |
| 2026 (year 2) | 0 - 4 years | 18.730 | 18.535 | 18.170 | 18.662 | 18.453 | 18.052 | 0.068 | 0.082 | 0.117 |
| | 5 - 9 years | 18.548 | 18.380 | 17.934 | 18.466 | 18.291 | 17.802 | 0.081 | 0.089 | 0.132 |
| | 10 - 14 years | 18.300 | 18.119 | 17.563 | 18.200 | 18.013 | 17.404 | 0.101 | 0.106 | 0.159 |
| | 15 - 19 years | 18.014 | 17.790 | 17.096 | 17.893 | 17.663 | 16.903 | 0.121 | 0.127 | 0.193 |
| | 20 - 24 years | 17.710 | 17.402 | 16.553 | 17.569 | 17.252 | 16.322 | 0.140 | 0.150 | 0.231 |
| | 25 - 29 years | 17.350 | 16.934 | 15.945 | 17.189 | 16.757 | 15.677 | 0.160 | 0.177 | 0.268 |
| | 30 - 34 years | 16.893 | 16.356 | 15.281 | 16.707 | 16.148 | 14.977 | 0.186 | 0.208 | 0.303 |
| | 35 - 39 years | 16.335 | 15.659 | 14.571 | 16.118 | 15.415 | 14.238 | 0.216 | 0.243 | 0.334 |
| | 40 - 44 years | 15.661 | 14.828 | 13.811 | 15.411 | 14.545 | 13.454 | 0.250 | 0.282 | 0.357 |
| | 45 - 49 years | 14.862 | 13.844 | 12.946 | 14.575 | 13.520 | 12.568 | 0.287 | 0.324 | 0.378 |
| | 50 - 54 years | 13.938 | 12.722 | 11.955 | 13.613 | 12.358 | 11.560 | 0.325 | 0.364 | 0.395 |
| | 55 - 59 years | 12.877 | 11.485 | 10.814 | 12.517 | 11.088 | 10.404 | 0.361 | 0.397 | 0.409 |
| | 60 - 64 years | 11.684 | 10.135 | 9.511 | 11.291 | 9.713 | 9.094 | 0.393 | 0.422 | 0.418 |
| | 65 - 69 years | 10.387 | 8.700 | 8.090 | 9.972 | 8.265 | 7.675 | 0.415 | 0.435 | 0.415 |

| | | | | | | | | | | |
|---|---|---|---|---|---|---|---|---|---|---|
| | 70 - 74 years | 9.000 | 7.265 | 6.607 | 8.574 | 6.833 | 6.209 | 0.426 | 0.432 | 0.398 |
| | 75+ years | 6.960 | 5.323 | 4.493 | 6.544 | 4.923 | 4.145 | 0.416 | 0.400 | 0.348 |

Healthy and post-TB life expectancy in years 1 and 2 per age group for each country. Values for the complete time horizon (up to 2050) are available in the GitHub repository: https://github.com/lshtm-tbmg/NIH2.
