## Supplementary materials 2 (S2) for "The potential impact, cost and cost-effectiveness of tuberculosis interventions - a modelling exercise"

### SUPPLEMENTARY MATERIAL - S4 - RESULTS:
### The potential impact, cost and cost-effectiveness of tuberculosis interventions - a modelling exercise

Katherine C. Horton [1 2], Alvaro Schwalb [1 2 3], Martin J. Harker [1 2 4], Lara Goscé [1 2], Elena Venero-Garcia [1 2], Lily O'Brien [1 2], Arkaprabha Gun [1 2 5], Tom Sumner [1 2], C. Finn McQuaid [1 2], Rebecca A. Clark [1 2], Tomos O. Prys-Jones [1 2], Roel Bakker [1 2 6], Yiran E Liu [7 8], Mmamapudi Kubjane [9], Christian Lienhardt [10 11], Richard G. White [1 2], Rein M.G.J. Houben [1 2]

1 - TB Modelling Group, TB Centre, LSHTM, London, UK; 2 - Department of Infectious Disease Epidemiology, LSHTM, London, UK; 3 - Instituto de Medicina Tropical Alexander von Humboldt, Universidad Peruana Cayetano Heredia, Lima, Peru; 4 - Global Health Economics Centre, LSHTM, London, UK; 5 - Department of Epidemiology, Biostatistics, and Occupational Health, School of Population and Global Health, McGill University, Montreal, QC, Canada; 6 – KNCV Tuberculosis Foundation, The Hague, Netherlands; 7 - SEICHE Center for Health and Justice, Yale University School of Medicine, New Haven, CT, USA; 8 - Justice Collaboratory, Yale Law School, New Haven, CT, USA; 9 - Health Economics and Epidemiology Research Office, Wits Health Consortium, Johannesburg, South Africa; 10 - French Institute for Research in Sustainable Development (IRD), Montpellier, France; 11 - CRDF Global, Arlington, VA, USA

**Correspondence:** Rein M.G.J Houben, Department of Infectious Disease Epidemiology, London School of Hygiene and Tropical Medicine, Keppel St, London, WC1E 7HT United Kingdom

**GitHub:** https://github.com/lshtm-tbmg/PACE-TB

### OVERVIEW

This document provides additional results to those reported in the main paper. Throughout, we report median and 95% quantilles for the uncertainty intervals, except for the ICERS, where we use mean differences (section 3.3-3.4).

### 1. CALIBRATION PLOTS AND POSTERIORS

See supplementary materials S1, section 7, for calibration targets, data sources and methods.

#### 1.1 Calibration plots

##### 1.1.1 Brazil Calibration results

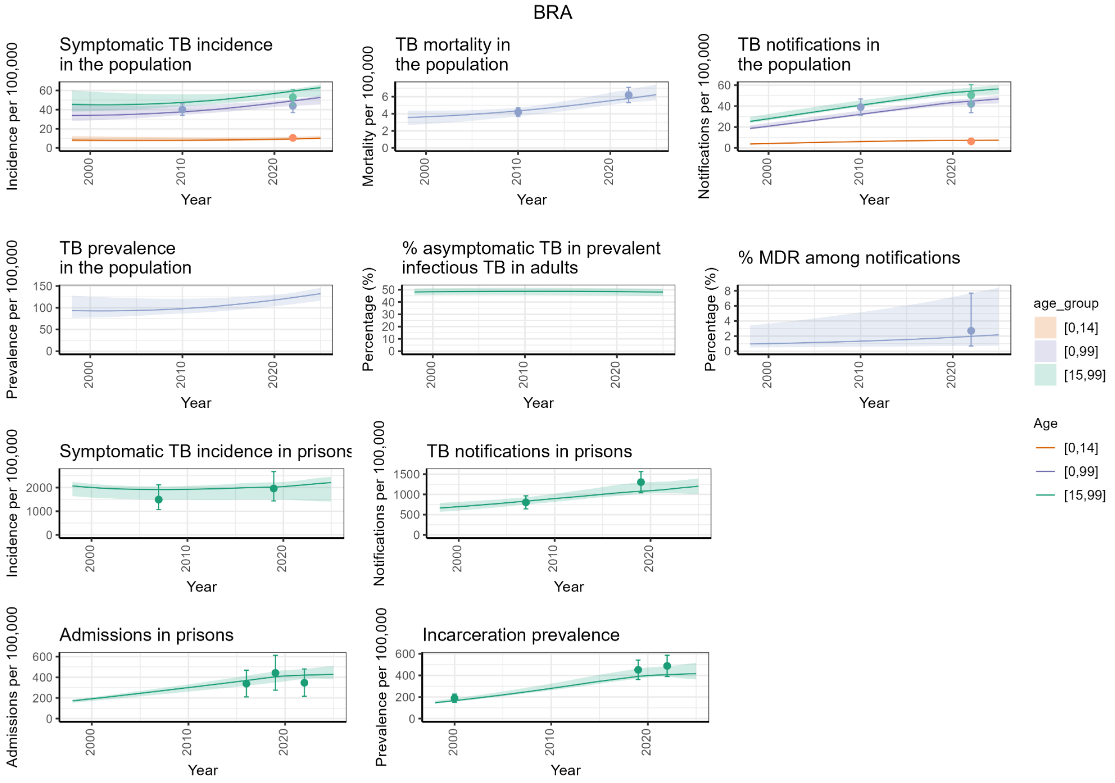

##### 1.1.2 India Calibration results

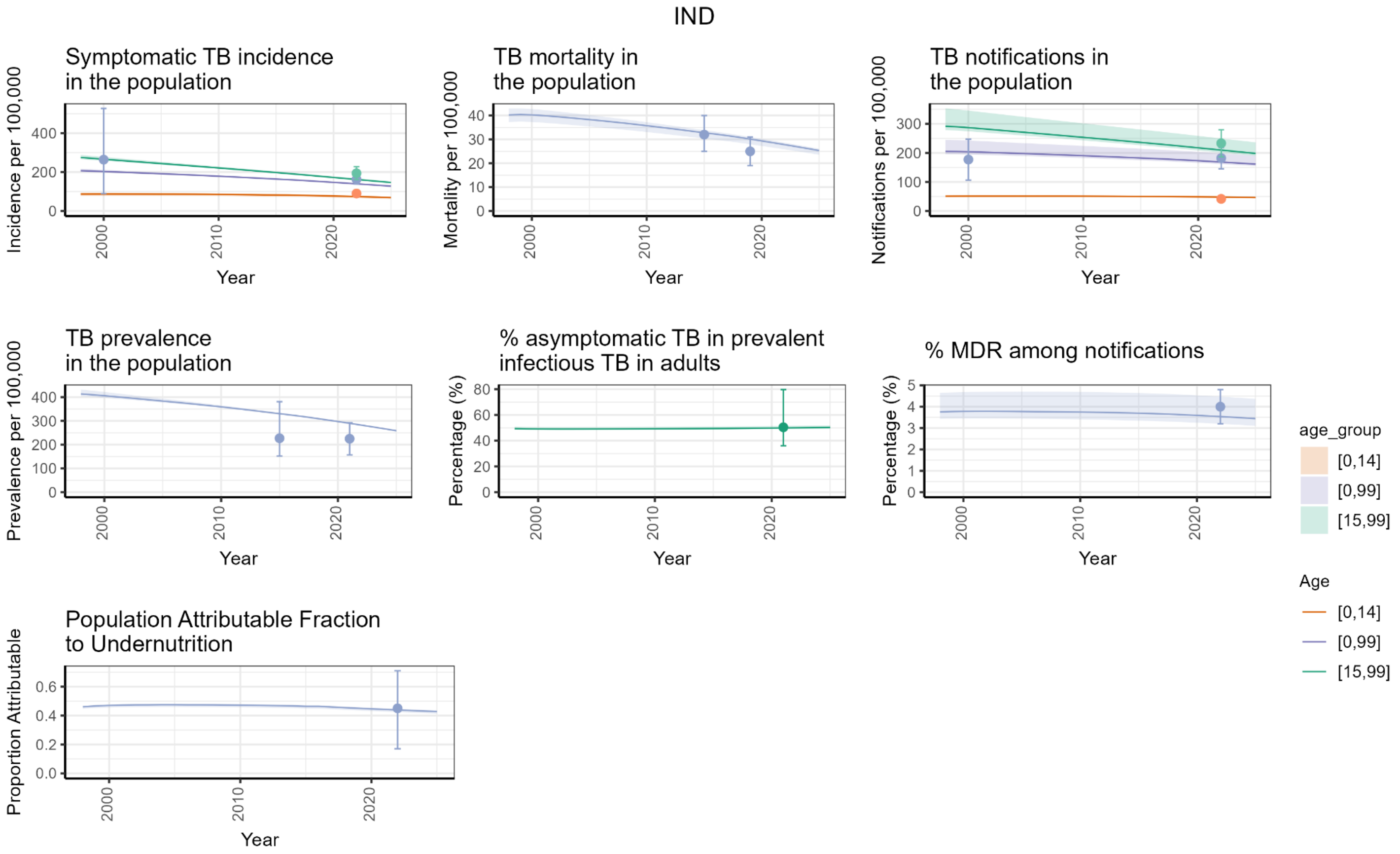

##### 1.1.3 South Africa Calibration results

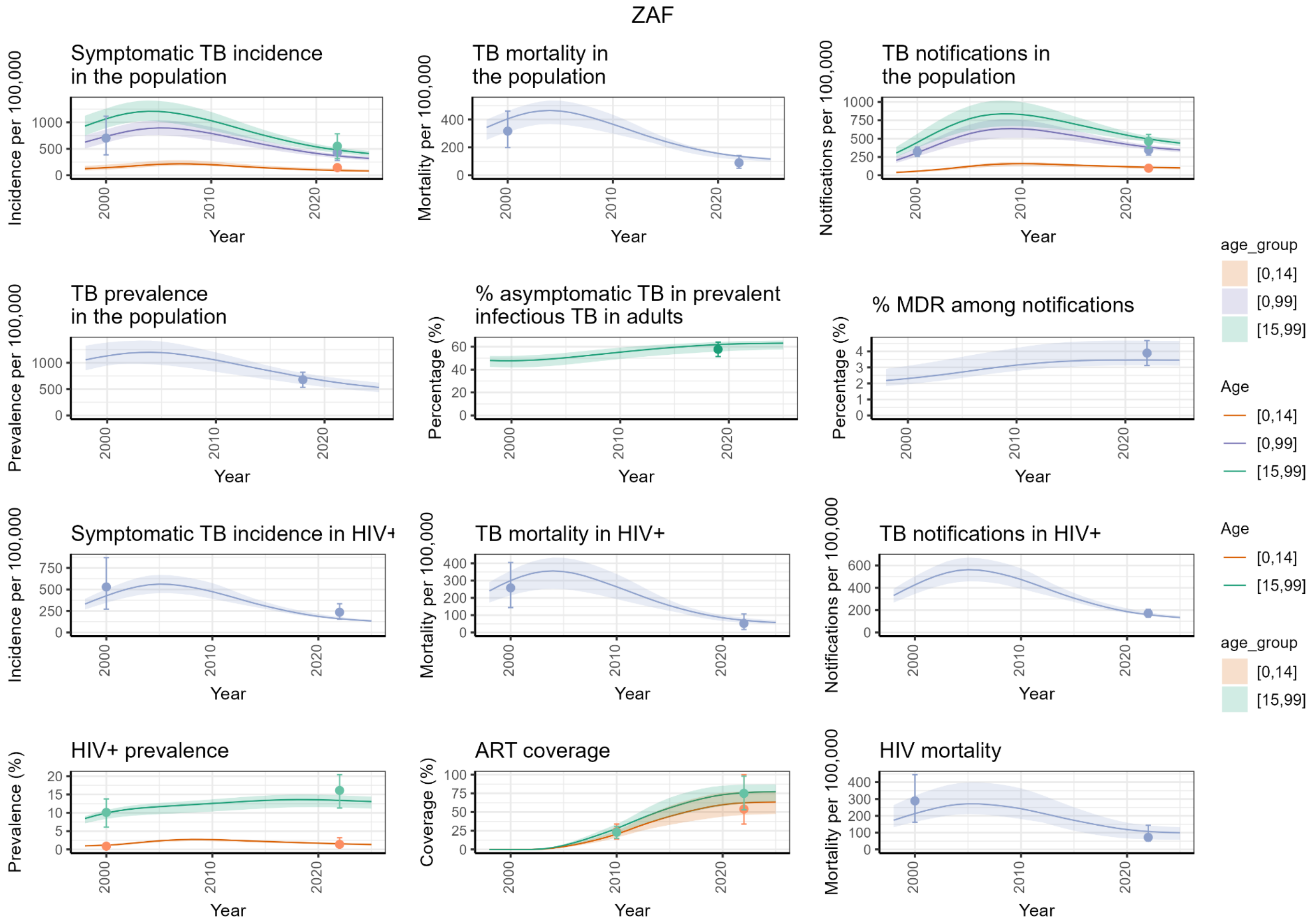

#### 1.2 Calibration posteriors

Figures posterior distributions of parameters for each country (BRA = Brazil, IND = India, ZAF = South Africa). See supplementary materials S1 for parameters and prior ranges

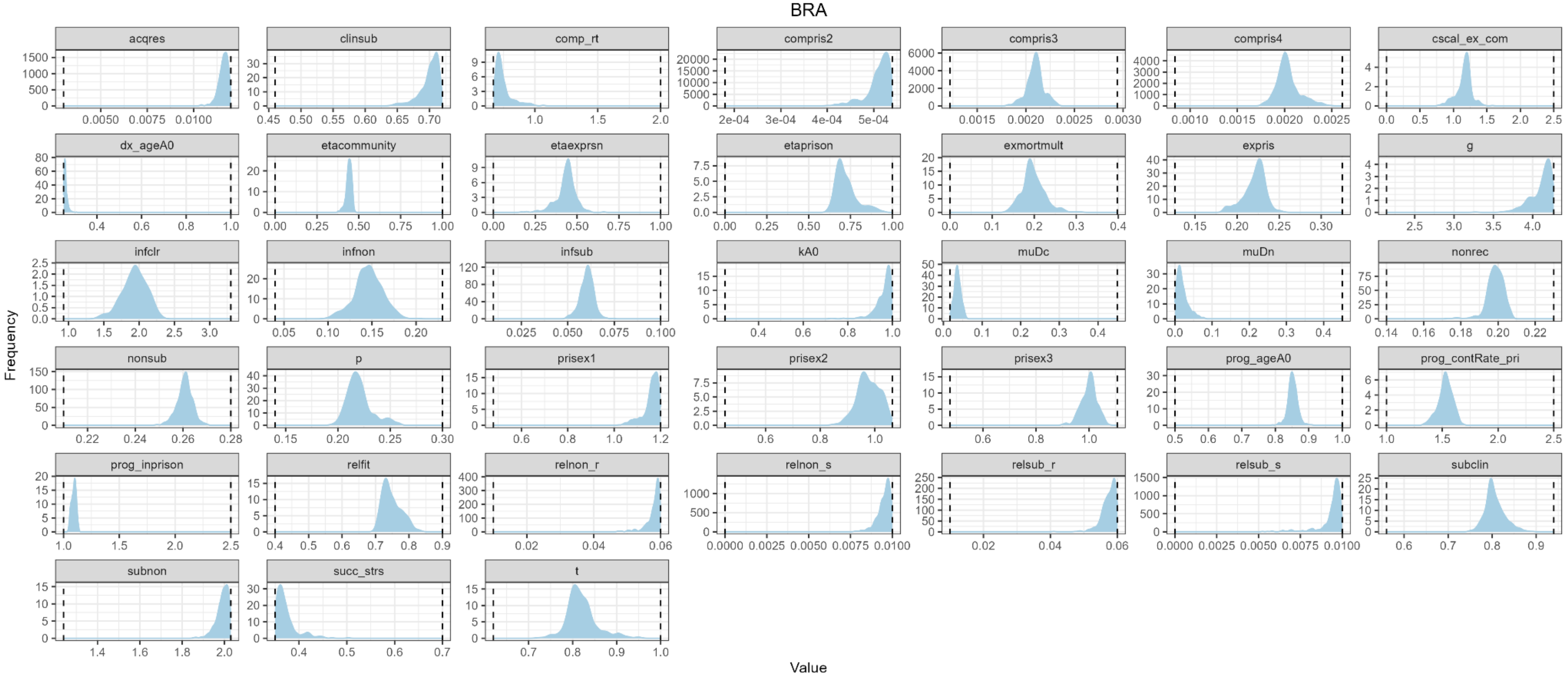

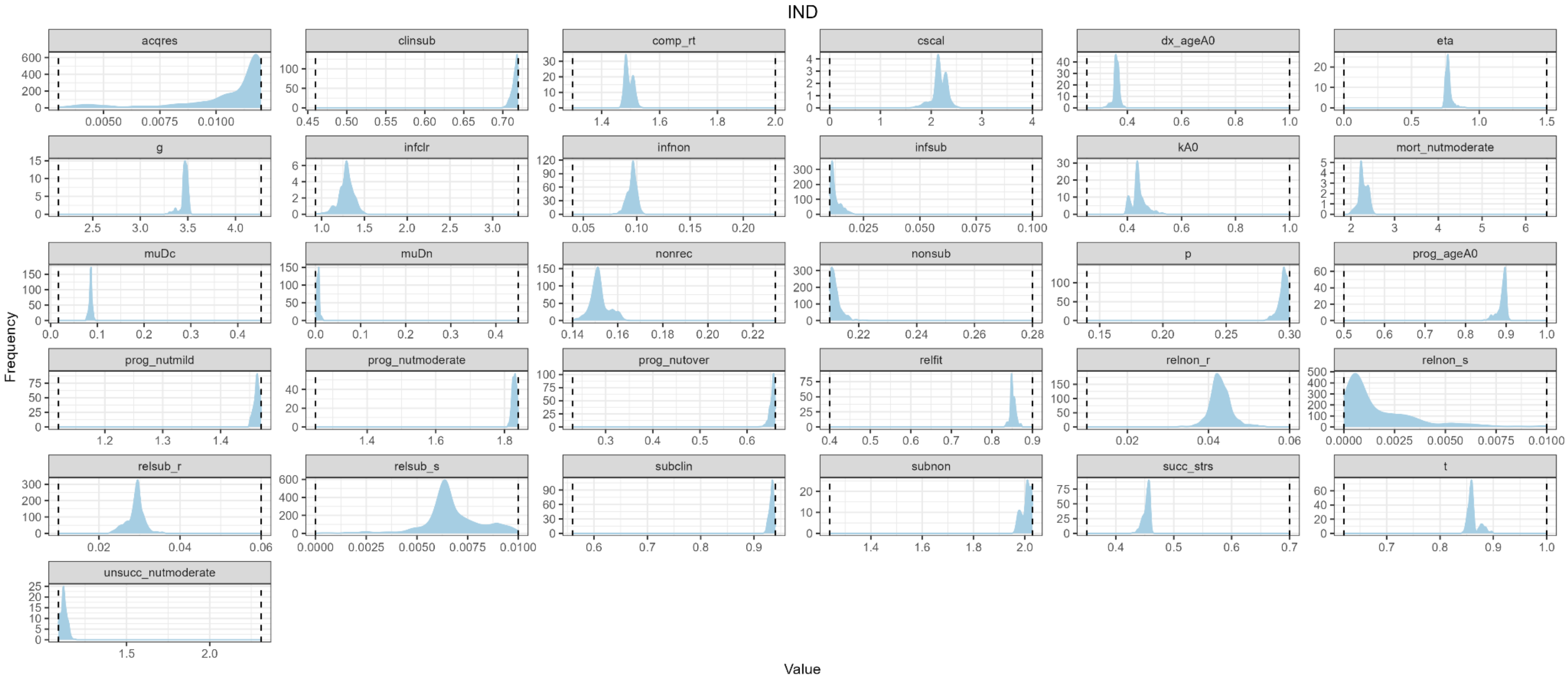
IND
acqres
clinsub
comp_rt
cscal
dx_ageA0
eta
g
infclr
infnon
infsub
kA0
mort_nutmoderate
muDc
muDn
nonrec
nonsub
p
prog_ageA0
prog_nutmild
prog_nutmoderate
prog_nutover
relfit
relnon_r
relnon_s
relsub_r
relsub_s
subclin
subnon
succ_strs
t
unsucc_nutmoderate
Frequency
Value

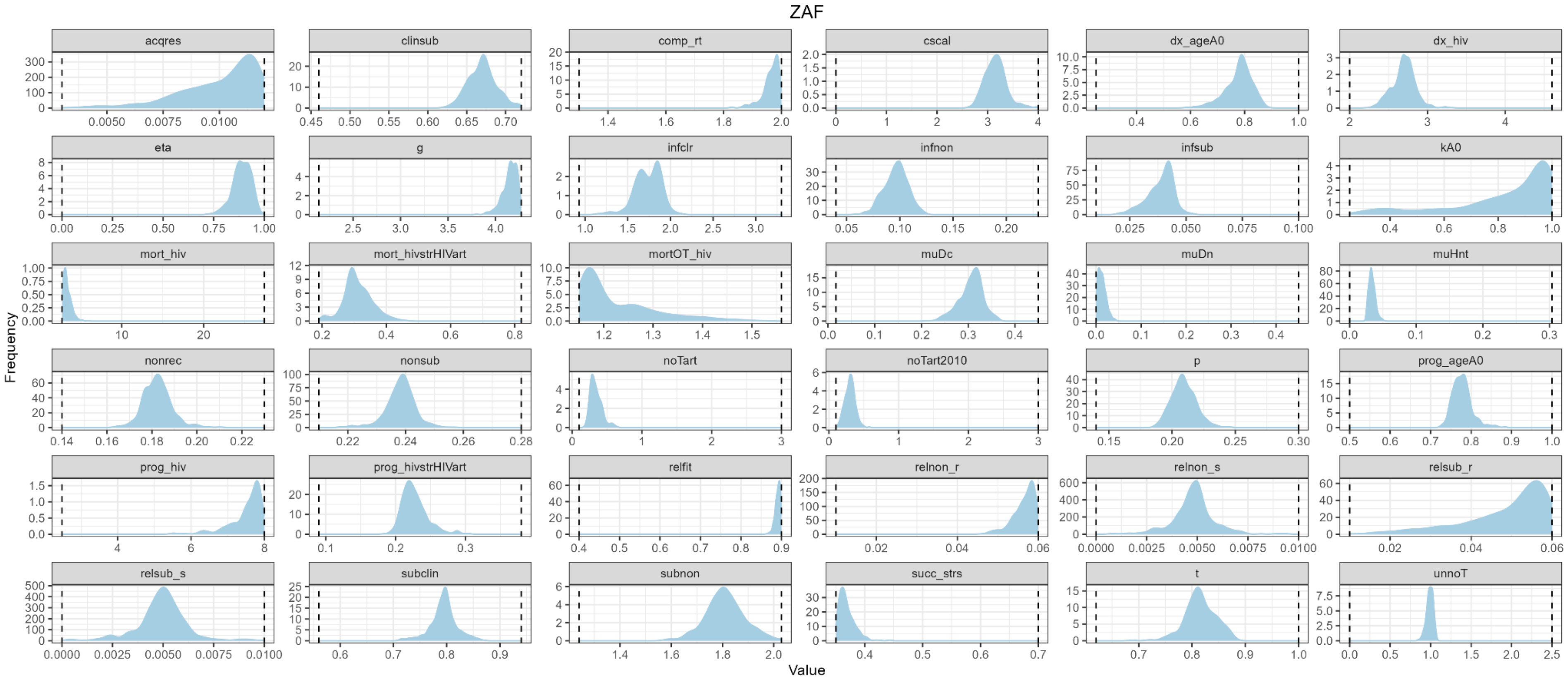
ZAF
acqres
clinsub
comp_rt
cscal
dx_ageA0
dx_hiv
eta
g
infclr
infnon
infsub
kA0
mort_hiv
mort_hivstrHIVart
mortOT_hiv
muDc
muDn
muHnt
nonrec
nonsub
noTart
noTart2010
p
prog_ageA0
prog_hiv
prog_hivstrHIVart
relfit
relnon_r
relnon_s
relsub_r
relsub_s
subclin
subnon
succ_strs
t
unnoT
Frequency
Value

### 2. EPIDEMIOLOGICAL IMPACT

#### 2.1 TB Prevalence trends

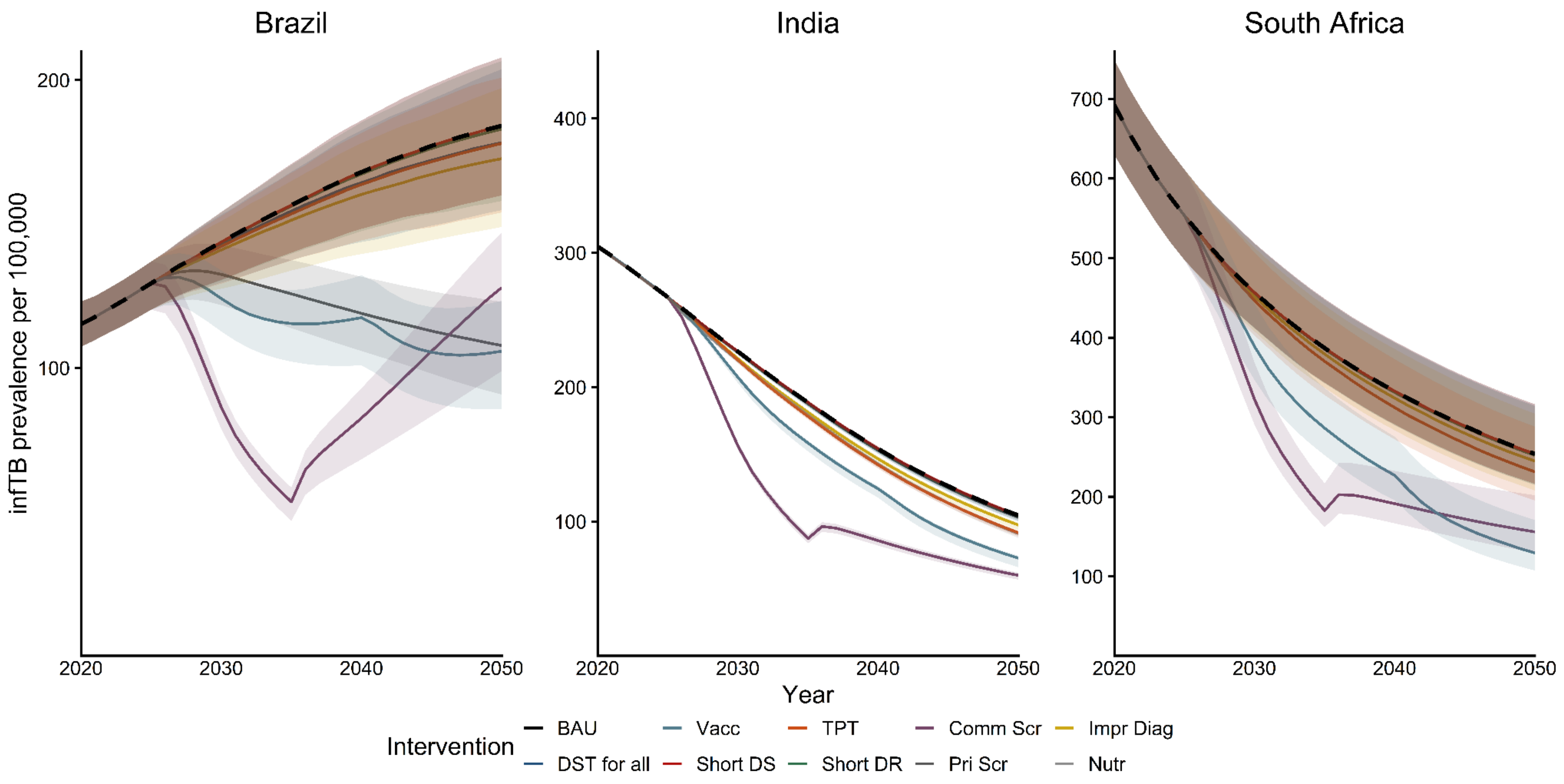

#### 2.2 TB Mortality trends

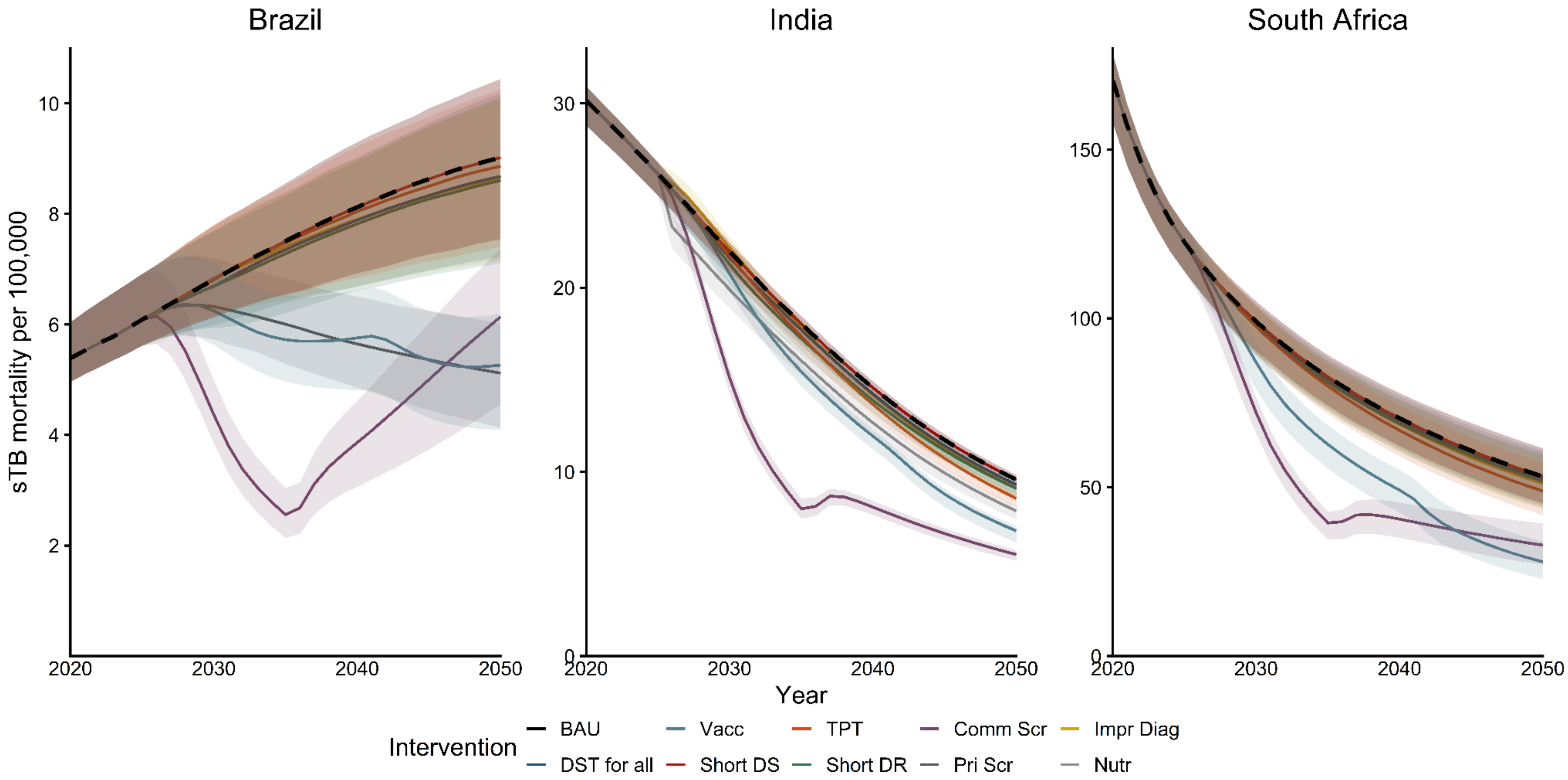

#### 2.3 Incident sTB episodes averted - proportion of total

| Incident symptomatic TB episodes averted by 2050 (percent relative to BAU) | | | |
|---|---|---|---|
| Intervention | Brazil | India | South Africa |
| Vacc | 27.9% (26.6-30.2%) | 14.8% (14.3-17.1%) | 25% (22.6-28%) |
| TPT | 1.6% (1.4-1.8%) | 4.3% (4.3-4.7%) | 3.5% (3-3.9%) |
| Comm Scr | 38% (35.8-42%) | 31.7% (30.5-32.8%) | 31% (29.8-32.4%) |
| Impr Diag | 2.4% (1.8-6%) | 2.2% (2-2.6%) | 1.3% (1.2-1.5%) |
| DST for all | 0.8% (0.2-4%) | 0.4% (0.3-0.5%) | 0.1% (0.1-0.1%) |
| Short DS | 0% (0-0%) | 0% (0-0%) | 0% (0-0%) |
| Short DR | 0.1% (0-0.7%) | 0.3% (0.2-0.3%) | 0.2% (0.1-0.2%) |
| Pri Scr | 23.3% (22-25.2%) | - | - |
| Nutr | - | 5.1% (4.9-5.5%) | - |

2.4 Incident sTB episodes averted - absolute numbers (1000s)

| Incident symptomatic TB episodes averted by 2050 (number relative to BAU, in thousands) | | | |
|---|---|---|---|
| Intervention | Brazil | India | South Africa |
| Vacc | 1,226K (1,086-1,333K) | 6,038K (5,738-6,895K) | 1,112K (963-1,328K) |
| TPT | 71K (60-81K) | 1,759K (1,726-1,904K) | 159K (133-179K) |
| Comm Scr | 1,662K (1,536-1,821K) | 12,860K (12,419-13,182K) | 1,382K (1,250-1,556K) |
| Impr Diag | 103K (74-262K) | 880K (816-1,052K) | 60K (51-71K) |
| DST for all | 34K (7-175K) | 146K (126-205K) | 4K (3-6K) |
| Short DS | 0K (0-0K) | 0K (0-0K) | 0K (0-0K) |
| Short DR | 6K (2-29K) | 105K (92-138K) | 7K (5-11K) |
| Pri Scr | 1,022K (870-1,151K) | - | - |
| Nutr | - | 2,050K (2,006-2,255K) | - |

2.5 Reduction in TB prevalence in 2050 - proportional (graph)

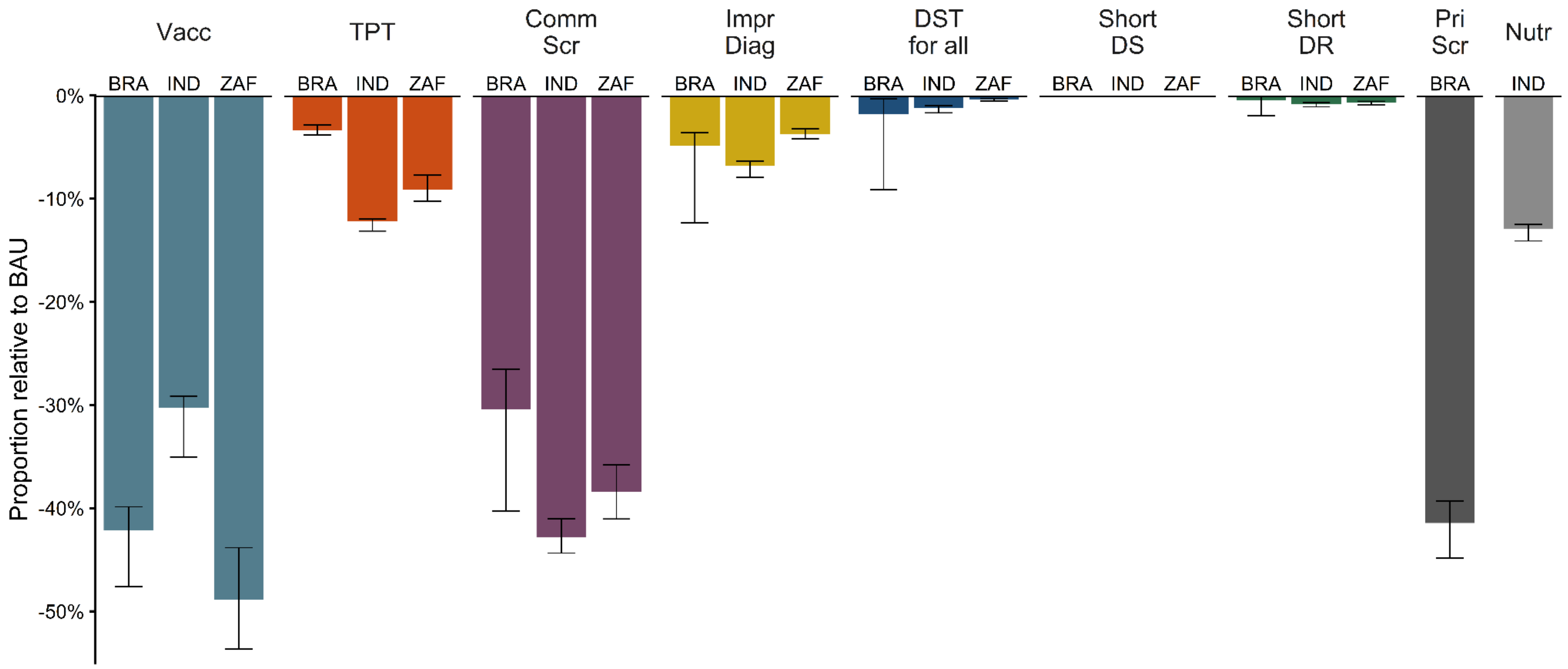

#### 2.6 Reduction in TB prevalence in 2050 - proportional (table)

| Decline in infectious TB prevalence in 2050 (percent relative to BAU) | | | |
|---|---|---|---|
| Intervention | Brazil | India | South Africa |
| Vacc | 42.2% (39.9-47.6%) | 30.3% (29.2-35%) | 48.8% (43.8-53.7%) |
| TPT | 3.4% (2.9-3.8%) | 12.2% (12-13.1%) | 9.1% (7.7-10.2%) |
| Comm Scr | 30.4% (26.5-40.3%) | 42.8% (41-44.4%) | 38.4% (35.8-41.1%) |
| Impr Diag | 4.9% (3.6-12.3%) | 6.8% (6.4-7.9%) | 3.8% (3.2-4.2%) |
| DST for all | 1.8% (0.3-9.1%) | 1.2% (1-1.7%) | 0.4% (0.3-0.5%) |
| Short DS | 0% (0-0%) | 0% (0-0%) | 0% (0-0%) |
| Short DR | 0.4% (0.1-2%) | 0.8% (0.7-1.1%) | 0.7% (0.6-0.9%) |
| Pri Scr | 41.4% (39.3-44.8%) | - | - |
| Nutr | - | 12.9% (12.5-14.1%) | - |

#### 2.7 TB deaths averted - proportion of total

| TB deaths averted by 2050 (percent relative to BAU) | | | |
|---|---|---|---|
| Intervention | Brazil | India | South Africa |
| Vacc | 25.6% (24.2-28%) | 12.8% (12.3-14.9%) | 23.6% (21.1-26.5%) |
| TPT | 0.8% (0.4-1.3%) | 4.1% (3.9-4.4%) | 3.6% (3-4%) |
| Comm Scr | 42.2% (40.3-45.9%) | 35.3% (33.4-36.3%) | 32.9% (31.7-34.4%) |
| Impr Diag | 1.8% (0.8-5.6%) | 1.2% (0.9-1.4%) | 1.8% (1.6-2%) |
| DST for all | 1.9% (0.4-7.2%) | 1.8% (1.5-2.3%) | 0.8% (0.7-1.1%) |
| Short DS | 0% (0-0%) | 0% (0-0%) | 0% (0-0%) |
| Short DR | 2.5% (0.7-8.6%) | 3.5% (3.1-4.6%) | 1.7% (1.5-2.3%) |
| Pri Scr | 25.2% (23.2-27.1%) | - | - |
| Nutr | - | 11.1% (10.7-11.9%) | - |

#### 2.8 TB deaths - absolute numbers (1000s)

| TB deaths averted by 2050 (number relative to BAU, in thousands) | | | |
|---|---|---|---|
| Intervention | Brazil | India | South Africa |
| Vacc | 117K (102-135K) | 871K (802-1,004K) | 324K (279-381K) |
| TPT | 4K (2-6K) | 274K (261-294K) | 50K (40-55K) |
| Comm Scr | 194K (174-217K) | 2,392K (2,238-2,458K) | 456K (410-498K) |
| Impr Diag | 8K (3-26K) | 80K (61-93K) | 26K (22-28K) |
| DST for all | 9K (2-33K) | 120K (104-159K) | 11K (9-15K) |
| Short DS | 0K (0-0K) | 0K (0-0K) | 0K (0-0K) |
| Short DR | 11K (3-39K) | 239K (209-304K) | 24K (20-32K) |
| Pri Scr | 115K (97-135K) | - | - |
| Nutr | - | 749K (727-798K) | - |

2.9 Disability-adjusted life years - total (millions)

| Disability-adjusted life years (number) | | | |
|---|---|---|---|
| Intervention | Brazil | India | South Africa |
| BAU | 10.8M (9.5-12.1M) | 137.7M (132.6-140.7M) | 23M (20.7-25M) |
| Vacc | 7.9M (6.8-8.9M) | 118.2M (112.4-121.3M) | 17.2M (15.3-18.8M) |
| Comm Scr | 10.7M (9.4-11.9M) | 131.6M (126.7-134.4M) | 22.1M (19.9-24.1M) |
| Impr Diag | 10.6M (9.1-11.8M) | 135.6M (130.6-138.5M) | 22.6M (20.3-24.5M) |
| DST for all | 10.6M (9.2-11.9M) | 135.7M (130.6-138.8M) | 22.8M (20.5-24.8M) |
| Short DS | 10.8M (9.5-12.1M) | 137.7M (132.6-140.7M) | 23M (20.7-25M) |
| Short DR | 10.6M (9.2-11.9M) | 133.9M (128.7-137.1M) | 22.7M (20.4-24.6M) |
| Pri Scr | 8.2M (7.2-9.2M) | - | - |
| Nutr | - | 124.1M (119.1-127.1M) | - |

2.10 Disability-adjusted life years averted compared to BAU (millions)

| Disability-adjusted life years averted (number relative to BAU) | | | |
|---|---|---|---|
| Intervention | Brazil | India | South Africa |
| Vacc | 2.9M (2.6-3.2M) | 19.3M (18-22.2M) | 5.7M (4.9-6.7M) |
| TPT | 0.1M (0.1-0.2M) | 6.1M (5.8-6.6M) | 0.9M (0.7-1M) |
| Comm Scr | 4.5M (4.1-5M) | 48.9M (46.5-50.3M) | 7.8M (7.1-8.5M) |
| DST for all | 0.2M (0-0.7M) | 1.9M (1.7-2.6M) | 0.2M (0.1-0.2M) |
| Short DS | 0M (0-0M) | 0M (0-0M) | 0M (0-0M) |
| Short DR | 0.2M (0-0.6M) | 3.6M (3.2-4.6M) | 0.3M (0.3-0.5M) |
| Pri Scr | 2.6M (2.2-3M) | - | - |
| Nutr | - | 13.4M (13-14.3M) | - |

### 3. ECONOMIC IMPACT

#### 3.1 Total cost of each intervention

| | Budget impact (USD) | | |
|---|---|---|---|
| Intervention | Brazil | India | South Africa |
| BAU | 1.6B (1.0-2.4B) | 17.5B (12.3-25.6B) | 2.5B (1.7-3.6B) |
| Vacc | 4.0B (2.6-5.5B) | 33.4B (23.8-44.6B) | 3.1B (2.2-4.2B) |
| TPT | 1.8B (1.2-3.0B) | 20.6B (15.3-29.0B) | 3.0B (2.2-4.1B) |
| Comm Scr (high) | 37.6B (22.6-60.4B) | 259.5B (165.7-370.2B) | 12.0B (8.4-17.5B) |
| Comm Scr (low) | 8.0B (5.7-10.8B) | 49.1B (36.6-63.7B) | 4.5B (3.4-5.8B) |
| Impr Diag | 2.1B (1.4-3.4B) | 29.0B (20.4-43.2B) | 2.7B (1.9-3.7B) |
| DST for all | 1.8B (1.1-2.9B) | 18.3B (13.0-26.5B) | 2.5B (1.7-3.6B) |
| Short DS | 2.9B (1.9-4.3B) | 32.5B (22.4-46.8B) | 4.4B (2.9-6.3B) |
| Short DR | 1.3B (0.8-1.9B) | 17.2B (12.0-25.2B) | 2.5B (1.6-3.5B) |
| Pri Scr | 1.8B (1.2-2.6B) | - | - |
| Nutr | - | 19.7B (14.5-28.0B) | - |

3.2 Incremental cost of each intervention compared to BAU

| | Incremental cost (USD relative to BAU) | | |
|---|---|---|---|
| Intervention | Brazil | India | South Africa |
| Vacc | 2.3B (1.3-3.8B) | 15.8B (8.9-25.4B) | 0.6B (0.3-1.0B) |
| TPT | 0.3B (0.1-0.7B) | 3.2B (2.4-4.3B) | 0.5B (0.4-0.7B) |
| Comm Scr (high) | 35.8B (20.8-58.8B) | 241.5B (146.9-353.6B) | 9.4B (5.9-14.9B) |
| Comm Scr (low) | 6.4B (4.4-9.0B) | 30.9B (21.4-42.8B) | 1.9B (1.4-2.7B) |
| Impr Diag | 0.5B (-0.2-1.4B) | 11.2B (0.8-25.2B) | 0.2B (-0.8-1.2B) |
| DST for all | 0.2B (0.1-0.7B) | 0.7B (0.5-1.2B) | 0.0B (0.0-0.0B) |
| Short DS | 1.3B (0.8-2.0B) | 14.8B (9.3-22.0B) | 1.9B (1.2-2.8B) |
| Short DR | -0.2B (-1.0--0.1B) | -0.3B (-0.6-0.0B) | 0.0B (-0.1-0.0B) |
| Pri Scr | 0.2B (0.1-0.3B) | - | - |
| Nutr | - | 2.2B (1.5-3.1B) | - |

##### 3.3 Cost–utility analysis by country

ICERs are calculated based on the mean costs and mean DALYs across the 500 model runs, hence these figures differ slightly from the median values shown in tables 2.9, 2.10, 3.1 and 3.2 above. The probability of being cost-effective at the lower or upper end of the estimated cost-effectiveness threshold range is based on the proportion of the 500 model runs that produced results giving an ICER below the relevant threshold value. For more on how the threshold ranges were selected see *Supplementary Material S3*, section 2.6; these ranges are not official policy thresholds in the respective countries.

###### 3.3a Brazil

| **Intervention** | **Mean total cost** (billion R$) | **Mean total cost** (billion US$) | **Incremental cost compared to BAU** (million US$) | **Mean total DALYs** (million) | **DALYs averted compared to BAU** (million) | **ICER compared to BAU** (US$ per DALY averted) | **Probability cost-effective at lower end of threshold range** | **Probability cost-effective at upper end of threshold range** |
|---|---|---|---|---|---|---|---|---|
| Business as usual | 8.04 | 1.61 | - | 10.80 | - | - | - | - |
| Vaccination | 19.84 | 3.98 | 2,365 | 7.87 | 2.93 | 807 | 100% | 100% |
| TB preventive treatment | 9.62 | 1.93 | 317 | 10.67 | 0.13 | 2,422 | 96% | 100% |
| Community screening (high-cost) | 192.29 | 38.54 | 36,924 | 6.29 | 4.52 | 8,174 | 31% | 87% |
| Community screening (low-cost) | 40.42 | 8.10 | 6,490 | 6.29 | 4.52 | 1,437 | 100% | 100% |

| | | | | | | | | |
|---|---|---|---|---|---|---|---|---|
| Improved diagnosis | 10.89 | 2.18 | 572 | 10.52 | 0.28 | 2,040 | 97% | 100% |
| Drug susceptibility testing for all | 9.27 | 1.86 | 247 | 10.58 | 0.22 | 1,103 | 100% | 100% |
| Shorter DS-TB treatment | 14.84 | 2.97 | 1,363 | 10.80 | 0 | Dominated by BAU | 0% | 0% |
| Shorter DR-TB treatment | 6.36 | 1.27 | -336 | 10.58 | 0.22 | Dominates BAU | 100% | 100% |
| Screening in prisons | 8.99 | 1.80 | 191 | 8.17 | 2.64 | 72 | 100% | 100% |

3.4b India

| **Intervention** | **Mean total cost** (trillion INR) | **Mean total cost** (billion US$) | **Incremental cost compared to BAU** (billion US$) | **Mean total DALYs** (million) | **DALYs averted compared to BAU** (million) | **ICER compared to BAU** (US$ per DALY averted) | **Probability cost-effective at lower end of threshold range** | **Probability cost-effective at upper end of threshold range** |
|---|---|---|---|---|---|---|---|---|
| Business as usual | 1.48 | 17.92 | - | 137.3 | - | - | - | - |
| Vaccination | 2.80 | 33.86 | 15.95 | 117.6 | 19.6 | 813 | 2% | 12% |
| TB preventive treatment | 1.75 | 21.10 | 3.18 | 131.2 | 6.1 | 523 | 7% | 74% |
| Community screening (high-cost) | 21.69 | 261.96 | 244.05 | 88.5 | 48.8 | 5,005 | 0% | 0% |
| Community screening (low-cost) | 4.06 | 49.09 | 31.17 | 88.5 | 48.8 | 639 | 1% | 25% |
| Improved diagnosis | 2.44 | 29.43 | 11.51 | 135.1 | 2.1 | 5,379 | 3% | 3% |
| Drug susceptibility testing for all | 1.55 | 18.68 | 0.76 | 135.3 | 2.0 | 390 | 60% | 94% |

| | | | | | | | | |
|---|---|---|---|---|---|---|---|---|
| Shorter DS-TB treatment | 2.73 | 32.92 | 15.00 | 137.3 | 0 | Dominated by BAU | 0% | 0% |
| Shorter DR-TB treatment | 1.46 | 17.61 | -0.31 | 133.6 | 3.7 | Dominates BAU | 100% | 100% |
| Nutrition | 1.67 | 20.17 | 2.25 | 123.8 | 13.5 | 167 | 100% | 100% |

3.4c South Africa

| **Intervention** | **Mean total cost** (billion R) | **Mean total cost** (billion US$) | **Incremental cost compared to BAU** (million US$) | **Mean total DALYs** (million) | **DALYs averted compared to BAU** (million) | **ICER compared to BAU** (US$ per DALY averted) | **Probability cost-effective at lower end of threshold range** | **Probability cost-effective at upper end of threshold range** |
|---|---|---|---|---|---|---|---|---|
| Business as usual | 47.22 | 2.56 | - | 22.94 | - | | - | - |
| Vaccination | 57.95 | 3.14 | 582 | 17.18 | 5.76 | 101 | 100% | 100% |
| TB preventive treatment | 56.92 | 3.09 | 526 | 22.06 | 0.88 | 599 | 100% | 100% |
| Community screening (high-cost) | 227.73 | 12.34 | 9,784 | 13.91 | 9.03 | 1,083 | 100% | 100% |
| Community screening (low-cost) | 83.44 | 4.52 | 1,963 | 13.91 | 9.03 | 217 | 100% | 100% |
| Improved diagnosis | 50.46 | 2.74 | 176 | 22.52 | 0.42 | 420 | 93% | 99% |
| Drug susceptibility testing for all | 47.62 | 2.58 | 22 | 22.78 | 0.16 | 137 | 100% | 100% |

| | | | | | | | | |
|---|---|---|---|---|---|---|---|---|
| Shorter DS-TB treatment | 82.72 | 4.48 | 1,924 | 22.94 | 0 | Dominated by BAU | 0% | 0% |
| Shorter DR-TB treatment | 46.53 | 2.52 | -37 | 22.60 | 0.34 | Dominates BAU | 100% | 100% |

3.4 Cost-effectiveness planes for all interventions (including high-cost community-wide screening)

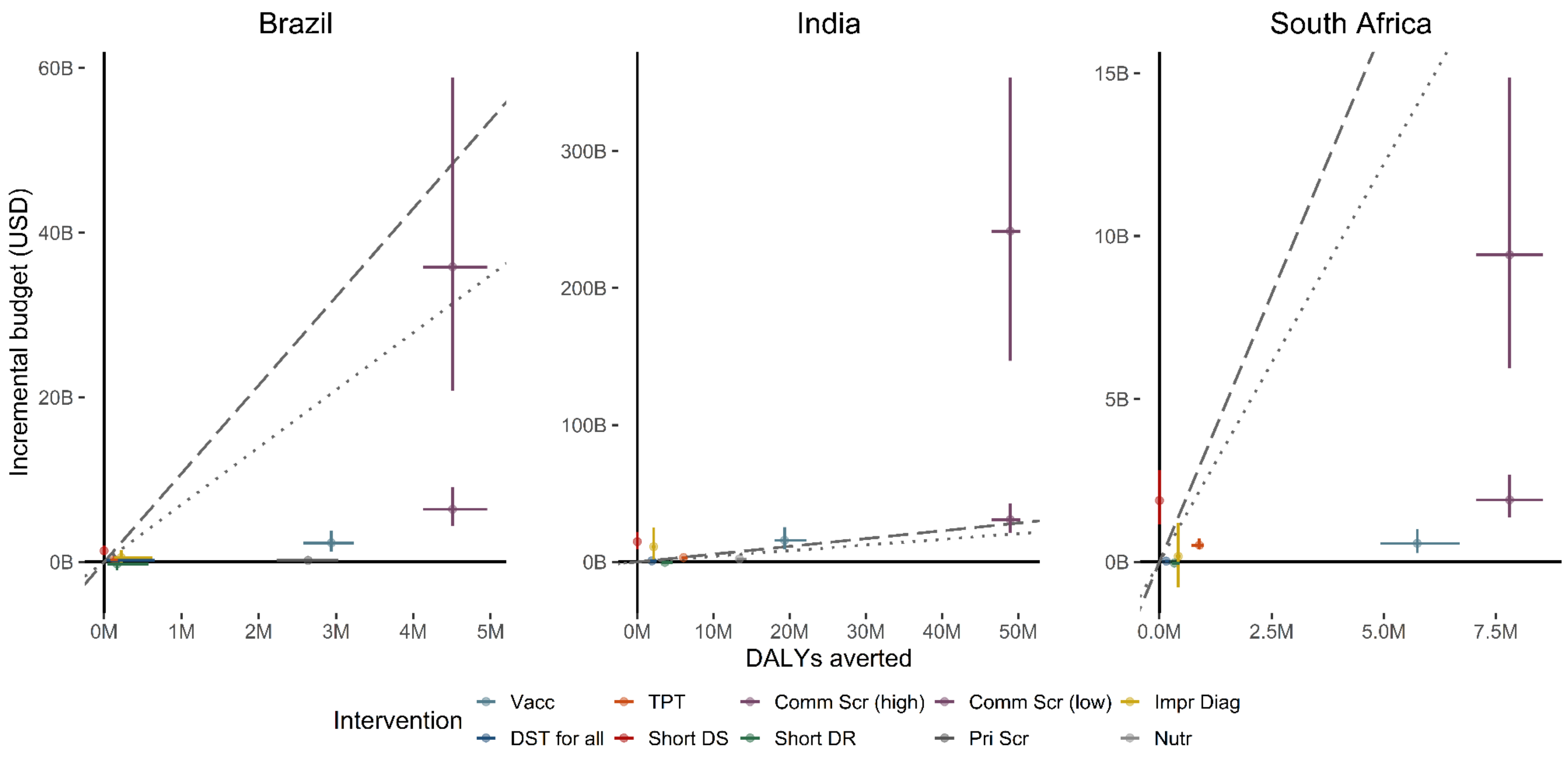

The dashed line shows the upper bound and the dotted line the lower bound of the estimated cost-effectiveness threshold range for each country. For a version of these graphs excluding high-cost community screening and showing the remaining interventions in more detail, see Figure 4 in the main paper.

### 4. Sensitivity analyses

#### 4.1 Aligned regression and progression parameters between India and South Africa

We transferred the posterior parameters for TB natural history variables (*infsub, nonsub, nonrec and infclr*) for the South Africa calibration to India. While the calibration targets were not met, the proportional epidemiological impact for vaccination was now similar for India and South Africa.

| Reduction in incident symptomatic TB in 2050 | | |
|---|---|---|
| | IND | ZAF |
| Vaccination | 30.3% (29.1–35.0) | 48.4% (43.3–53.2) |
| Sensitivity analysis | 43.2% (NA–NA) | NA |
